# Genetic correlation and causality of cancers and Parkinson’s disease

**DOI:** 10.1101/2020.10.07.20208124

**Authors:** Konstantin Senkevich, Sara Bandres-Ciga, Eric Yu, Upekha E. Liyanage, International Parkinson Disease Genomics Consortium (IPDGC), Alastair J Noyce, Ziv Gan-Or

**Affiliations:** Montreal Neurological Institute, McGill University, Montréal, QC, H3A 1A1, Canada; Department of Neurology and neurosurgery, McGill University, Montréal, QC, H3A 0G4, Canada, Canada; Molecular Genetics Section, Laboratory of Neurogenetics, NIA, NIH, Bethesda, MD, USA; Department of Human Genetics, McGill University, Montréal, QC, H3A 1A1, Canada; Cancer and Population Studies group, Population Health department, QIMR Berghofer Medical Research Institute, Locked Bag 2000, Royal Brisbane Hospital, Queensland 4006, Australia; Preventive Neurology Unit, Wolfson Institute of Preventive Medicine, Queen Mary University of London, London, UK; Department of Clinical and Movement Neurosciences, University College London Institute of Neurology, London, UK

**Keywords:** Parkinson’s disease, Genetic correlation, Mendelian randomization, Cancer, Melanoma

## Abstract

**Background and objectives:** Most cancers appear with reduced frequency in Parkinson’s disease (PD), but the prevalence of melanoma and brain cancers are often reported to be increased. Shared genetic architecture and causal relationships to explain these associations have not been fully explored.

**Methods:** Linkage disequilibrium score regression (LDSC) was applied for five cancer studies with available genome-wide association studies (GWAS) summary statistics to examine genetic correlations with PD. Additionally, we used GWAS summary statistics of 15 different types of cancers as exposures and two-sample Mendelian randomization to study the causal relationship with PD (outcome).

**Results:** LDSC analysis revealed a potential genetic correlation between PD and melanoma, breast cancer and prostate cancer. There was no evidence to support a causal relationship between the studied cancers and PD.

**Conclusions:** Our results suggest shared genetic architecture between PD and melanoma, breast, and prostate cancers, but no obvious causal relationship between cancers and PD.

## Introduction

Parkinson’s disease (PD) is a complex disorder, influenced by numerous environmental and genetic factors. Observational studies have suggested associations between PD and different types of cancers (lung, skin, pancreatic cancers and others),^1-7^ such that overall PD is associated with a reduced risk of cancer.^1, 2^ However, the prevalence of melanoma and brain tumors may be increased in patients with PD.^3-6^ In the absence of a causal effect, positive associations may be explained by confounding factors (such as toxins that casually influence the risk of specific cancers and PD), shared genetic susceptibility or biological pathways, or ascertainment bias (PD and cancer patients both being under regular medical follow-up).^8, 9^

Linkage disequilibrium (LD) score regression (LDSC) can be used to estimating heritability using summary statistics from genome-wide association studies (GWASs) and examine correlations between two traits occurring through shared genetic architecture.^10^ Mendelian randomization (MR) uses SNPs associated with an exposure of interest (such as cancer) as proxies to determine the causal association between that exposure and an outcome (in this case PD).^11^ In the current study, we used LDSC and MR to examine whether certain types of cancers have genetic correlation or causal relationships with PD.

## Methods

### Linkage disequilibrium score regression

To investigate whether there is overlapping genetic etiology between PD and the studied cancers we performed a search on the GWAS Catalog^12^ for publicly available full summary statistics using keywords “cancer”, “carcinoma”, “glioma”, “lymphoma”, “leukemia”, “melanoma” and selected GWASs with a minimum of 1000 cases and of European ancestry. Additionally, we contacted authors and requested for full summary statistics. Overall, we were able to collect full summary statistics for melanoma,^13^ breast,^14^ prostate,^15^ endometrial^16^ and keratinocytes cancers (basal cell carcinoma and squamous cell carcinoma).^17^ Keratinocytes cancer summary statistics include meta-analysis of QSkin, eMERGE and UK Biobank (UKB) cohorts with a total of 28,218 cases and 353,855 controls. Of the cancer studies with full summary statistics, endometrial cancer, melanoma and keratinocyte cancer studies included data from the UKB. We also used GWAS summary statistics from the latest PD GWAS excluding 23andMe and UKB data, to avoid potential bias due to overlapping samples.^18^ After the exclusions, a total of 15,056 PD patients and 12,637 controls were included in the summary statistics.^19^ We utilized the LDSC method as previously described.^10, 20^ Summary statistics were formatted using the standard settings of the munge_sumstats.py script.^10^

### Mendelian randomization

#### Exposure data

For the construction of genetic instruments, we selected studies from the GWAS Catalog^12^ using the R package MRInstruments.^21, 22^ First, we searched for traits using the same keywords as before. We then selected the most recent available GWAS for each cancer, with a minimum of 1000 cases and at least the same number of controls of European ancestry. Additionally, recent GWASs on melanoma^13^ and combined analysis of keratinocyte cancers^17^ were added as they were not available in the GWAS catalog. Fifteen studies were selected for this part of the analysis **(Supplementary Table 1)**. UKB participants were included in some of these studies (colorectal cancer, combined analysis of keratinocyte cancers, endometrial cancer, lung cancer, melanoma, uterine fibroids).

**Table 1.** Genetic correlations using LD Score regression between PD and cancers.

| Trait 1 | Trait 2 | rg | SE | Z | P-value |
| --- | --- | --- | --- | --- | --- |
| PD | Prostate cancer | 0.0949 | 0.0486 | 1.9547 | 0.0506 |
| PD | Breast cancer | -0.1476 | 0.0698 | -2.1156 | 0.0344 |
| PD | Endometrial cancer | -0.0126 | 0.0764 | -0.1645 | 0.8693 |
| PD | Keratinocytes cancers | 0.0008 | 0.0574 | 0.0143 | 0.9886 |
| PD | Melanoma | 0.1265 | 0.0644 | 1.9641 | 0.0495 |

PD, Parkinson's disease; rg, genetic correlation; SE, standard error; P-value not corrected

We constructed genetic instruments for cancer susceptibility using SNPs with GWAS significant *p*-values (<5×10^−8^) from each study. The extracted data included rs-numbers, log odds ratios, standard errors (SE), *p*-values, effect alleles, and effect allele frequency. SNPs for each exposure were clumped using standard parameters (clumping window of 10,000 kb, r^2^ cutoff 0.001) to discard variants in LD. Additionally, we calculated r^2^, which reflects the proportion of variability explained by genetic variants and F-statistics to estimate the strength of IVs selected for exposures as previously described.^23, 24^

#### Outcome data

As an outcome, we used summary statistics data from the latest PD GWAS.^19^ To minimize the risk of overlap in participants between studies, which could create bias, again we excluded 23andMe and UKB data.

#### Power calculation

We calculated estimated power to detect an equivalent effect size of OR 1.2 on PD risk utilizing an online Mendelian randomization power calculation (https://sb452.shinyapps.io/power/).^25^

#### Mendelian randomization analyses

MR methods implemented in the Two-sample MR R package^21, 22^ were used and are described in detail elsewhere.^26-28^ Firstly, we performed Steiger filtering to exclude SNPs that explain more variance in the outcome than in the exposure.^22^ We then used the inverse variance weighted (IVW) method, in which we pooled estimates from individual Wald ratios for each SNP and meta-analyzed using random effects.^26-28^ We applied MR Egger to detect net directional pleiotropy and provide a better estimate of the true causal effect allowing to detect possible violations of instrumental variable assumptions.^28^ Additionally, we used weighted median (WM) which is a median of the weighted estimates and provides consistent effect even if 50% of IVs are invalid.^29^ These sensitivity analyses were performed to explore heterogeneity and horizontal pleiotropy. Heterogeneity was tested using Cochran’s Q test in the IVW and MR-Egger methods.^30^ For each method, we constructed funnel plots to detect pleiotropic outliers **(Supplementary Figure 1-6)**. Additionally, we performed MR-PRESSO test to detect outlier SNPs which may be biasing estimates through horizontal pleiotropy, and adjust for this.^31^

#### Data availability

All code used in the current study is available at our GitHub at https://github.com/gan-orlab/MR_Cancers-PD

## Results

Genetic correlation with LDSC was performed for melanoma, breast, prostate, endometrial and keratinocytes cancers. We observed nominal genetic correlations with PD for prostate cancer (rg=0.095; *p*=0.051), breast cancer (rg=-0.148; *p*=0.034) and melanoma (rg=0.127; *p*=0.049) **(Table 1; Supplementary Table 2)**. Although bias might be a concern, LDSC intercepts were close to one in both PD and all the assessed cancer summary statistics.

**Table 2.**
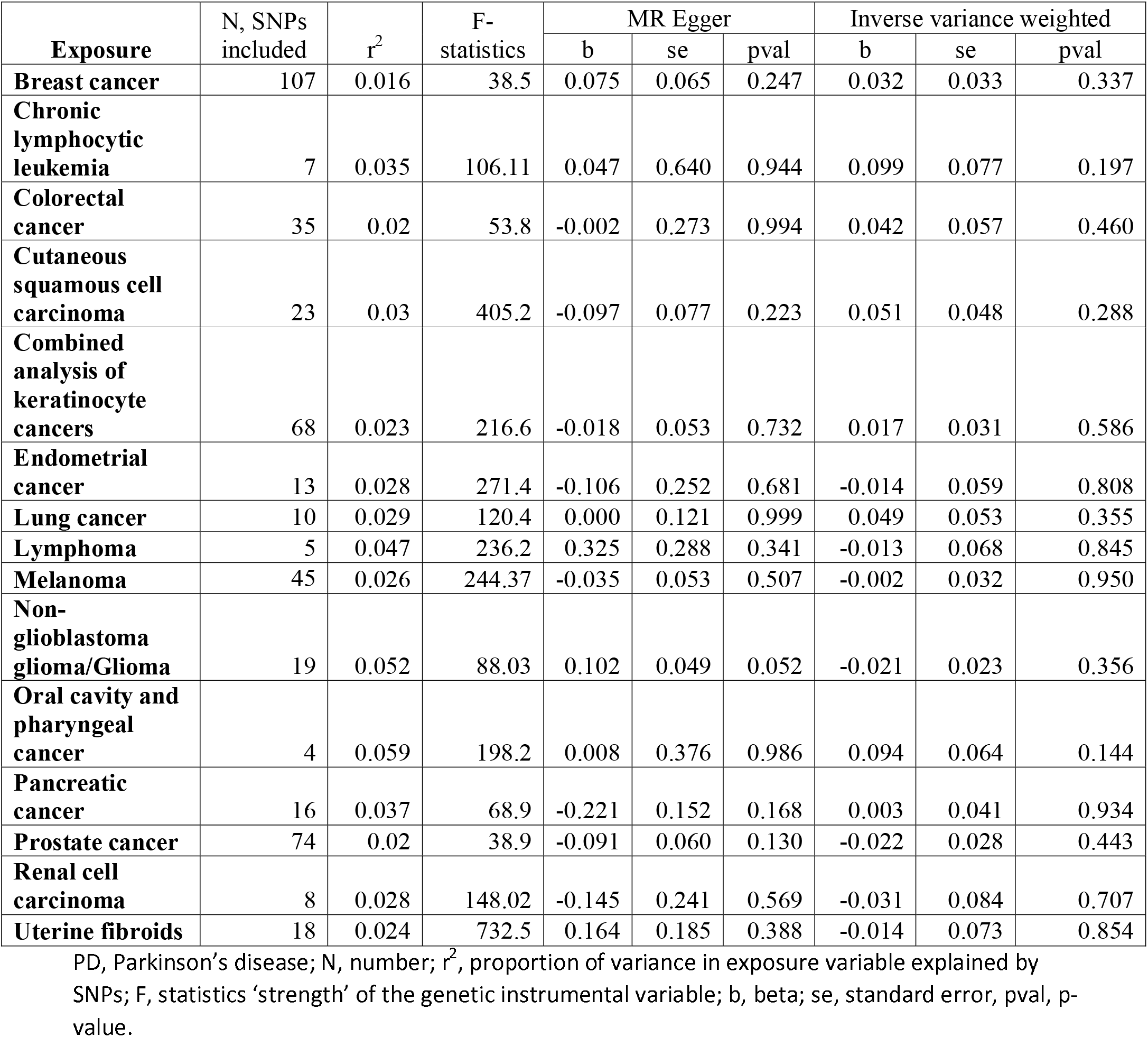
MR analysis between exposure (cancers) and outcome (PD), PD GWAS summary statistics excluding 23andMe and UK biobank data.

| Exposure | N, SNPs included | $r^2$ | F-statistics | MR Egger | | | Inverse variance weighted | | |
| --- | --- | --- | --- | --- | --- | --- | --- | --- | --- |
|  |  |  |  | b | se | pval | b | se | pval |
| **Breast cancer** | 107 | 0.016 | 38.5 | 0.075 | 0.065 | 0.247 | 0.032 | 0.033 | 0.337 |
| **Chronic lymphocytic leukemia** | 7 | 0.035 | 106.11 | 0.047 | 0.640 | 0.944 | 0.099 | 0.077 | 0.197 |
| **Colorectal cancer** | 35 | 0.02 | 53.8 | -0.002 | 0.273 | 0.994 | 0.042 | 0.057 | 0.460 |
| **Cutaneous squamous cell carcinoma** | 23 | 0.03 | 405.2 | -0.097 | 0.077 | 0.223 | 0.051 | 0.048 | 0.288 |
| **Combined analysis of keratinocyte cancers** | 68 | 0.023 | 216.6 | -0.018 | 0.053 | 0.732 | 0.017 | 0.031 | 0.586 |
| **Endometrial cancer** | 13 | 0.028 | 271.4 | -0.106 | 0.252 | 0.681 | -0.014 | 0.059 | 0.808 |
| **Lung cancer** | 10 | 0.029 | 120.4 | 0.000 | 0.121 | 0.999 | 0.049 | 0.053 | 0.355 |
| **Lymphoma** | 5 | 0.047 | 236.2 | 0.325 | 0.288 | 0.341 | -0.013 | 0.068 | 0.845 |
| **Melanoma** | 45 | 0.026 | 244.37 | -0.035 | 0.053 | 0.507 | -0.002 | 0.032 | 0.950 |
| **Non-glioblastoma glioma/Glioma** | 19 | 0.052 | 88.03 | 0.102 | 0.049 | 0.052 | -0.021 | 0.023 | 0.356 |
| **Oral cavity and pharyngeal cancer** | 4 | 0.059 | 198.2 | 0.008 | 0.376 | 0.986 | 0.094 | 0.064 | 0.144 |
| **Pancreatic cancer** | 16 | 0.037 | 68.9 | -0.221 | 0.152 | 0.168 | 0.003 | 0.041 | 0.934 |
| **Prostate cancer** | 74 | 0.02 | 38.9 | -0.091 | 0.060 | 0.130 | -0.022 | 0.028 | 0.443 |
| **Renal cell carcinoma** | 8 | 0.028 | 148.02 | -0.145 | 0.241 | 0.569 | -0.031 | 0.084 | 0.707 |
| **Uterine fibroids** | 18 | 0.024 | 732.5 | 0.164 | 0.185 | 0.388 | -0.014 | 0.073 | 0.854 |

PD, Parkinson's disease; N, number; $r^2$ , proportion of variance in exposure variable explained by SNPs; F, statistics 'strength' of the genetic instrumental variable; b, beta; se, standard error, pval, p-value.

### Mendelian randomization does not support a causal role for different cancers and PD

We performed MR with the five cancers for which we had full summary statistics. We further included 10 additional cancers with limited summary statistics, resulting in 15 cancers being included in this part of the analysis **(Supplementary Table 1)**. The variance in the exposure variables explained by SNPs ranged from 0.016 to 0.059 **(Table 2)**. All instruments had F-statistics of >10, which is the cut-off that most studies apply to indicate sufficient instrument strength **(Table 2; Supplementary Table 3)**. No causal effect of any cancer on PD was observed **(Table 2; Supplementary Table 3, Supplementary Figure 1-2)**. Significant heterogeneity was apparent for cutaneous squamous cell carcinoma (IVW, Q *p*-value=0.02) and combined analysis of keratinocyte cancers (MR Egger, Q *p*-value=0.012; IVW, Q *p*-value=0.012, **Supplementary Table 4, Supplementary Figure 3**). There was some evidence for net horizontal pleiotropy for brain tumors (*p*=0.011) and cutaneous squamous cell carcinoma (*p*=0.029, **Supplementary Table 4**) which may have resulted in bias to IVW estimates, but the slopes from Egger regression were imprecisely estimated. MR-PRESSO identified potential outliers for cutaneous squamous cell carcinoma **(Supplementary Table 4)**. The distortion test did not suggest significant changes in the effect estimates after these outliers were removed **(Supplementary Table 4)**. The sensitivity analyses revealed no clear evidence for bias in the IVW estimate due to invalid instruments with other cancers.

Additionally, we performed reverse MR using PD-associated SNPs as exposure and cancer summary statistics as outcome and did not find any evidence of causal relationship **(Supplementary Table 5; Supplementary Figure 4-6)**.

## Discussion

In the current study, we performed a comprehensive analysis of the genetic correlation and causal relationships between various cancers and PD. We found a nominally significant genetic correlation between PD and both prostate and breast cancers. These cancers are mainly sex-specific and thus, our results could be biased. The genetic correlation between PD and sex-specific cancers should be tested with sex-stratified outcome data, which was not available for the current analysis. The most thoroughly studied genetic relationship is between melanoma and PD.^32^ Thus far, there is no strong genetic evidence for an association between melanoma-related genes and PD.^33-35^ Previous MR studies also did not demonstrate evidence of a causal relationship between PD and melanoma.^23^ However, a recent, comprehensive LDSC-based analysis of genetic correlation suggested a significant genetic correlation between melanoma and PD, with gene expression overlap.^8^ Together with our current results, it is possible that genetic correlation between PD and melanoma, and hence shared biological pathways, explains the increased frequency of melanoma in PD patients and the increased frequency of PD in melanoma patients.^3, 36^

Our results do not support a causal relationship between the cancers we studied and PD. Although MR can help reduce confounding and the possibility of reverse causality, a recent study demonstrated that MR is not immune to survival bias.^37^ PD is mainly an elderly disease, thus it could also be prone to survival biases. On the other hand, early mortality from cancer could reduce cancer prevalence in PD.^38^ It is likely that the low incidence of most cancers observed in PD is due to survival biases. The high incidence of brain cancers in PD might be related to closer medical attention (i.e. more frequent MRI in PD patients compared to the general population).

Our study has several limitations. We had a limited number of available full summary statistics for cancers. Thus, we were not able to perform LDSC between all cancers with known associations with PD based on epidemiological studies. Another limitation is that this is European-based study only, and these associations or lack thereof should be studied in other populations. In both LDSC and MR analyses, we excluded UKB data to decrease the chances of overlapping samples between studies, which reduced the statistical power in the different analyses. Lack of availability of sex-specific PD GWAS data is the another limitation, which would be crucial for studying genetic correlations with sex-specific cancers, or with cancers that have meaningful sex differences.^39^ Despite the fact that some cancers are more prevalent in PD patients, it was not possible to perform bi-directional MR for all of the cancer studies since full summary statistics were not available. However, we performed bi-directional MR with PD and cancers with available full summary statistics and did not find evidence of causal relationships.

To conclude, our results do not support a causal relationship between the tested cancers and PD and confirm a possible genetic correlation between melanoma and PD. Sex stratified PD data should be used to verify possible shared genetic architecture with breast and prostate cancers. Once larger datasets become available, as well as sex-specific PD datasets, additional LDSC and MR studies should be performed on cancers and PD.

## Acknowledgments

We would like to thank the relevant consortia for making their data available. We would like to also thank all members of the International Parkinson Disease Genomics Consortium (IPDGC). For a complete overview of members, acknowledgements and funding, please see http://pdgenetics.org/partners. ZGO is supported by the Fonds de recherche du Québec – Santé (FRQS) Chercheurs-boursiers award, in collaboration with Parkinson Quebec, and by the Young Investigator Award by Parkinson Canada. KS is supported by a postdoctoral fellowship from the Canada First Research Excellence Fund (CFREF), awarded to McGill University for the Healthy Brains for Healthy Lives initiative (HBHL). We would like to also thank Stuart MacGregor, Matthew Law and David Whiteman from QIMR Berghofer Medical Research Institute, Locked Bag 2000, Royal Brisbane Hospital, Queensland 4006, Australia for providing summary statistics data on keratinocytes cancers. The endometrial cancer genome-wide association analyses were supported by the National Health and Medical Research Council of Australia (APP552402, APP1031333, APP1109286, APP1111246 and APP1061779), the U.S. National Institutes of Health (R01-CA134958), European Research Council (EU FP7 Grant), Wellcome Trust Centre for Human Genetics (090532/Z/09Z) and Cancer Research UK. OncoArray genotyping of ECAC cases was performed with the generous assistance of the Ovarian Cancer Association Consortium (OCAC), which was funded through grants from the U.S. National Institutes of Health (CA1×01HG007491-01 (C.I. Amos), U19-CA148112 (T.A. Sellers), R01-CA149429 (C.M. Phelan) and R01-CA058598 (M.T. Goodman); Canadian Institutes of Health Research (MOP-86727 (L.E. Kelemen)) and the Ovarian Cancer Research Fund (A. Berchuck). We particularly thank the efforts of Cathy Phelan. OncoArray genotyping of the BCAC controls was funded by Genome Canada Grant GPH-129344, NIH Grant U19 CA148065, and Cancer UK Grant C1287/A16563. All studies and funders are listed in O’Mara et al (2018). The breast cancer genome-wide association analyses were supported by the Government of Canada through Genome Canada and the Canadian Institutes of Health Research, the ‘Ministère de l’Économie, de la Science et de l’Innovation du Québec’ through Genome Québec and grant PSR-SIIRI-701, The National Institutes of Health (U19 CA148065, X01HG007492), Cancer Research UK (C1287/A10118, C1287/A16563, C1287/A10710) and The European Union (HEALTH-F2-2009-223175 and H2020 633784 and 634935). All studies and funders supported breast cancer GWAS are listed in Michailidou et al., (Nature, 2017). For acknowledgements for the melanoma meta-analysis see Landi et al (Nature genetics, 2020). We would like to thank The PRACTICAL consortium, CRUK, BPC3, CAPS, PEGASUS. The Prostate cancer genome-wide association analyses are supported by the Canadian Institutes of Health Research, European Commission’s Seventh Framework Programme grant agreement n° 223175 (HEALTH-F2-2009-223175), Cancer Research UK Grants C5047/A7357, C1287/A10118, C1287/A16563, C5047/A3354, C5047/A10692, C16913/A6135, and The National Institute of Health (NIH) Cancer Post-Cancer GWAS initiative grant: No. 1 U19 CA 148537-01 (the GAME-ON initiative). We would also like to thank the following for funding support: The Institute of Cancer Research and The Everyman Campaign, The Prostate Cancer Research Foundation, Prostate Research Campaign UK (now PCUK), The Orchid Cancer Appeal, Rosetrees Trust, The National Cancer Research Network UK, The National Cancer Research Institute (NCRI) UK. We are grateful for support of NIHR funding to the NIHR Biomedical Research Centre at The Institute of Cancer Research and The Royal Marsden NHS Foundation Trust. The Prostate Cancer Program of Cancer Council Victoria also acknowledge grant support from The National Health and Medical Research Council, Australia (126402, 209057, 251533,, 396414, 450104, 504700, 504702, 504715, 623204, 940394, 614296,), VicHealth, Cancer Council Victoria, The Prostate Cancer Foundation of Australia, The Whitten Foundation, PricewaterhouseCoopers, and Tattersall’s. EAO, DMK, and EMK acknowledge the Intramural Program of the National Human Genome Research Institute for their support. Genotyping of the OncoArray was funded by the US National Institutes of Health (NIH) [U19 CA 148537 for ELucidating Loci Involved in Prostate cancer SuscEptibility (ELLIPSE) project and X01HG007492 to the Center for Inherited Disease Research (CIDR) under contract number HHSN268201200008I] and by Cancer Research UK grant A8197/A16565. Additional analytic support was provided by NIH NCI U01 CA188392 (PI: Schumacher). Funding for the iCOGS infrastructure came from: the European Community’s Seventh Framework Programme under grant agreement n° 223175 (HEALTH-F2-2009-223175) (COGS), Cancer Research UK (C1287/A10118, C1287/A 10710, C12292/A11174, C1281/A12014, C5047/A8384, C5047/A15007, C5047/A10692, C8197/A16565), the National Institutes of Health (CA128978) and Post-Cancer GWAS initiative (1U19 CA148537, 1U19 CA148065 and 1U19 CA148112 – the GAME-ON initiative), the Department of Defence (W81XWH-10-1-0341), the Canadian Institutes of Health Research (CIHR) for the CIHR Team in Familial Risks of Breast Cancer, Komen Foundation for the Cure, the Breast Cancer Research Foundation, and the Ovarian Cancer Research Fund. The BPC3 was supported by the U.S. National Institutes of Health, National Cancer Institute (cooperative agreements U01-CA98233 to D.J.H., U01-CA98710 to S.M.G., U01-CA98216 toE.R., and U01-CA98758 to B.E.H., and Intramural Research Program of NIH/National Cancer Institute, Division of Cancer Epidemiology and Genetics). CAPS GWAS study was supported by the Swedish Cancer Foundation (grant no 09-0677, 11-484, 12-823), the Cancer Risk Prediction Center (CRisP; www.crispcenter.org), a Linneus Centre (Contract ID 70867902) financed by the Swedish Research Council, Swedish Research Council (grant no K2010-70X-20430-04-3, 2014-2269). PEGASUS was supported by the Intramural Research Program, Division of Cancer Epidemiology and Genetics, National Cancer Institute, National Institutes of Health. This research was supported in part by the Intramural Research Program of the NIH, National institute on Aging.

## Authors’ Roles

1. Research project: A. Conception, B. Organization, C. Execution;
2. Statistical Analysis: A. Design, B. Execution, C. Review and Critique;
3. Manuscript Preparation: A. Writing of the first draft, B. Review and Critique.

KS: 1A, 1B, 1C, 2A, 2B, 3A

AJN: 1A, 2C, 3B

EY: 1C, 2C, 3B

UEL: 1C, 2B, 3B

SBC: 2C, 3 B

ZGO: 1A, 1B, 2C, 3B

## Financial Disclosures of all authors (for the preceding 12 months)

ZGO has received consulting fees from Lysosomal Therapeutics Inc., Idorsia, Prevail Therapeutics, Denali, Ono Therapeutics, Neuron23, Handl Therapeutics, Deerfield and Inception Sciences (now Ventus). None of these companies were involved in any parts of preparing, drafting and publishing this study. AJN received grants from the Barts Charity, Parkinson’s UK and Aligning Science Across Parkinson’s; and honoraria from Britannia, BIAL, AbbVie, Global Kinetics Corporation, Profile, Biogen, and Roche. The rest of the authors have nothing to report.

