## Supplementary Figures 1-6 for "Genetic correlation and causality of cancers and Parkinson’s disease"

**Supplementary Figure 1. Forest plots showing point estimates of the exposures of interest, Exposure of interest at the top of each forest plot..…………………………………………………………………………………….......2**

**Supplementary Figure 2. Plots showing point estimates of the exposures of interest; Exposure of interest at the top of each plot……………………..……………………………………………………….……………….18**

**Supplementary Figure 3. Funnel plots evaluated the presence of possible heterogeneity across the estimates. Exposure of interest at the top of each plot……………………………………………………………………………………….…..34**

**Supplementary Figure 4. Reverse MR (PD as exposure; Cancers as outcome). Forest plots showing point estimates of the exposures of interest, Exposure of interest at the top of each forest plot..……………………………………………………..…………….............50**

**Supplementary Figure 5. Reverse MR (PD as exposure; Cancers as outcome). Plots showing point estimates of the exposures of interest; Exposure of interest at the top of each plot ……………………..………………………………………………………..………………..….56**

**Supplementary Figure 6. Reverse MR (PD as exposure; Cancers as outcome). Funnel plots evaluated the presence of possible heterogeneity across the estimates. Exposure of interest at the top of each plot…………………………………………………………………………..62**

**Supplementary Figure 1. PD without UKBB. Forest plots showing point estimates of the exposures of interest, Exposure of interest at the top of each forest plot.**

Black points represent log-odds ratio of each SNP on the risk of PD. Red points represent the log-odds ration when combining all SNPs together (Inverse variance weighted and MR Egger methods). Lines from points represent 95% confidence intervals.

Breast cancer as exposure

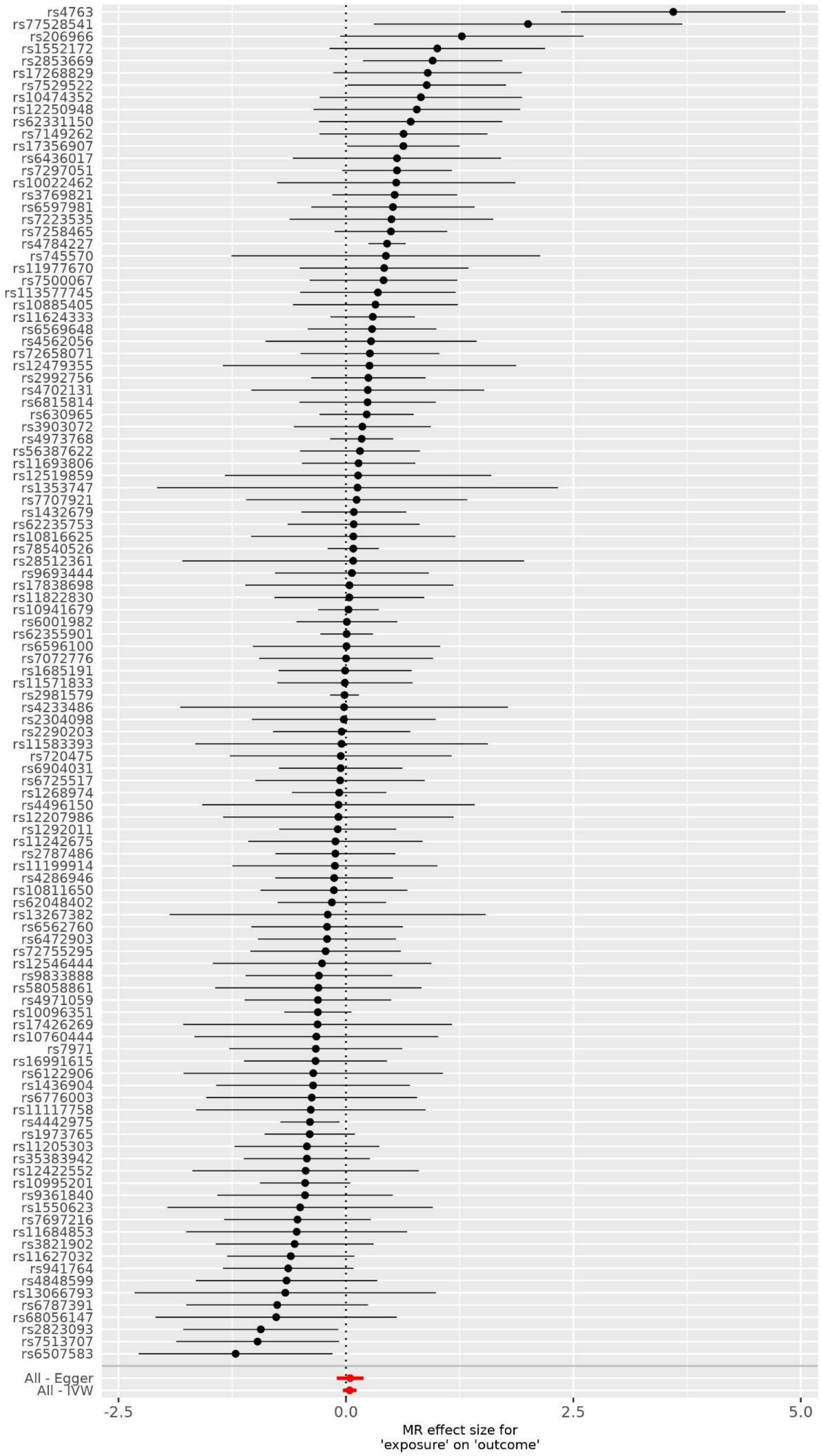

Chronic lymphocytic leukemia as exposure

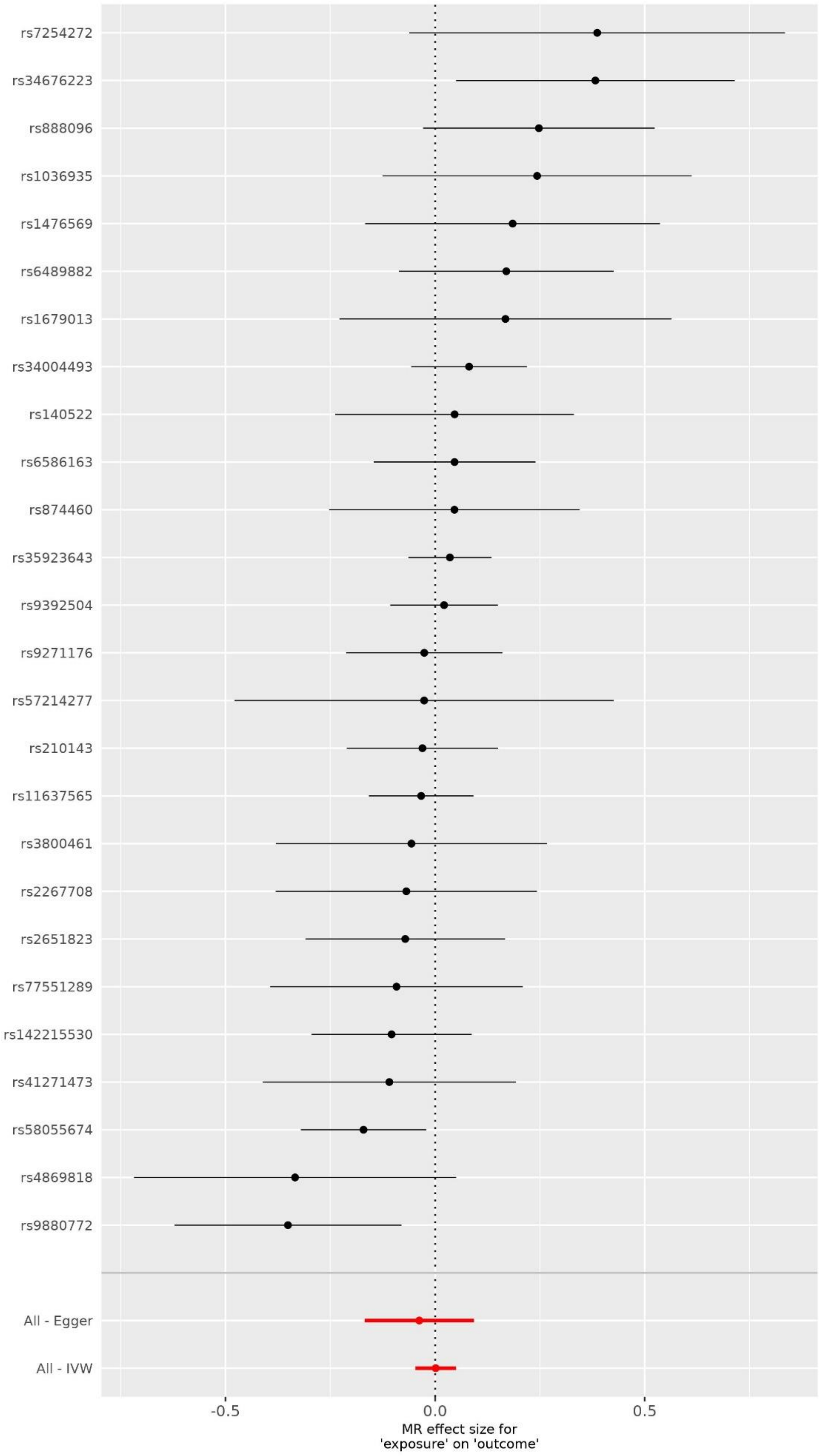

Colorectal cancer as exposure

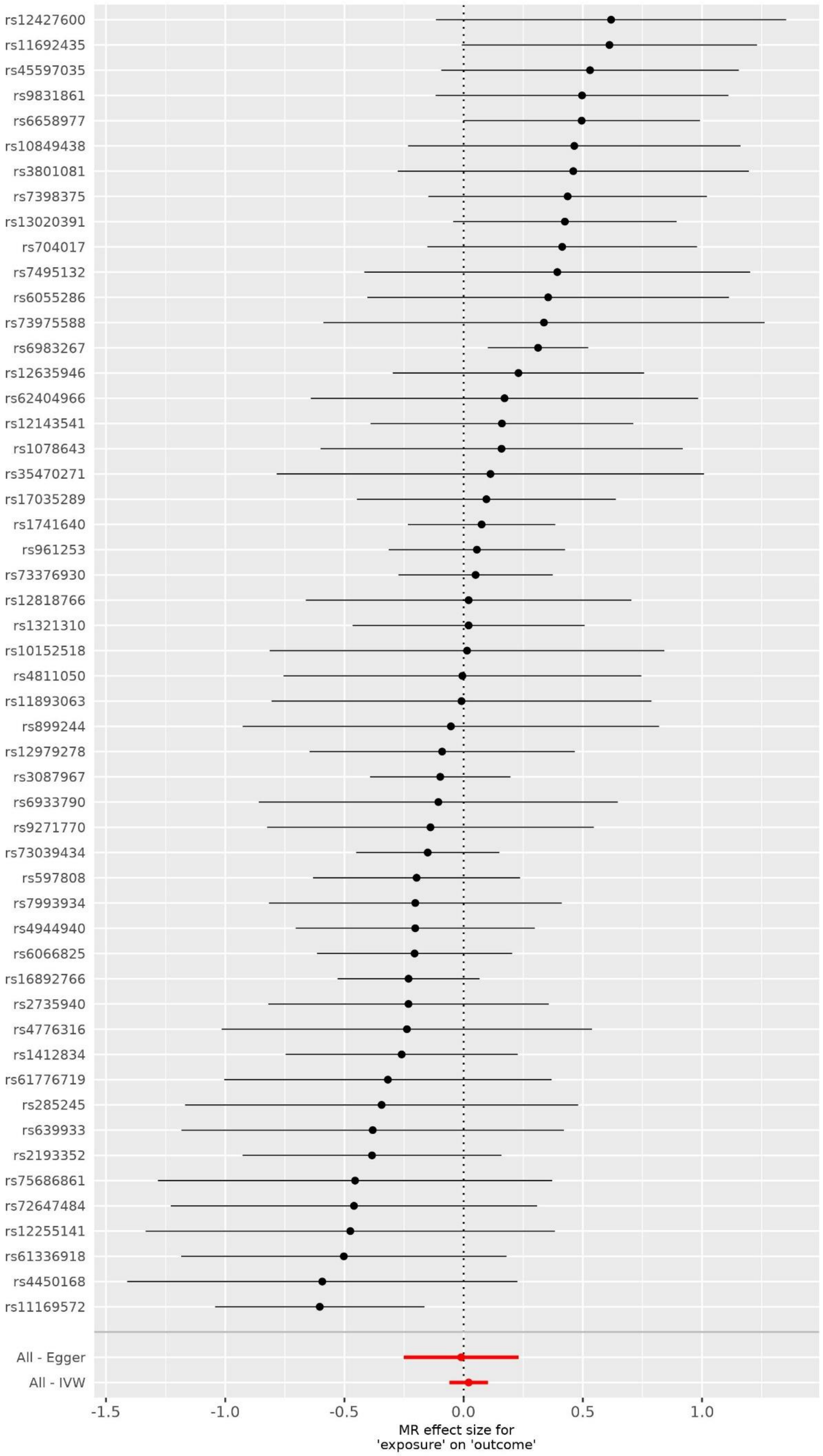

Cutaneous squamous cell carcinoma as exposure

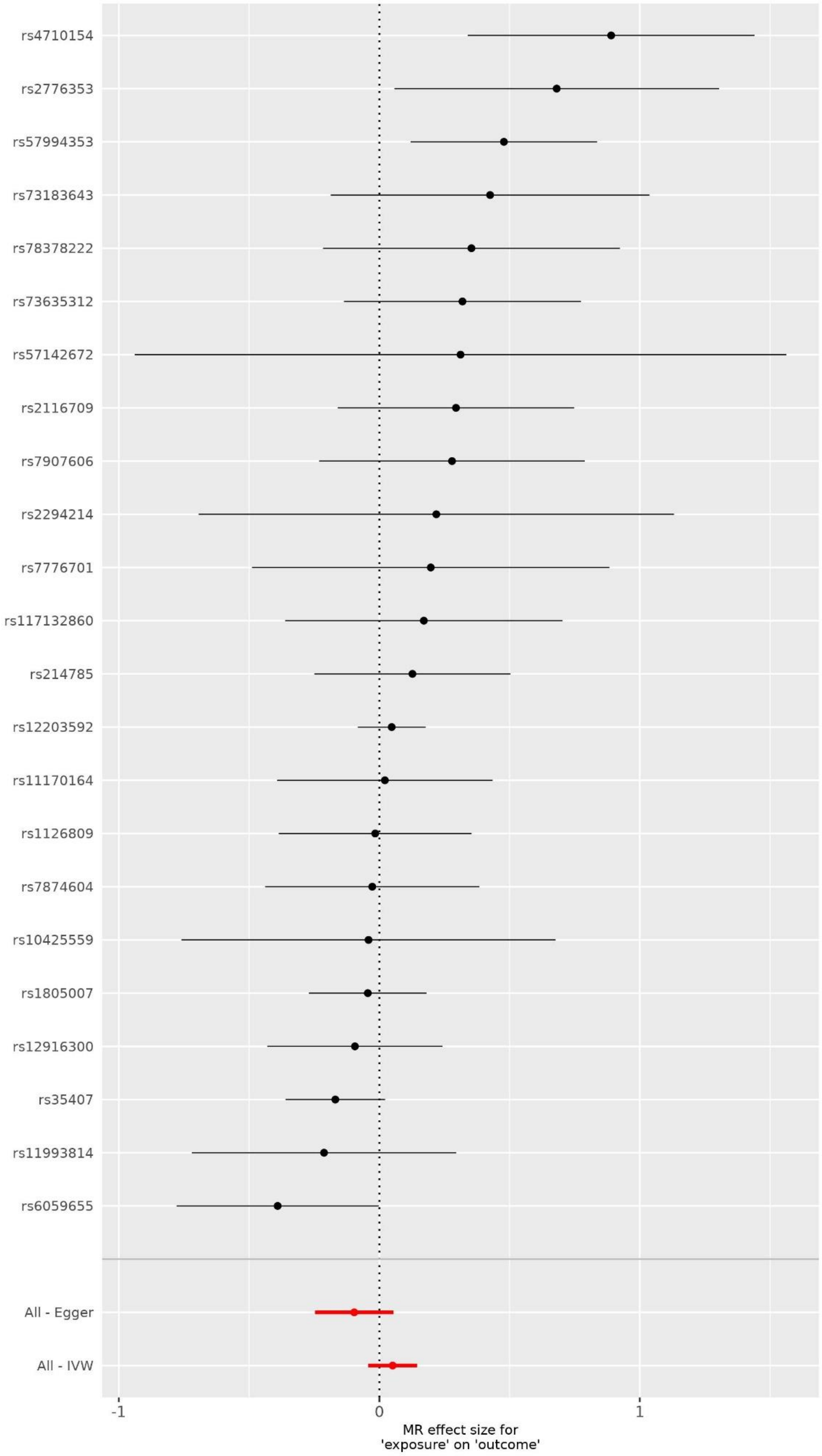

Combined analysis of keratinocyte cancers as exposure

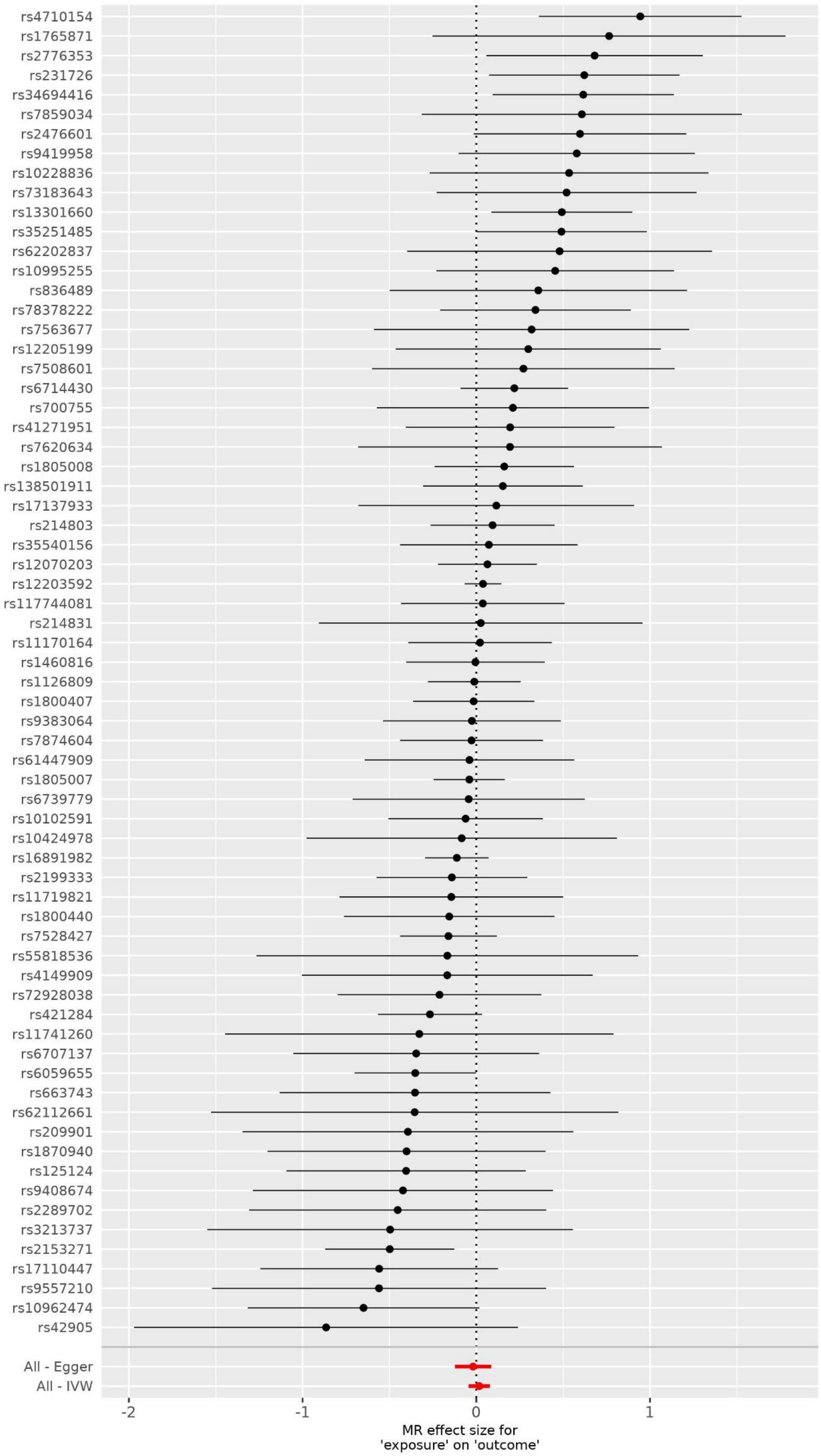

Endometrial cancer as exposure

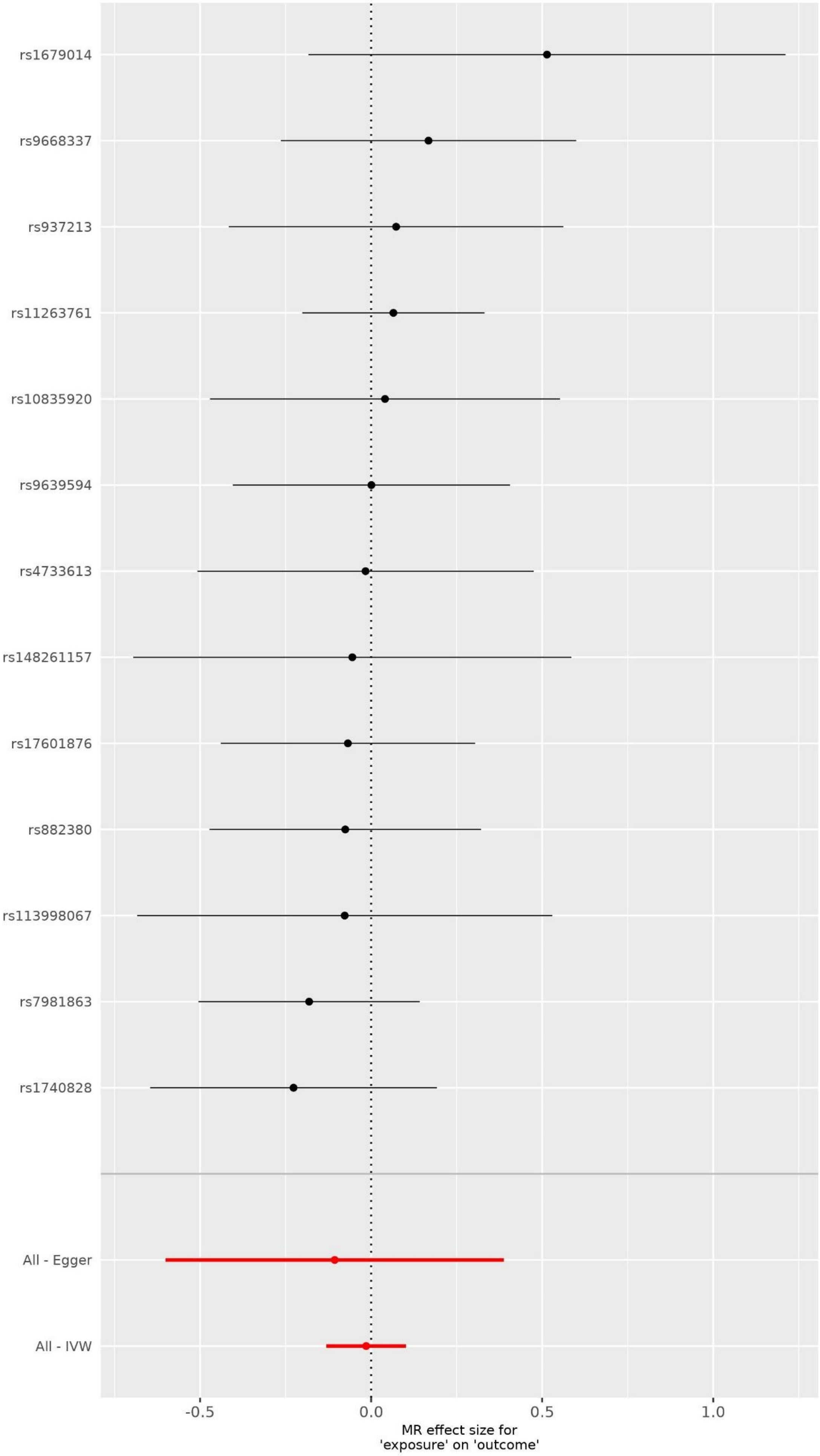

Lung cancer as exposure

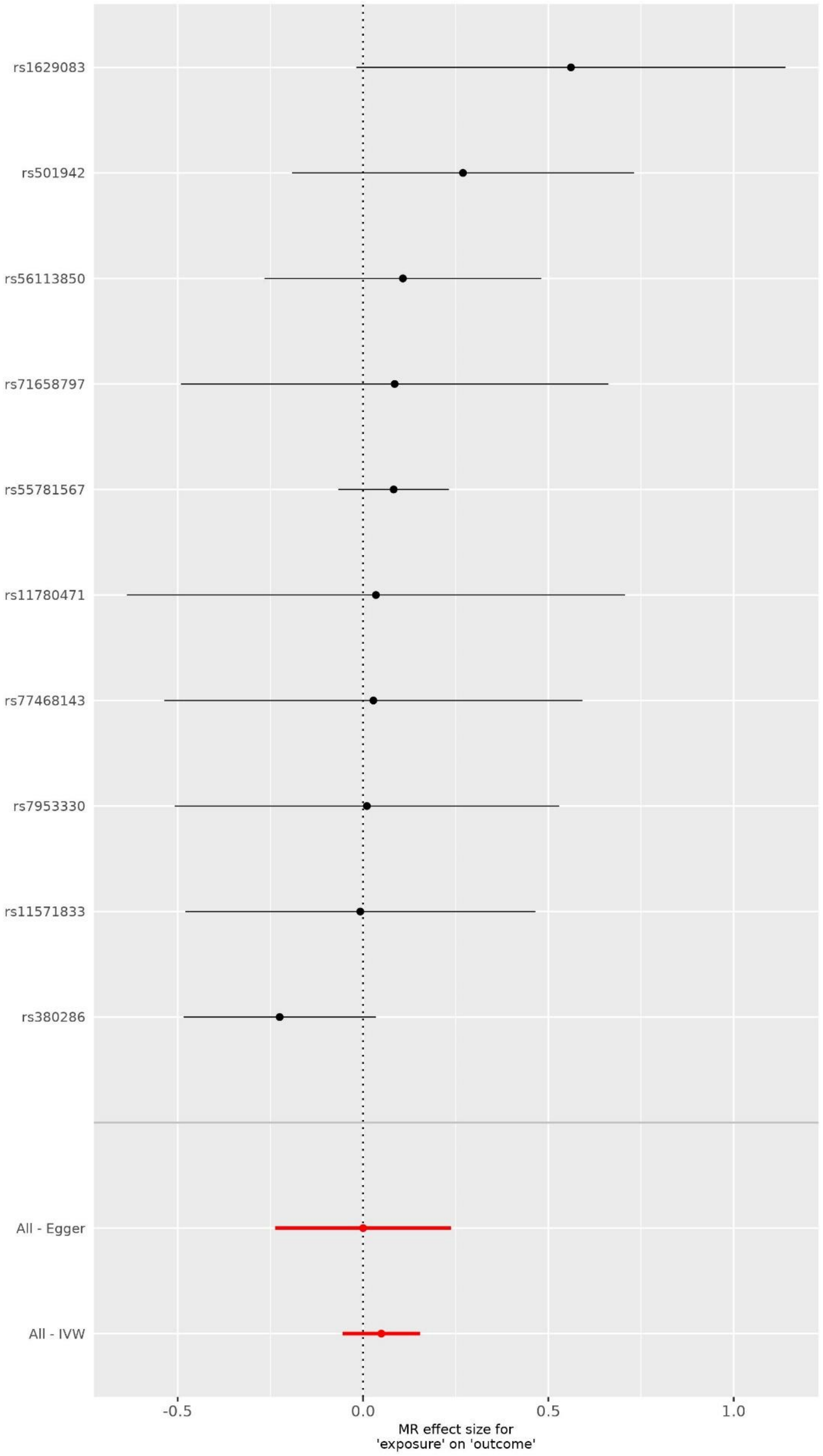

Lymphoma as exposure

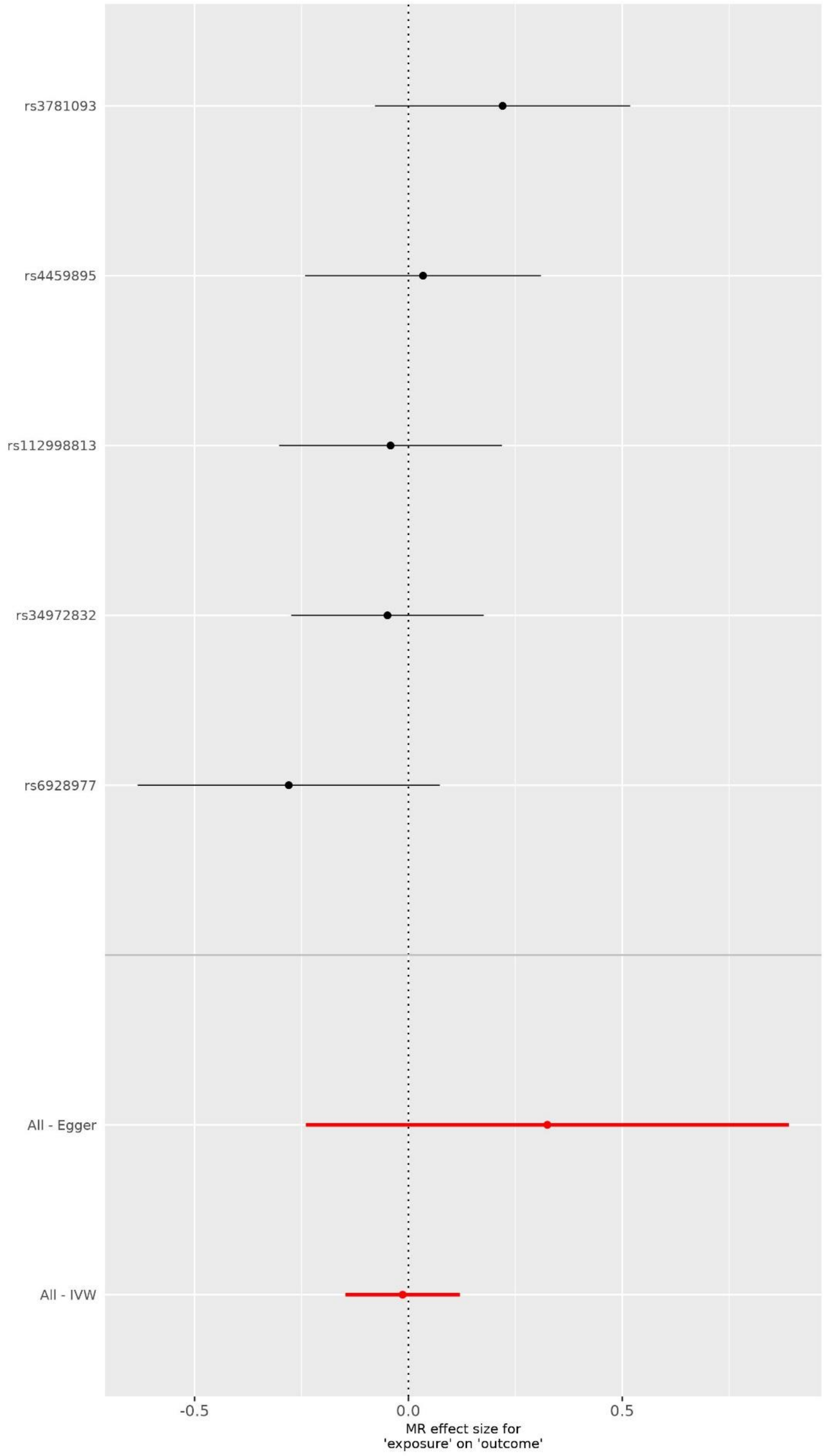

Melanoma as exposure

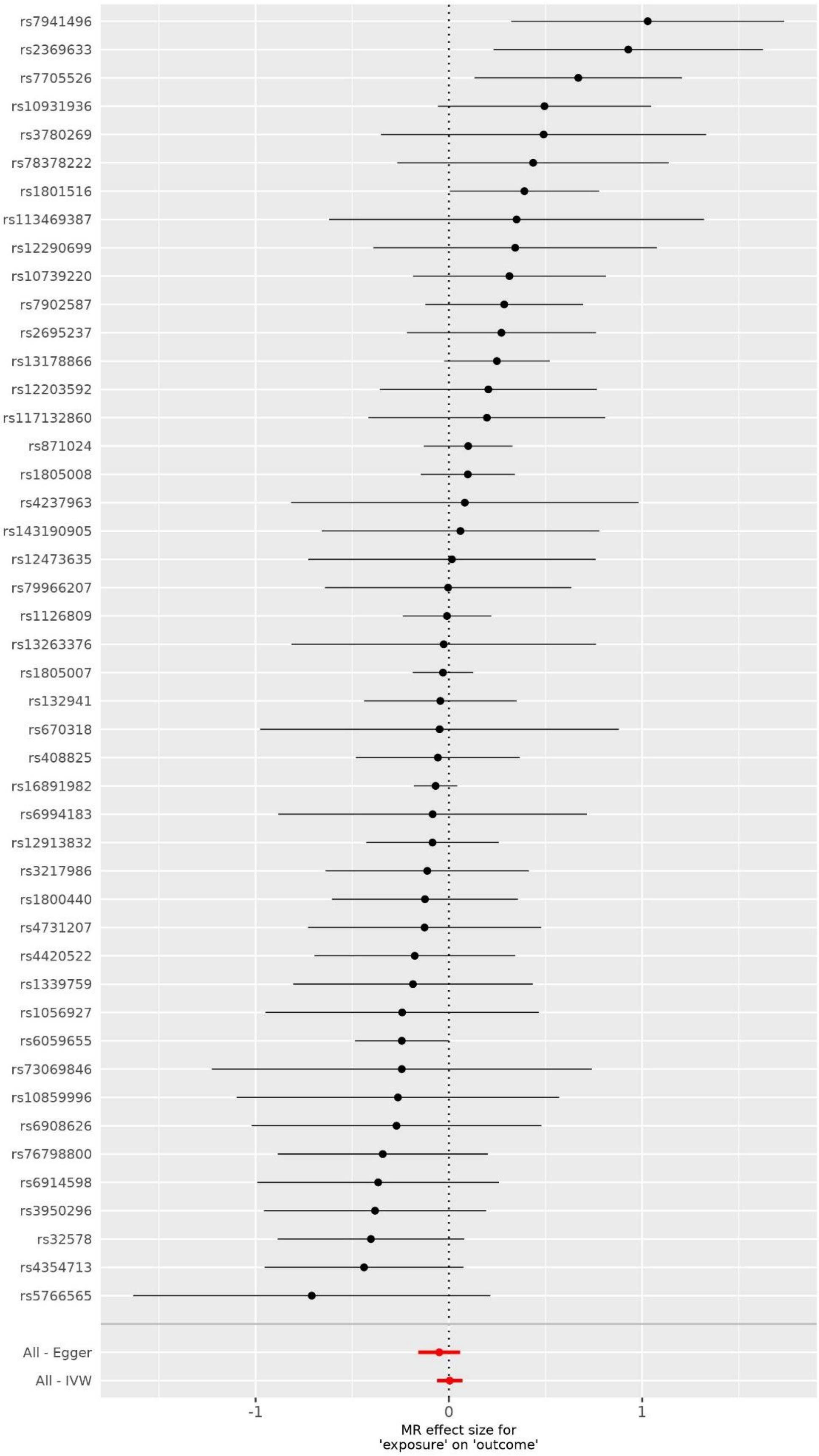

Non-glioblastoma glioma/glioma as exposure

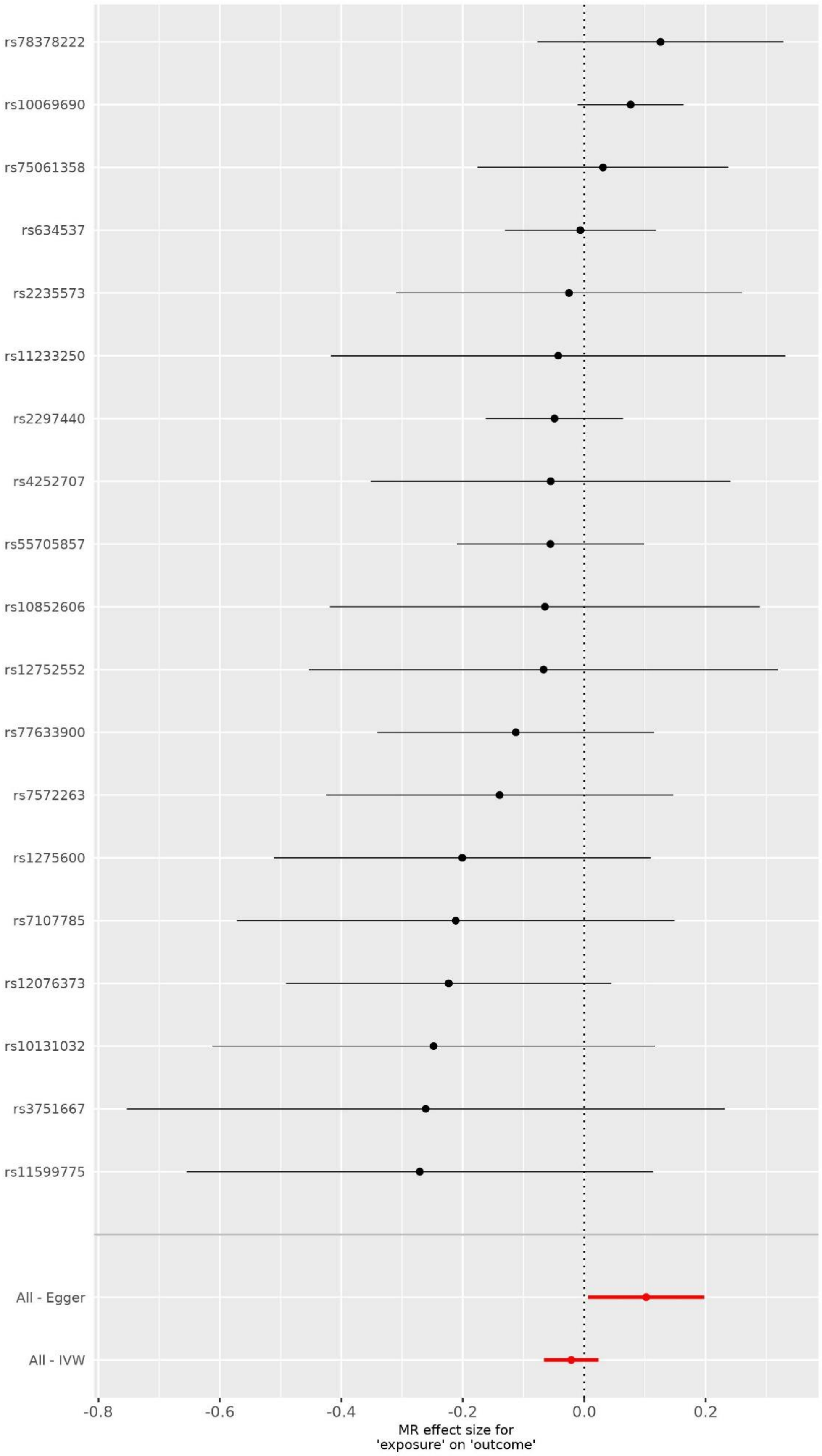

Oral cavity and pharyngeal cancer as exposure

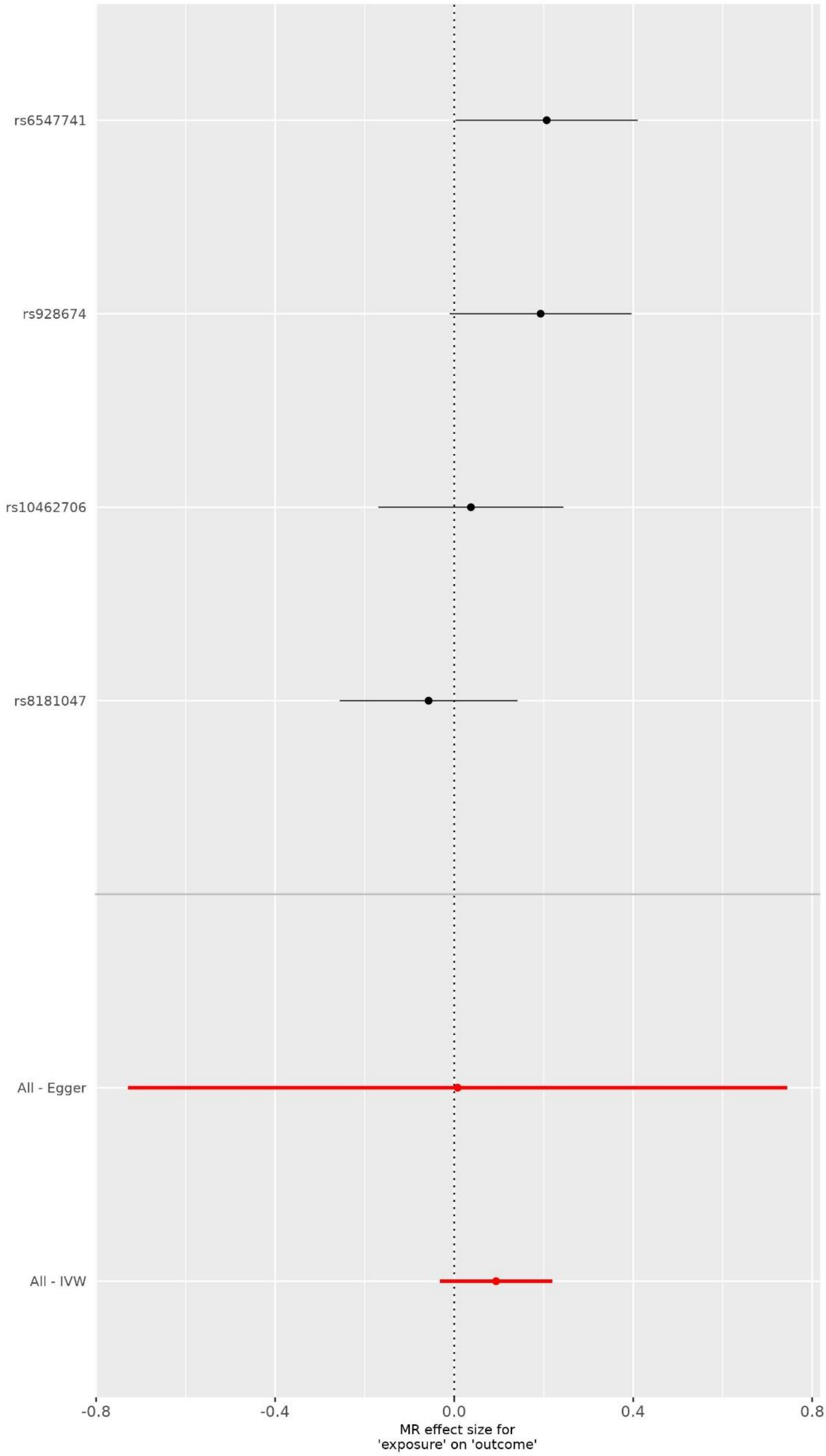

Pancreatic cancer

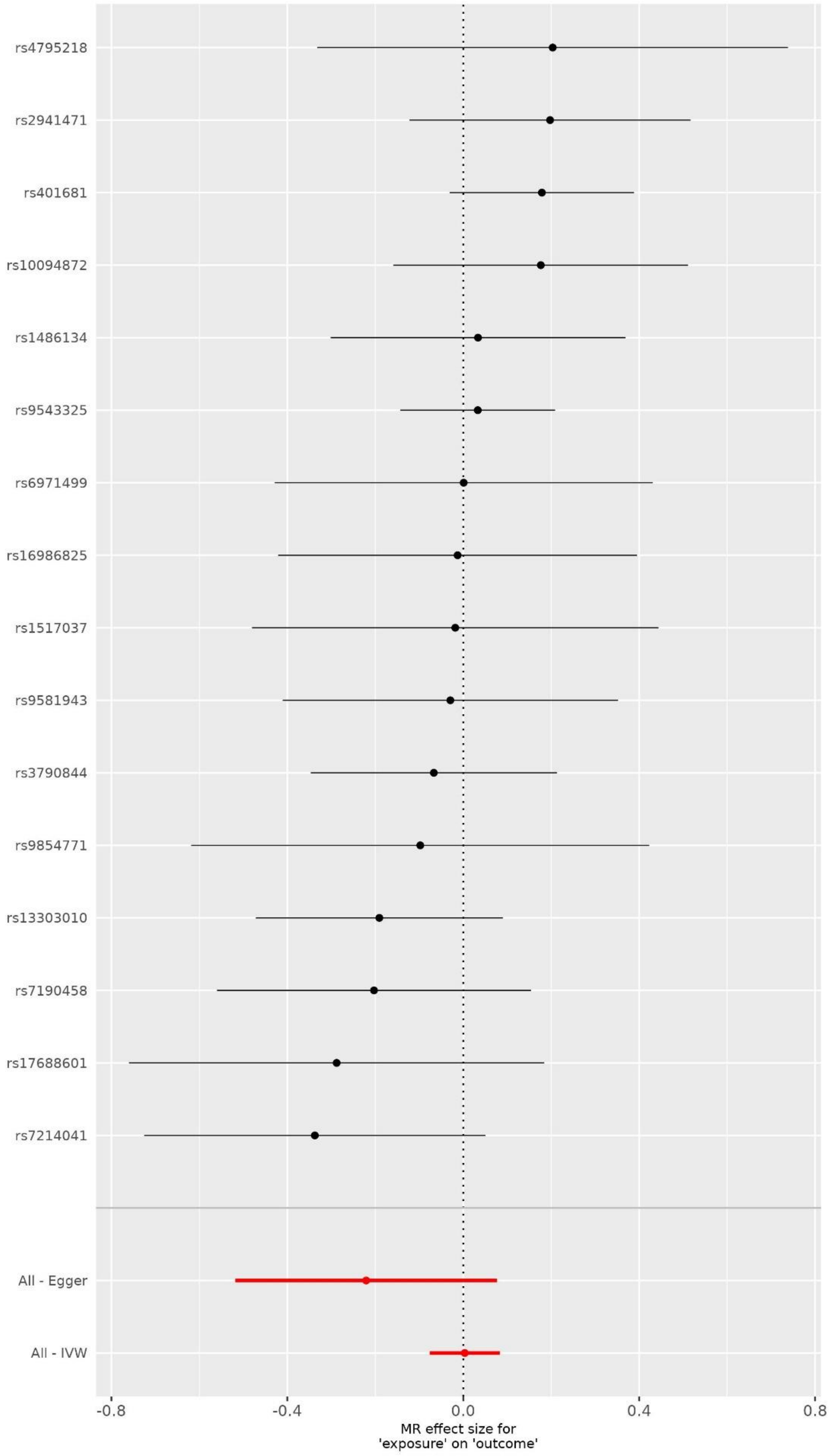

Prostate cancer

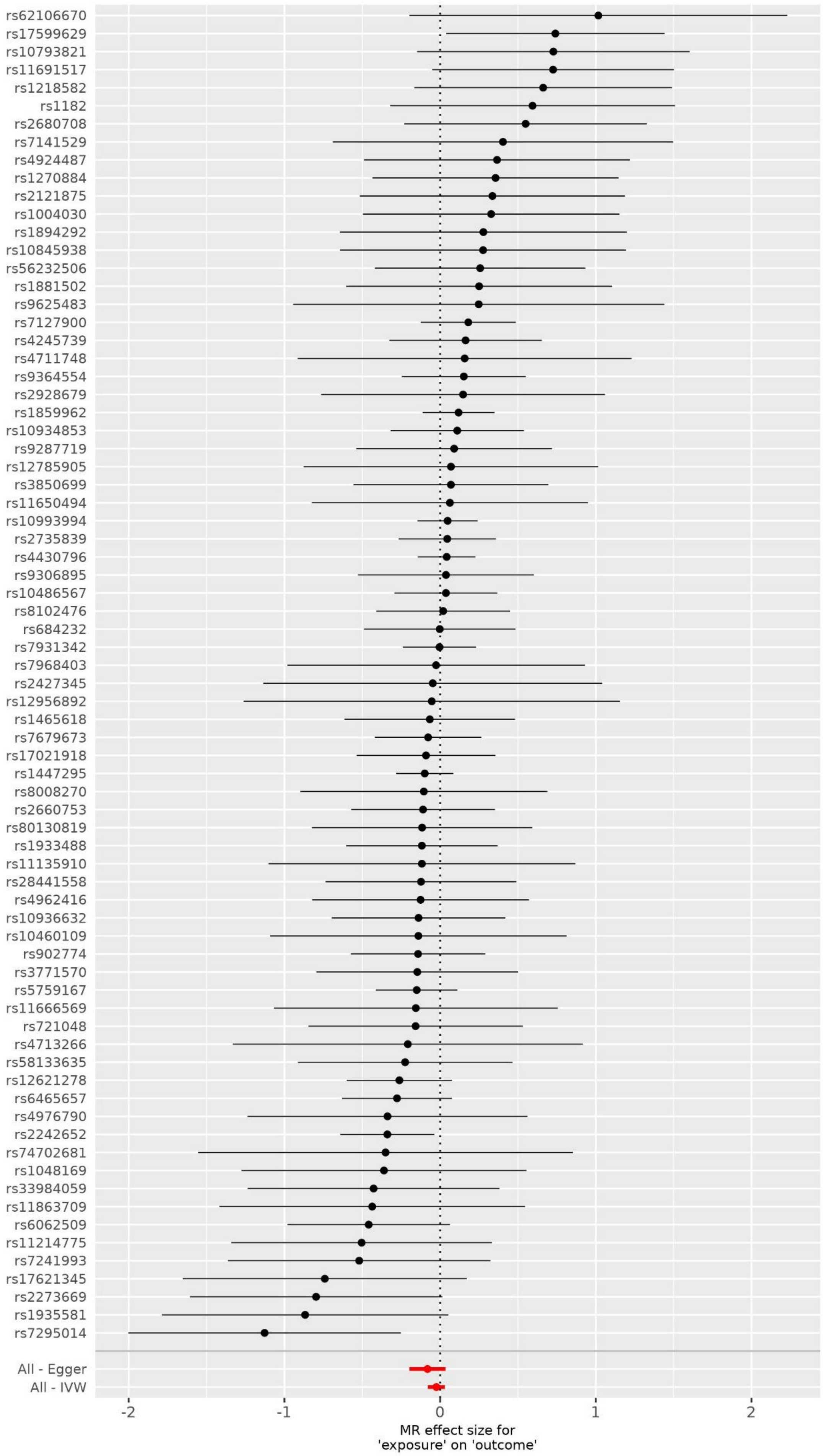

Renal cell carcinoma

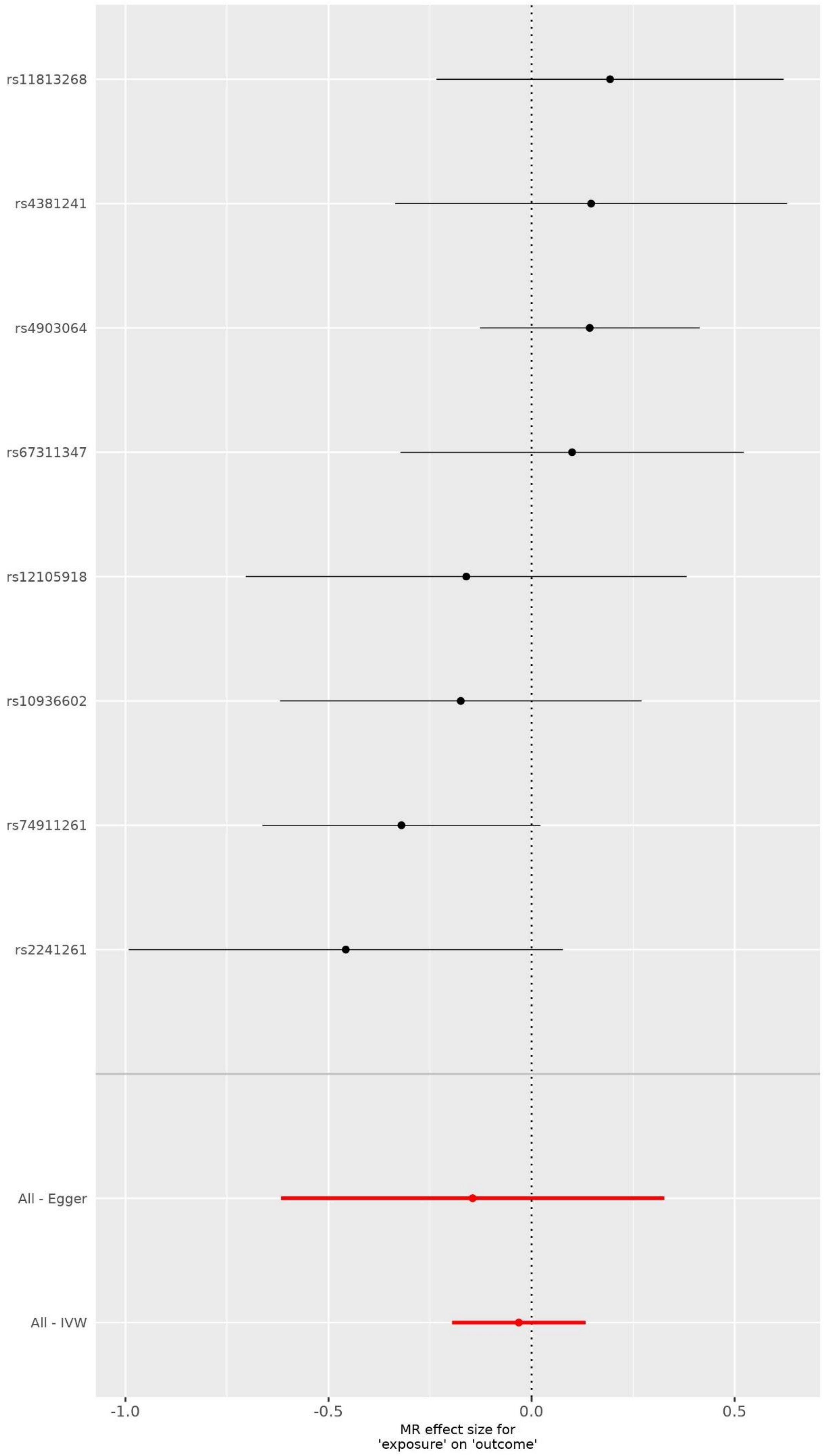

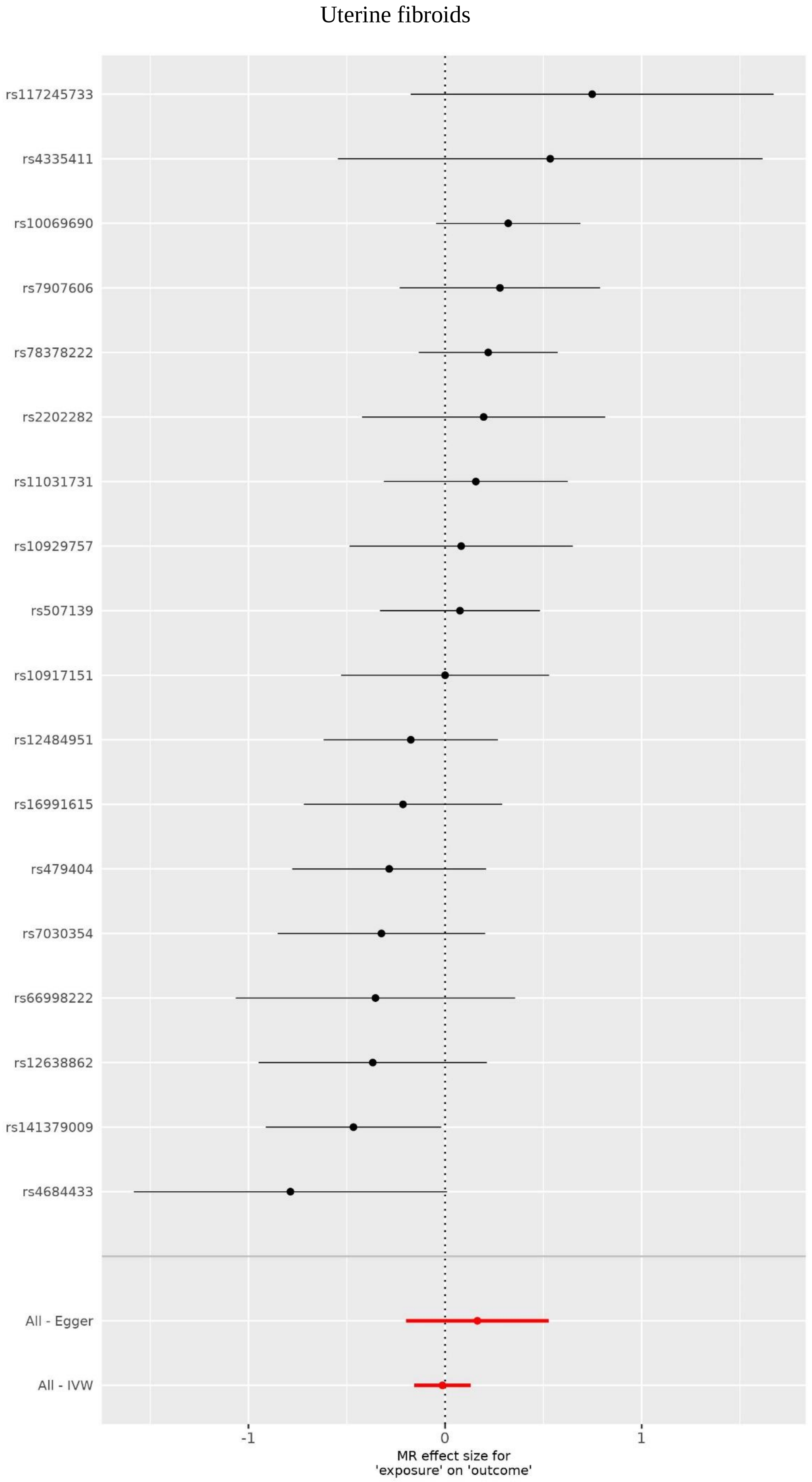
Uterine fibroids
rs117245733
rs4335411
rs10069690
rs7907606
rs78378222
rs2202282
rs11031731
rs10929757
rs507139
rs10917151
rs12484951
rs16991615
rs479404
rs7030354
rs66998222
rs12638862
rs141379009
rs4684433
All - Egger
All - IVW
-1
0
1
MR effect size for
'exposure' on 'outcome'

**Supplementary Figure 2. PD without UKBB. Plots showing point estimates of the exposures of interest; Exposure of interest at the top of each plot.**

A plot relating the effect sizes of the SNP-exposure association and the SNP-outcome associations with standard error bars. Lines correspond to causal estimates using each of the methods.

Breast cancer as exposure

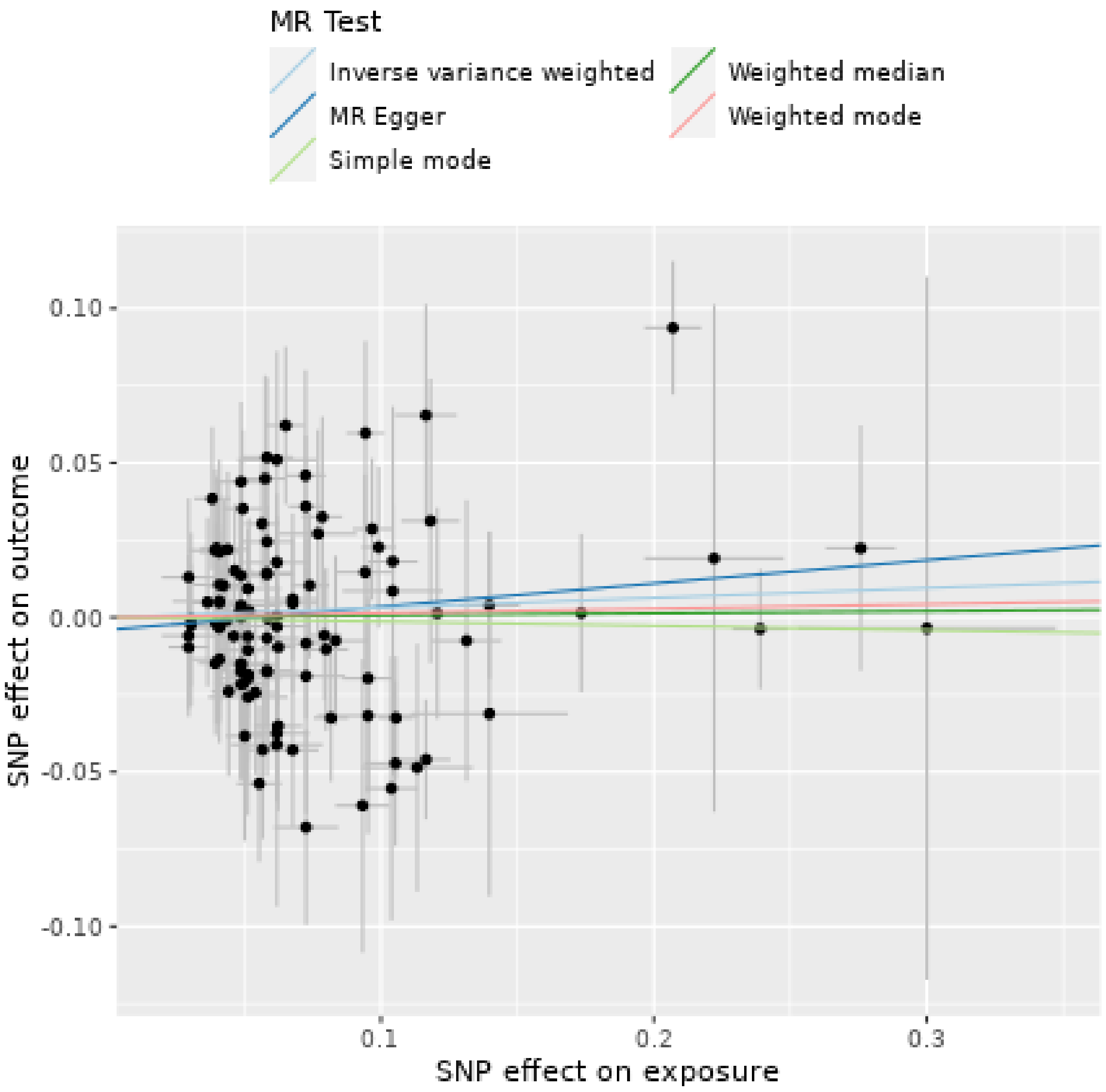

Chronic lymphocytic leukemia as exposure

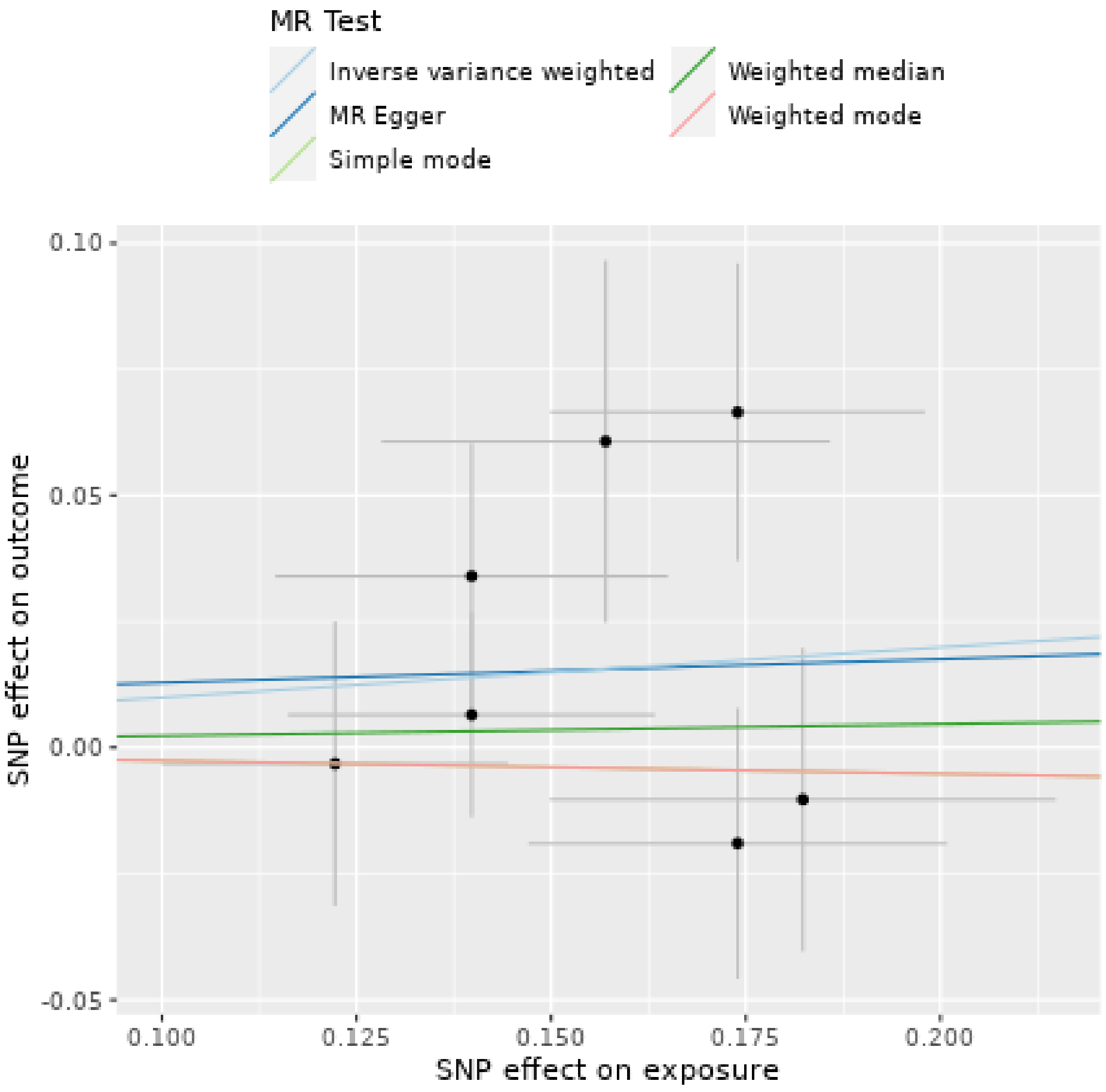

Colorectal cancer as exposure

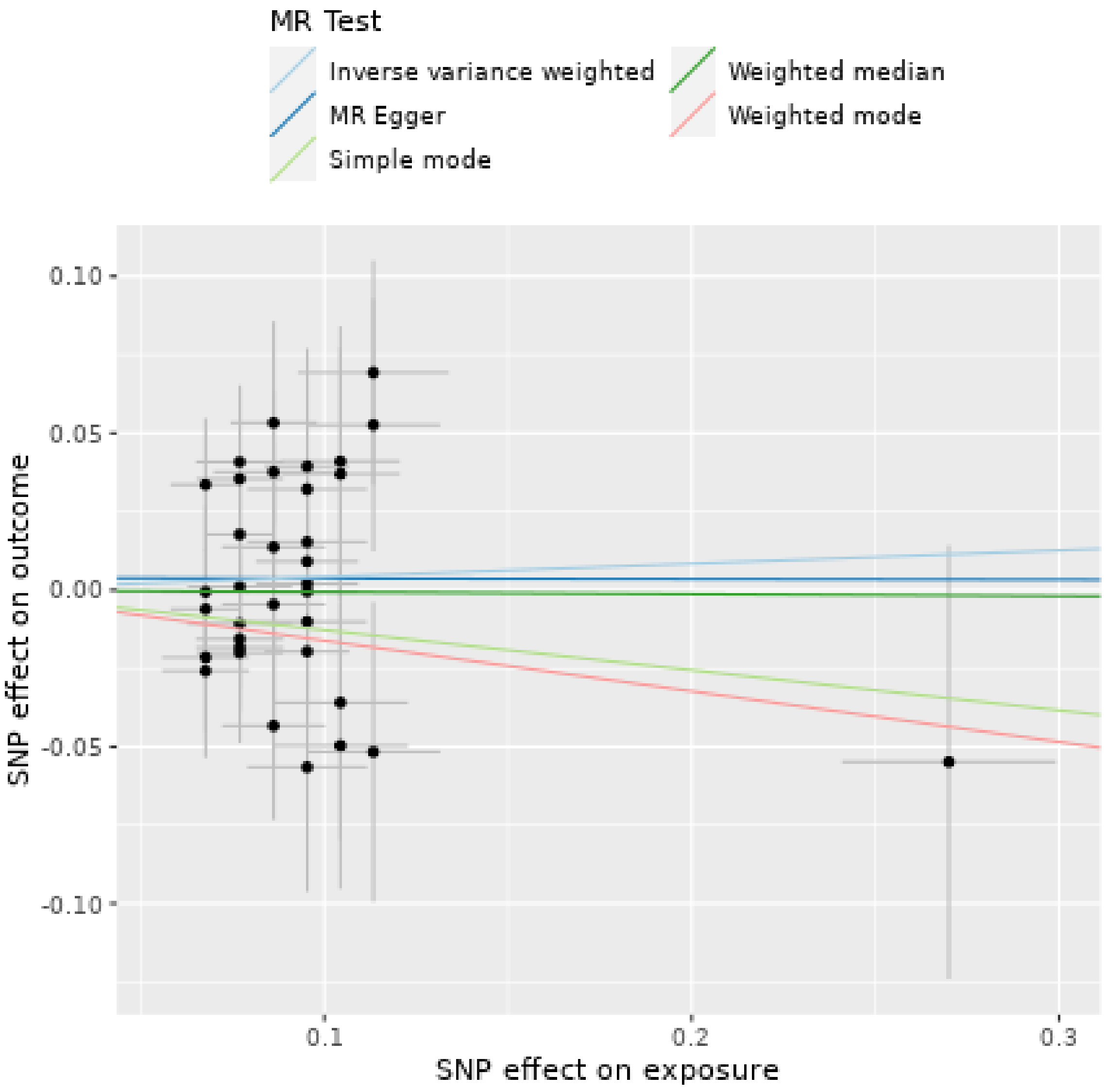

Cutaneous squamous cell carcinoma as exposure

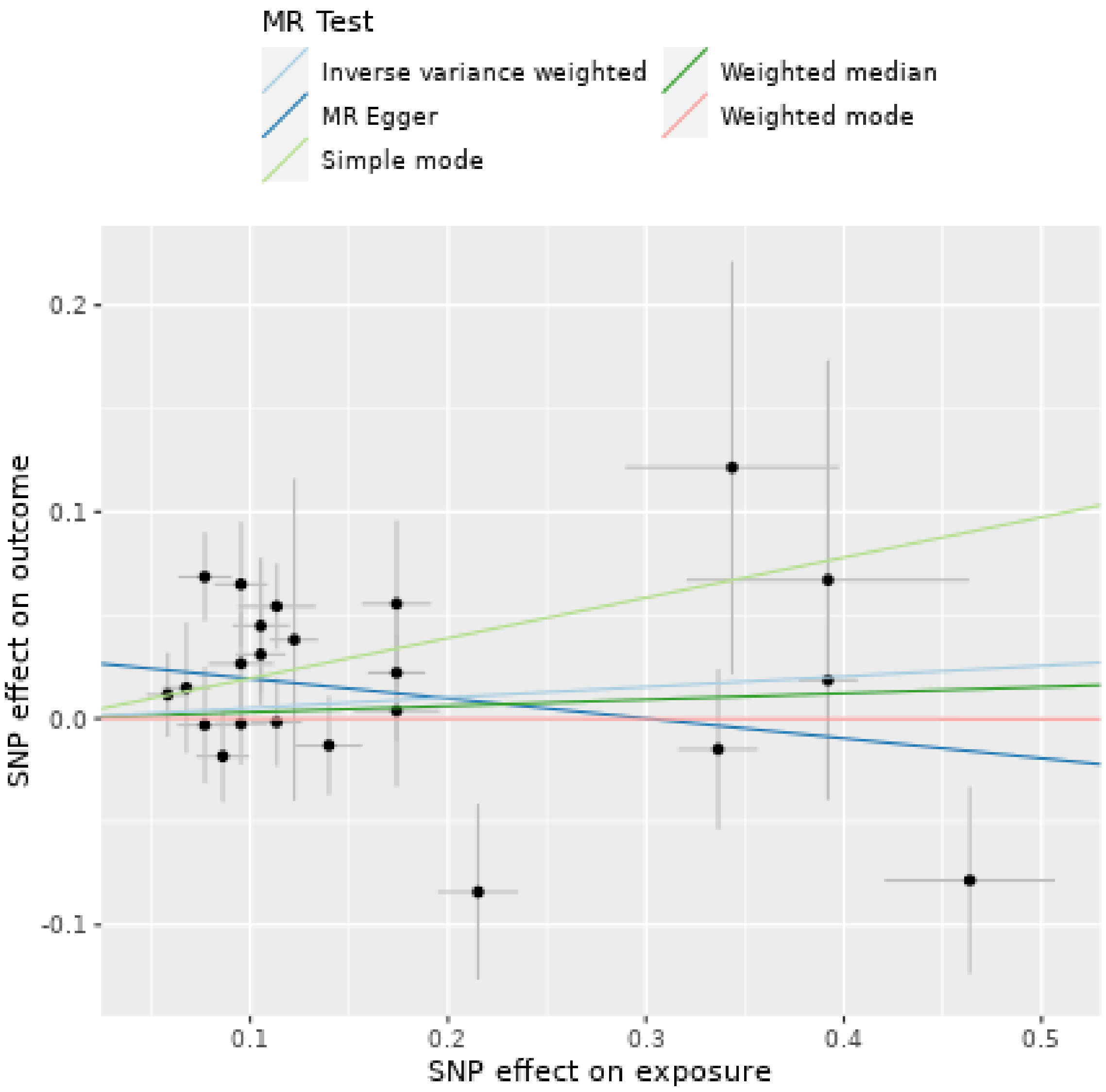

Combined analysis of keratinocyte cancers as exposure

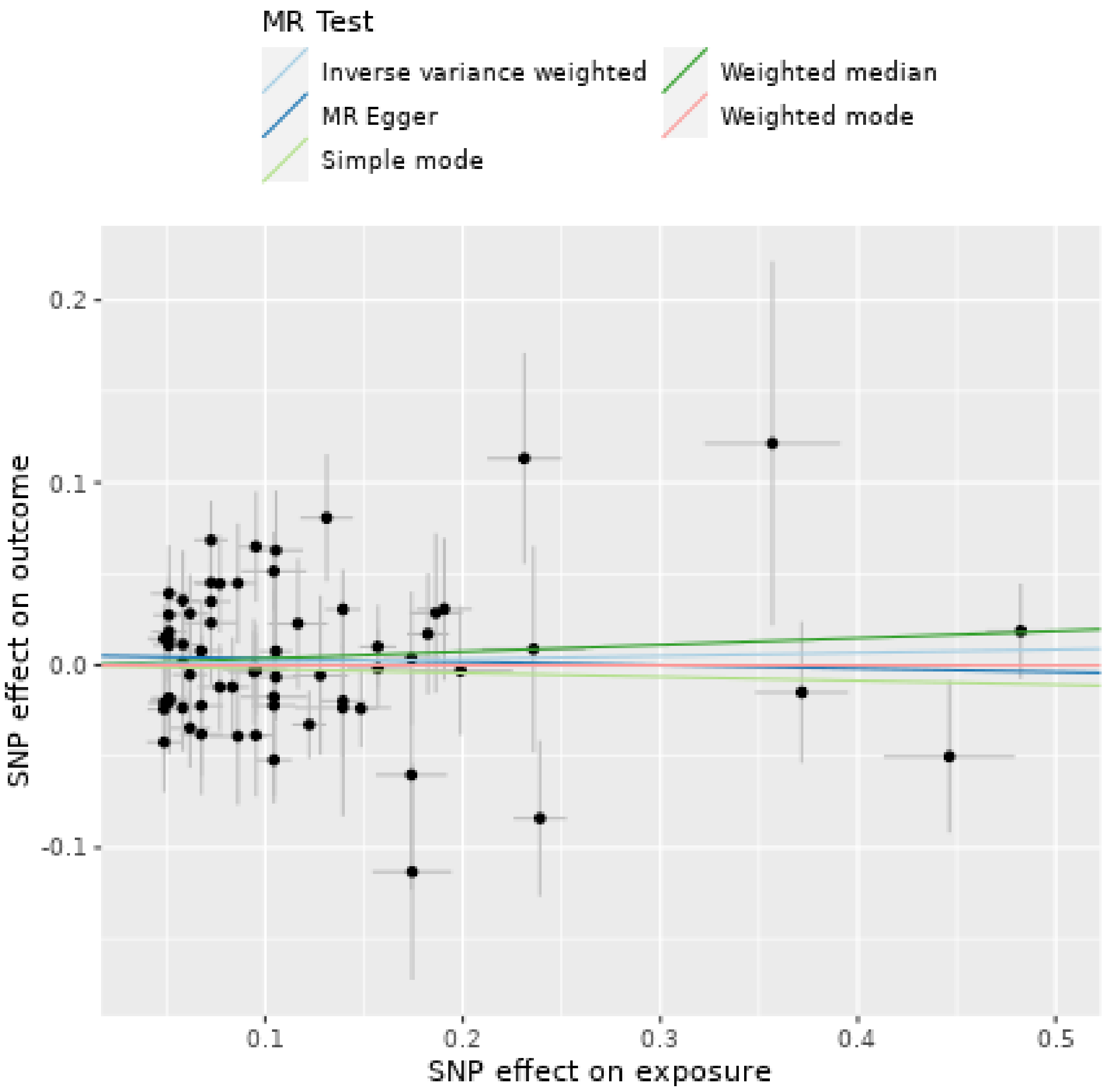

Endometrial cancer as exposure

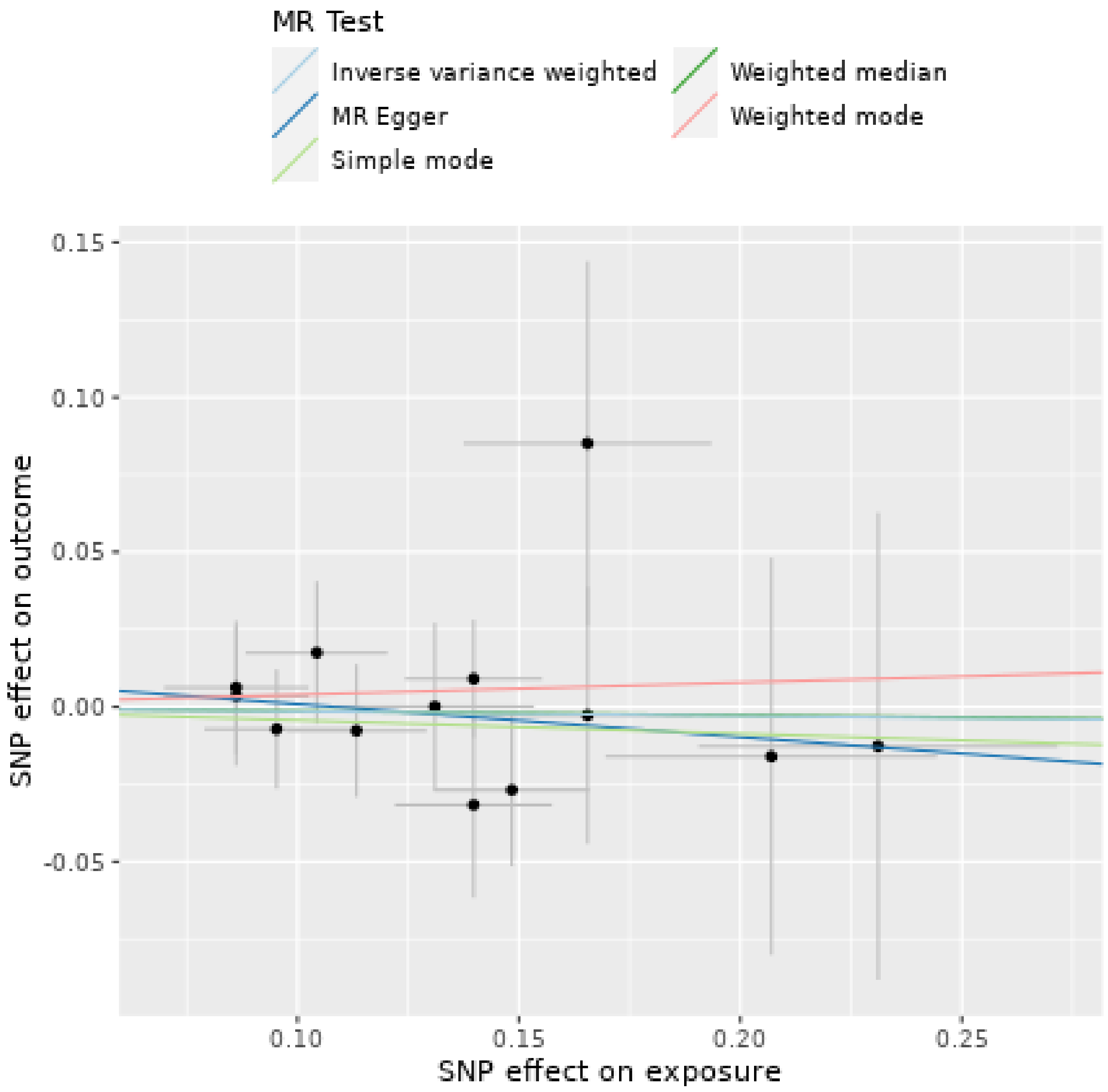

Lung cancer as exposure

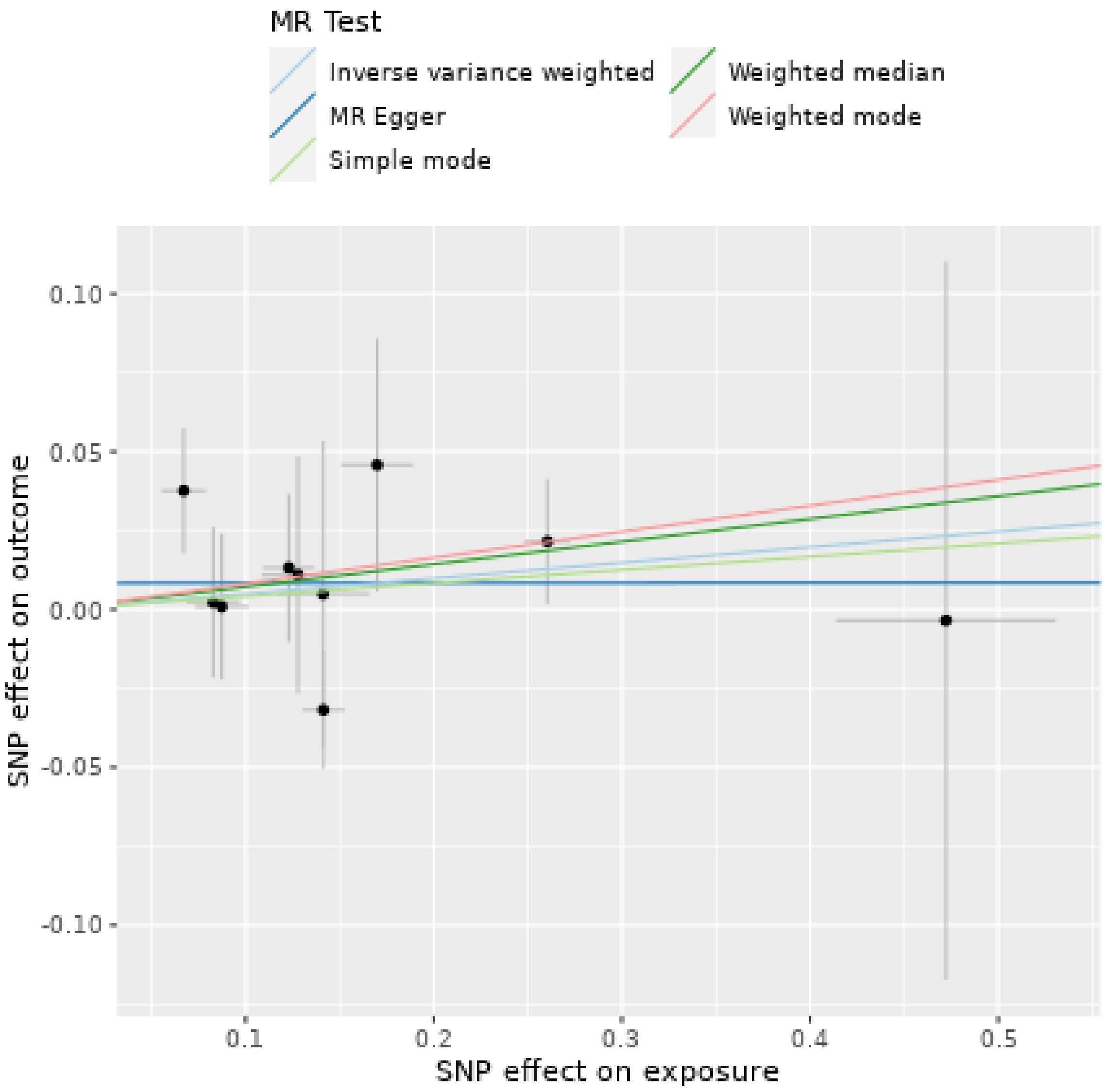

Lymphoma as exposure

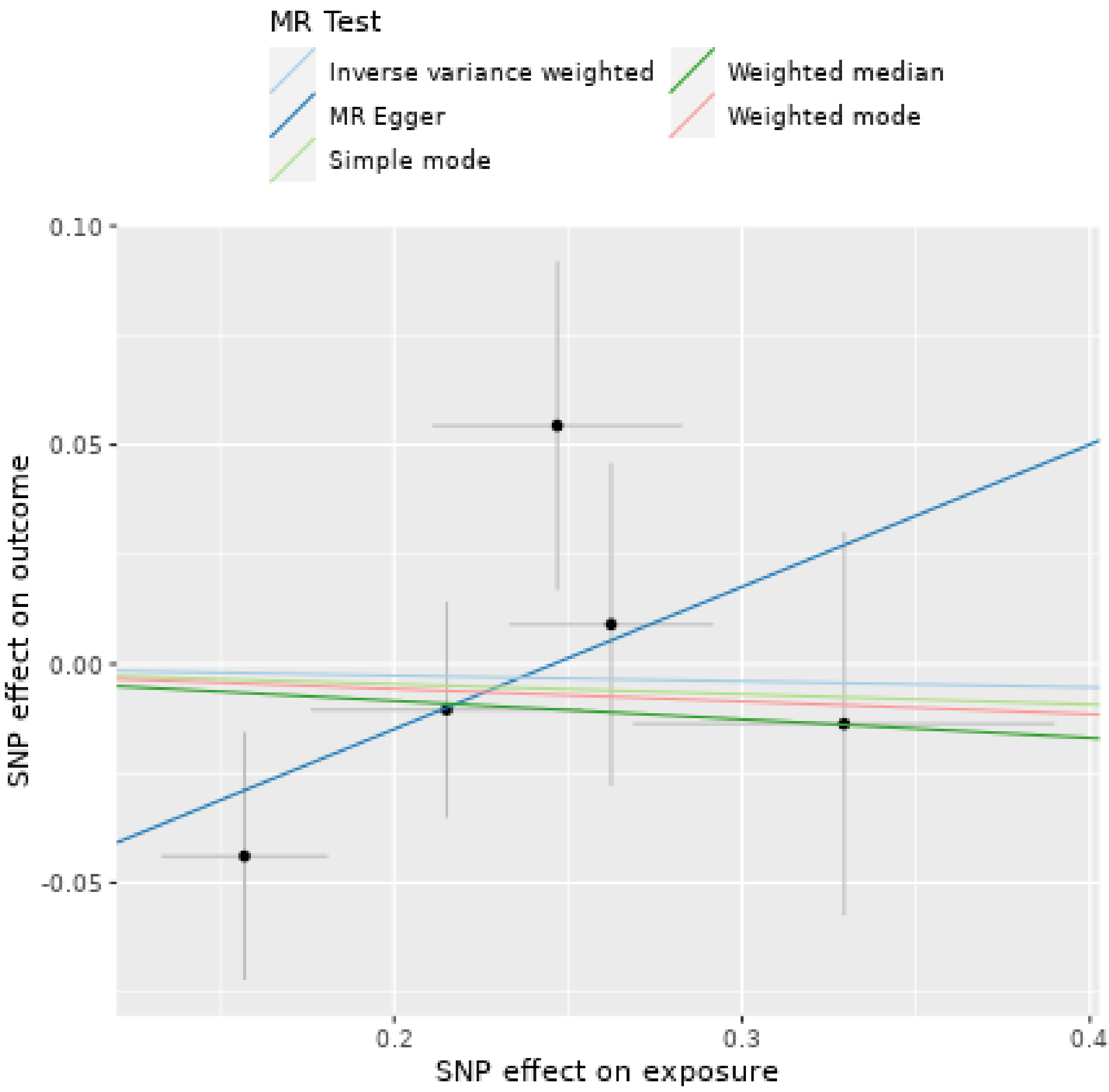

Melanoma as exposure

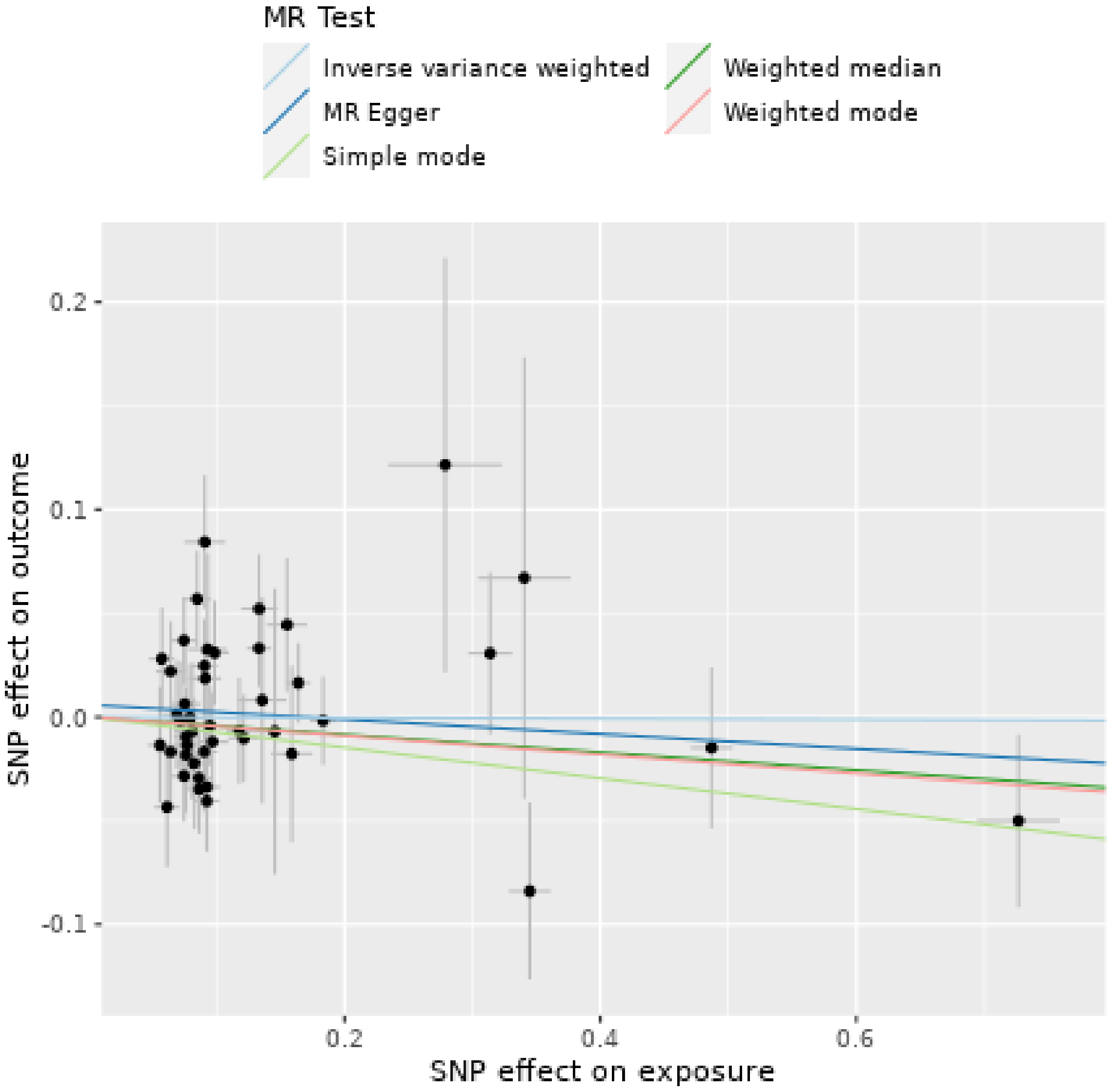

Non-glioblastoma glioma/Glioma as exposure

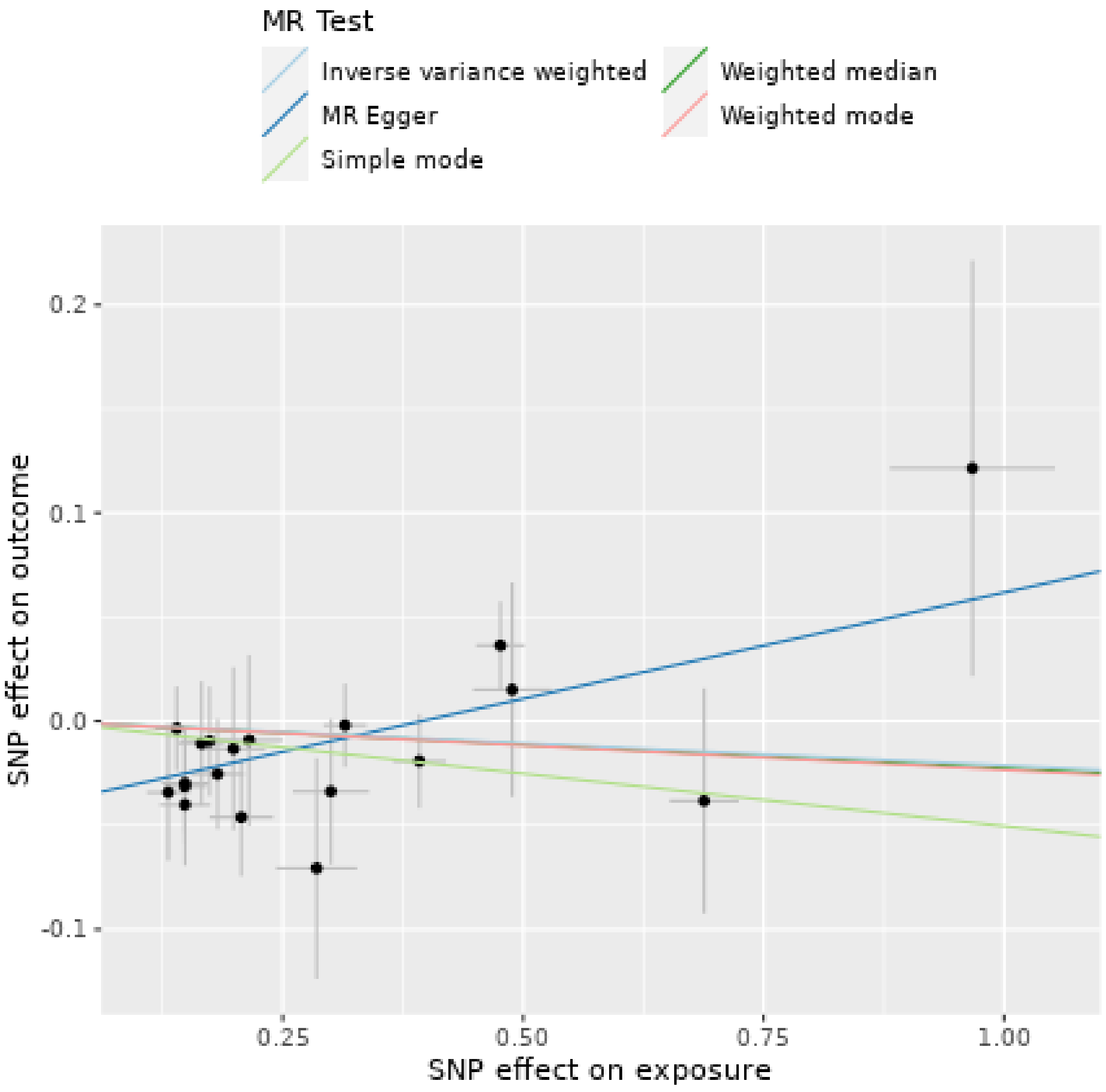

Oral cavity and pharyngeal cancer as exposure

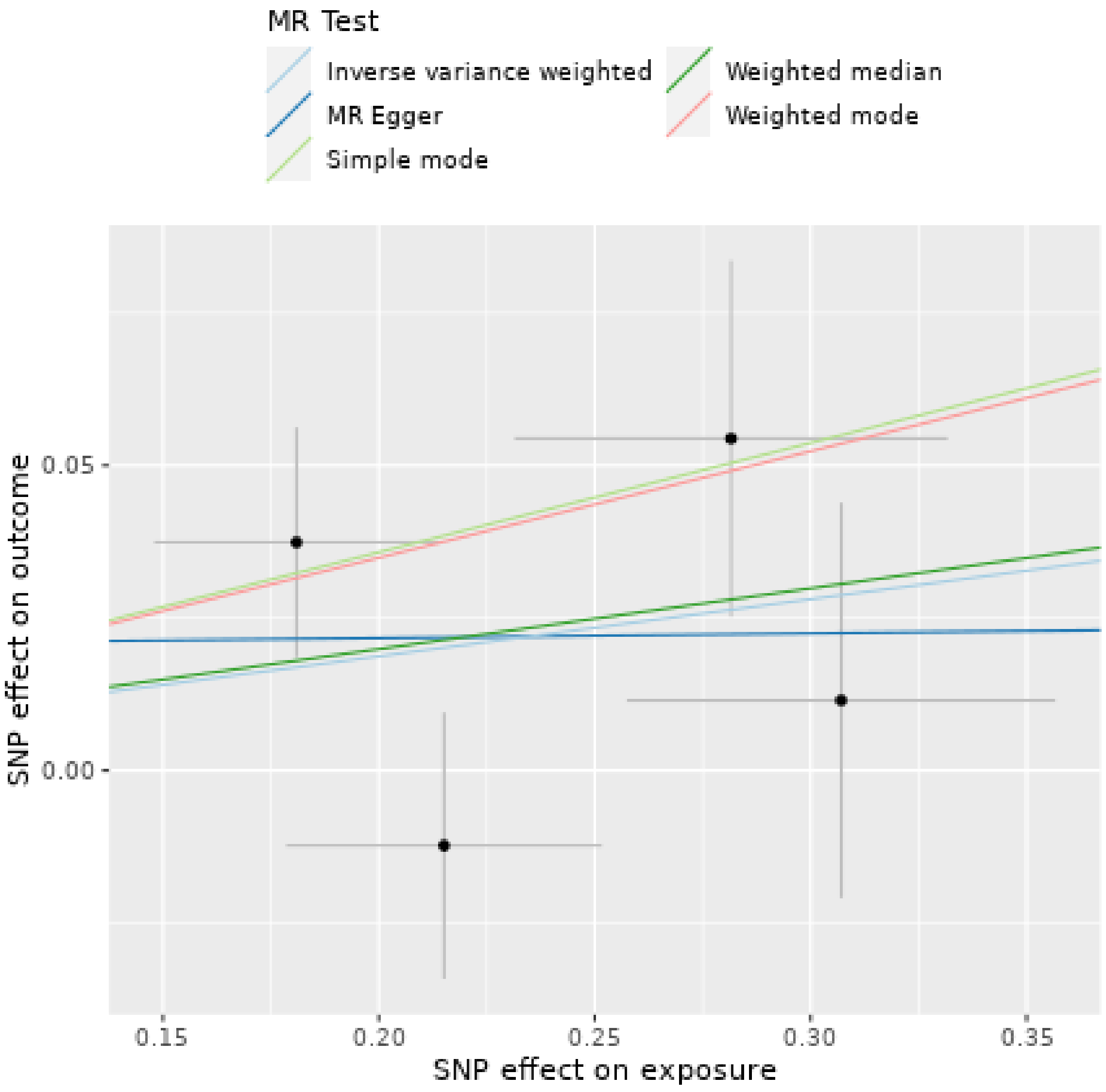

Pancreatic cancer as exposure

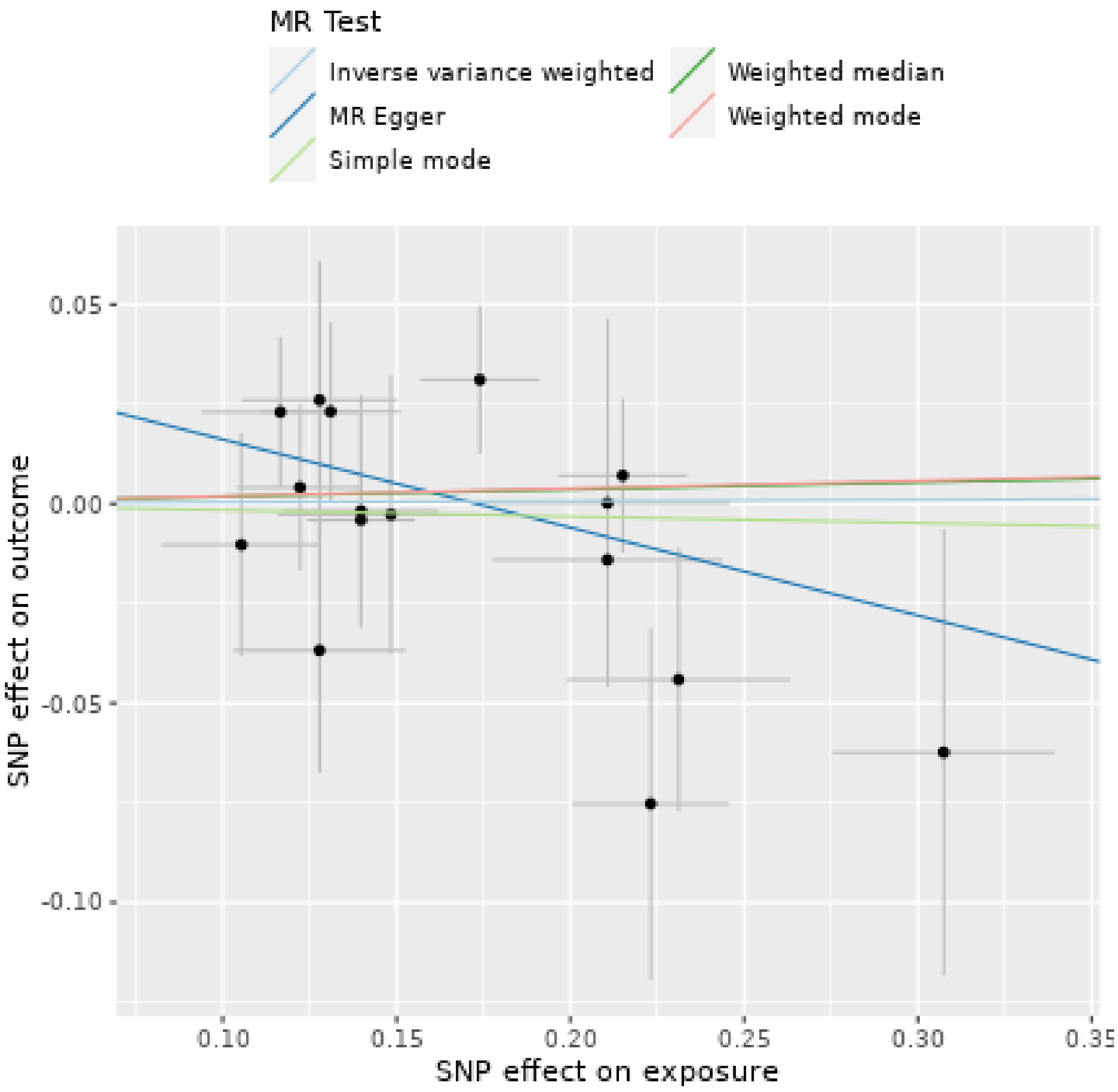

Prostate cancer as exposure

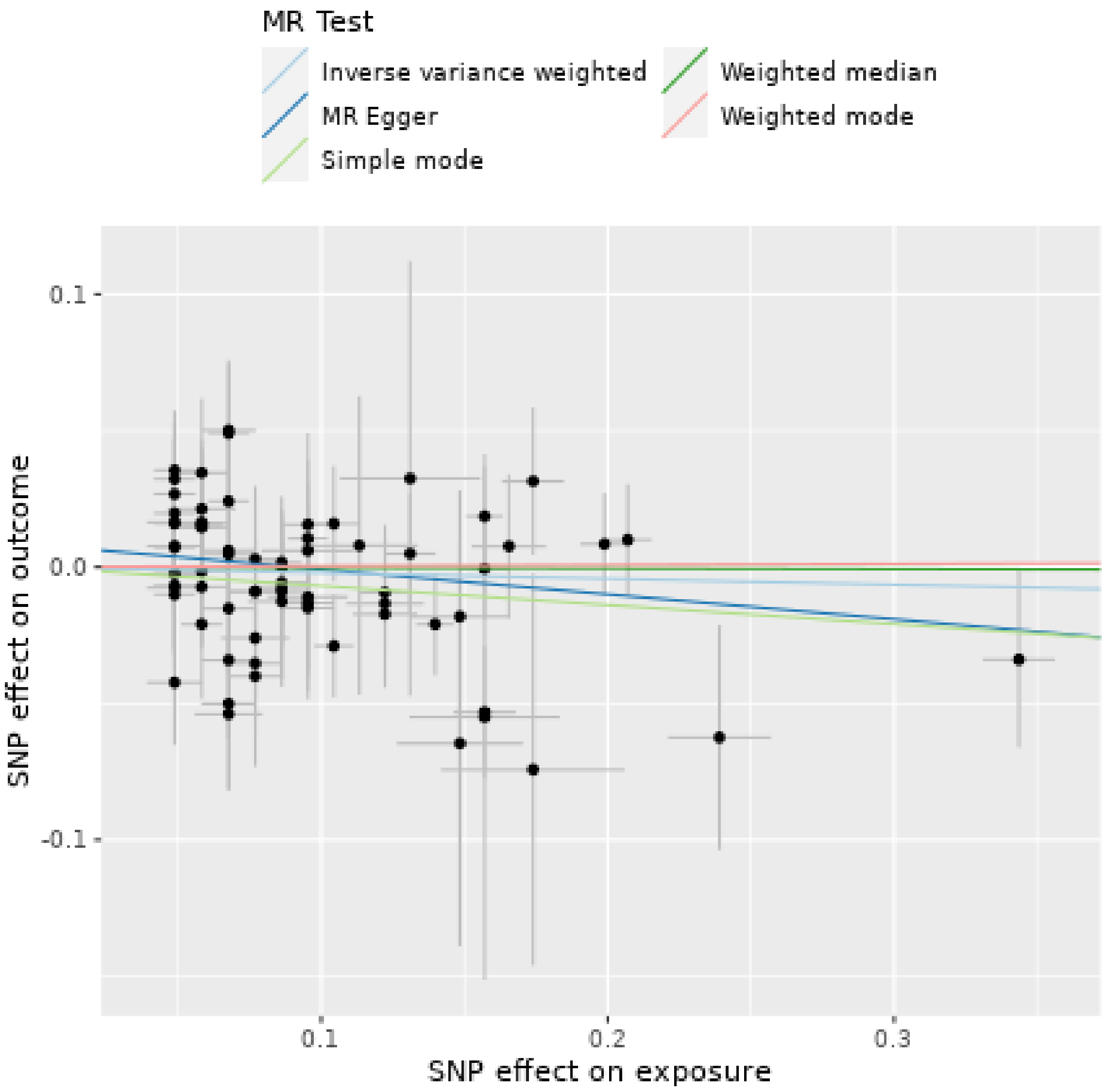

Renal cell carcinoma as exposure

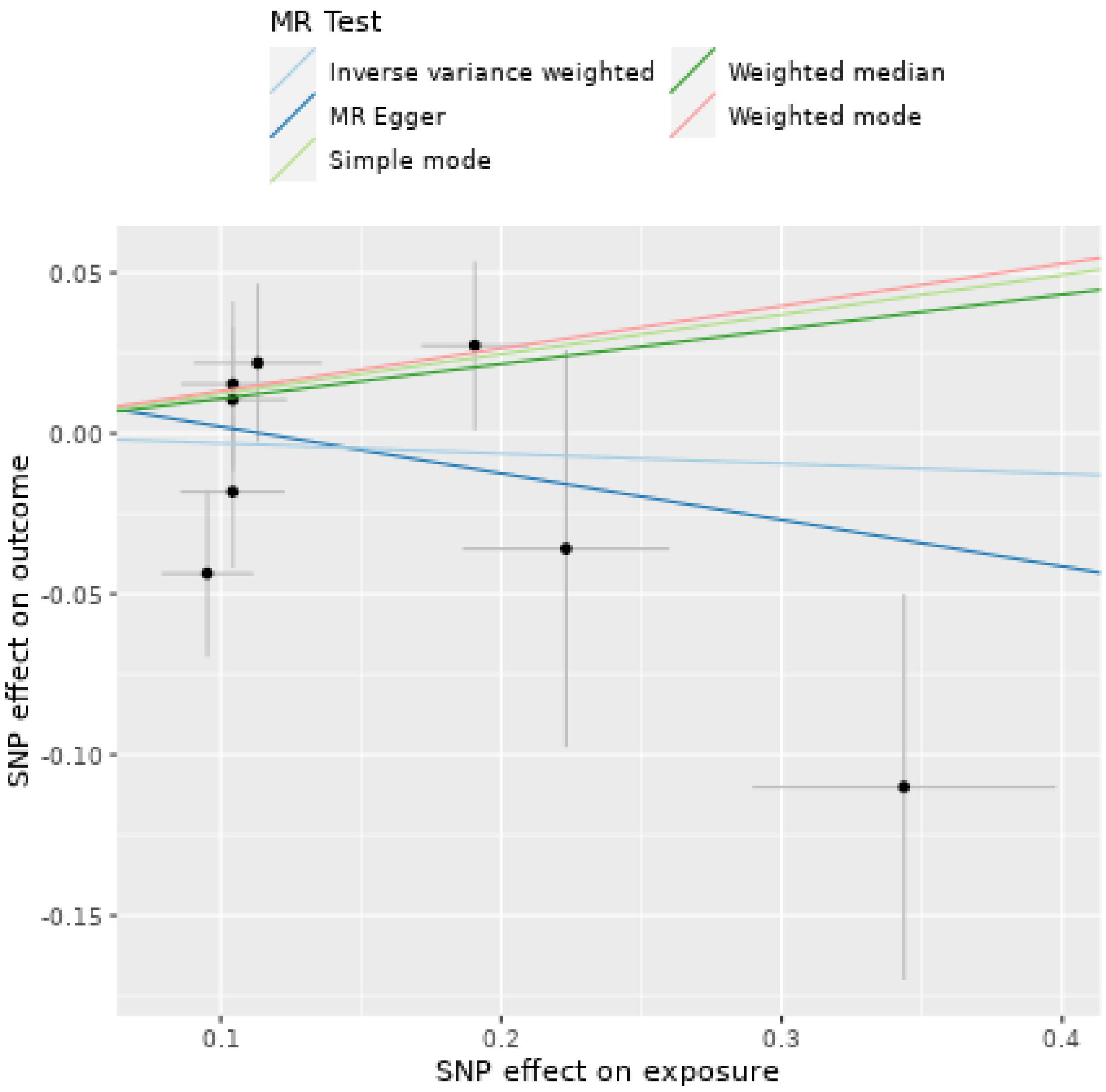

Uterine fibroids as exposure

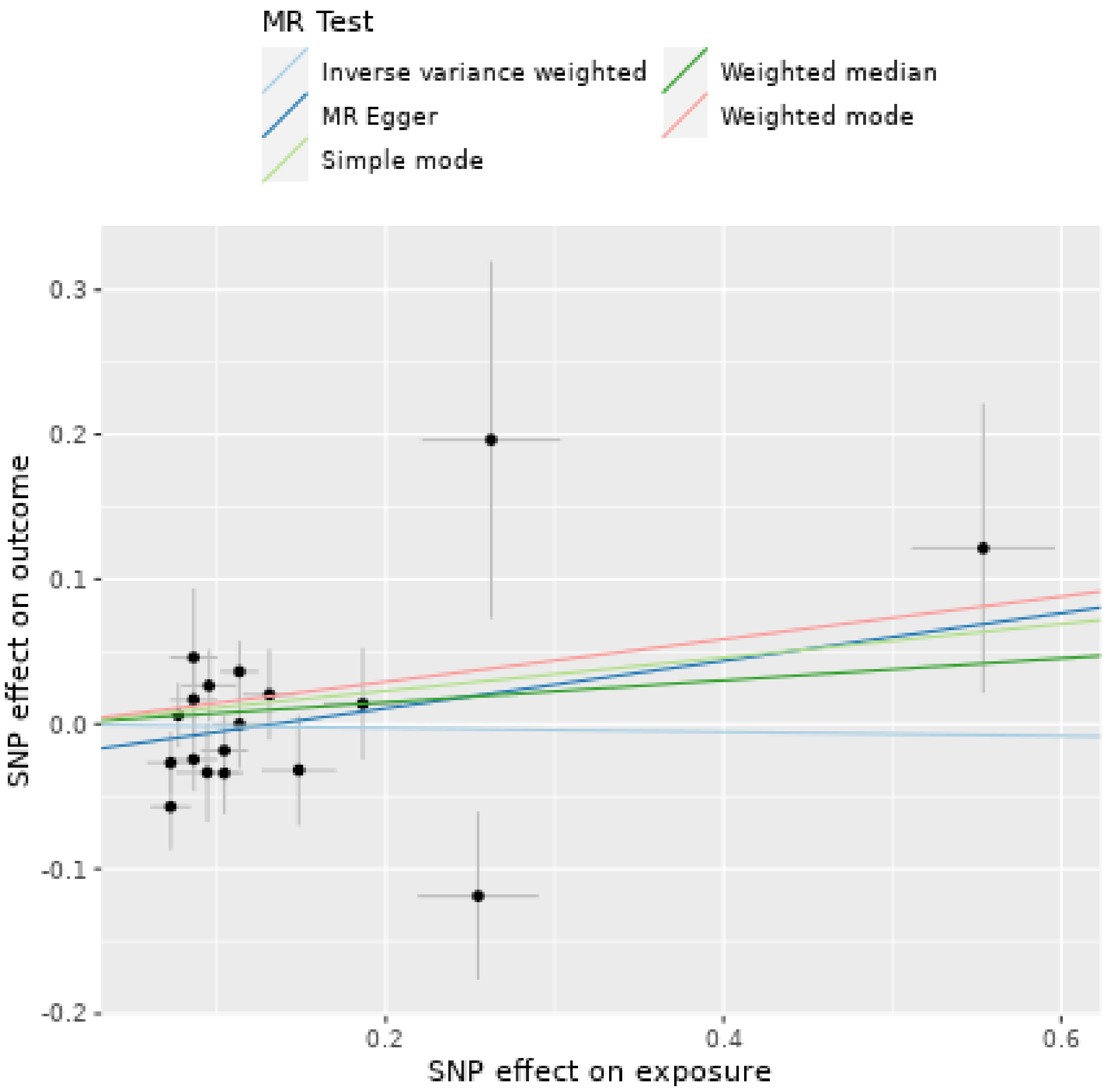

**Supplementary Figure 3. PD without UKBB. Funnel plots evaluated the presence of possible heterogeneity across the estimates. Exposure of interest at the top of each plot.**
Each SNPs represented by dots. Inverse variance weighted and MR Egger method averaged causal effect of all SNPs.

Breast cancer as exposure

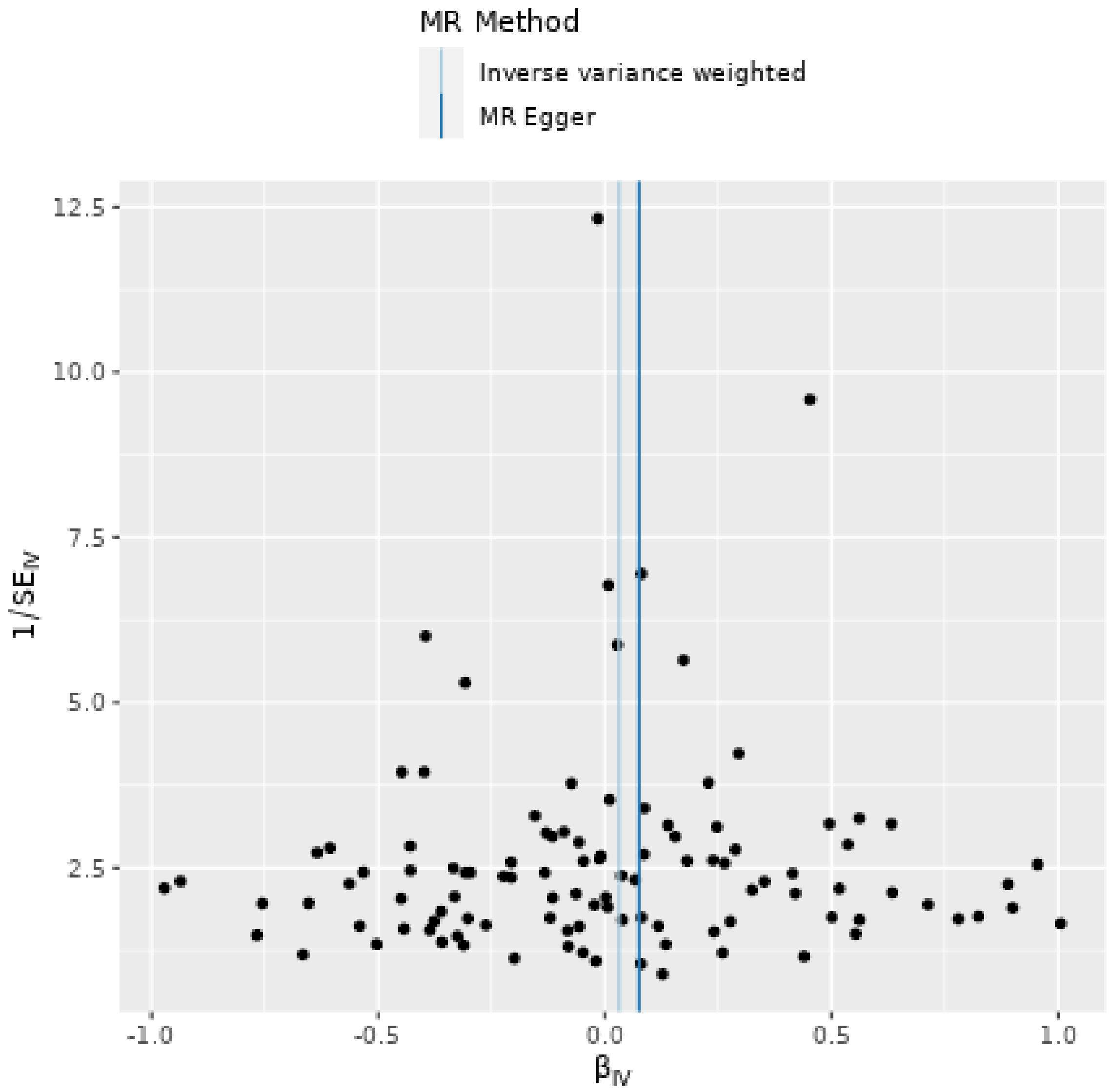

Chronic lymphocytic leukemia as exposure

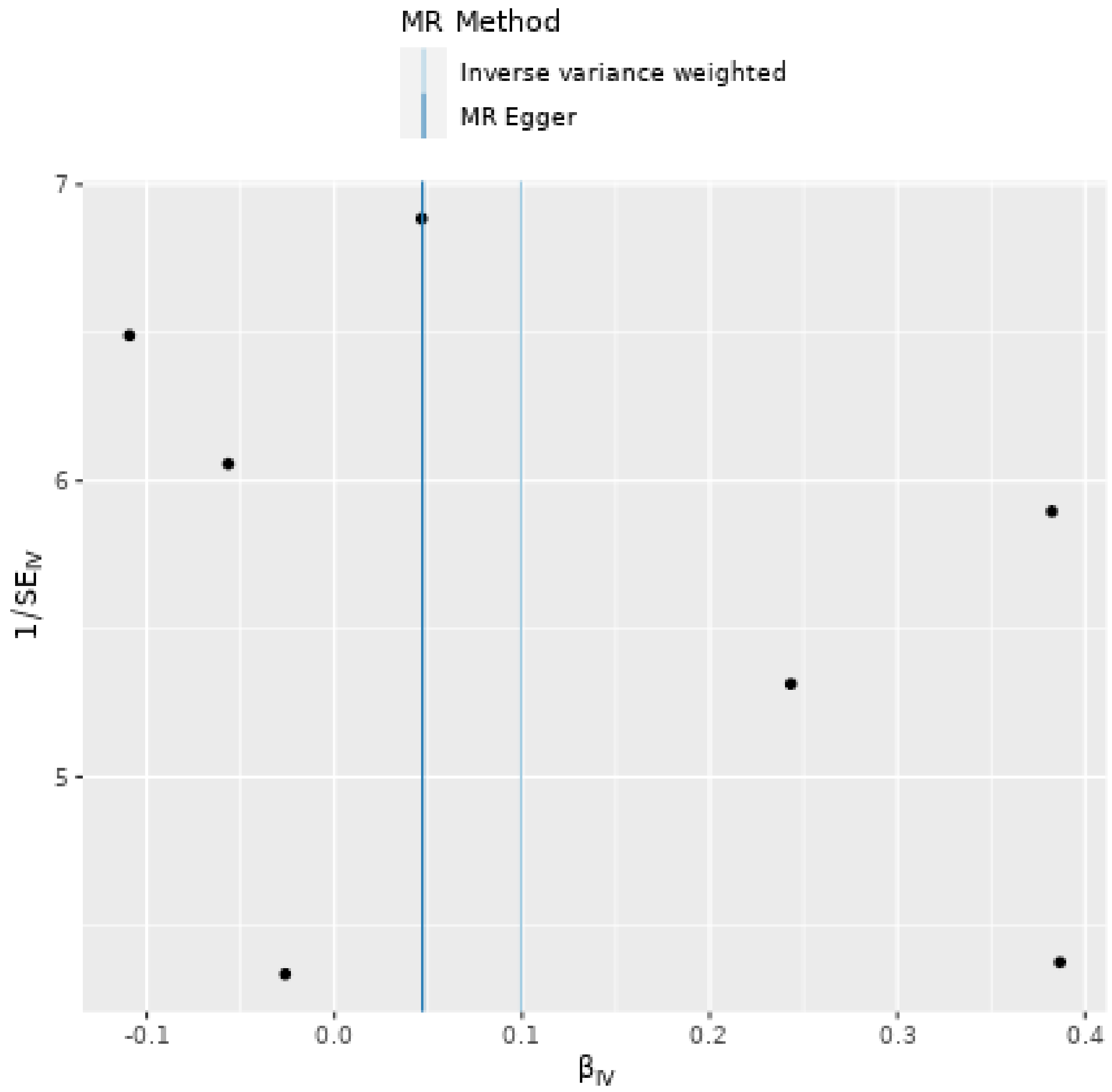

Colorectal cancer as exposure

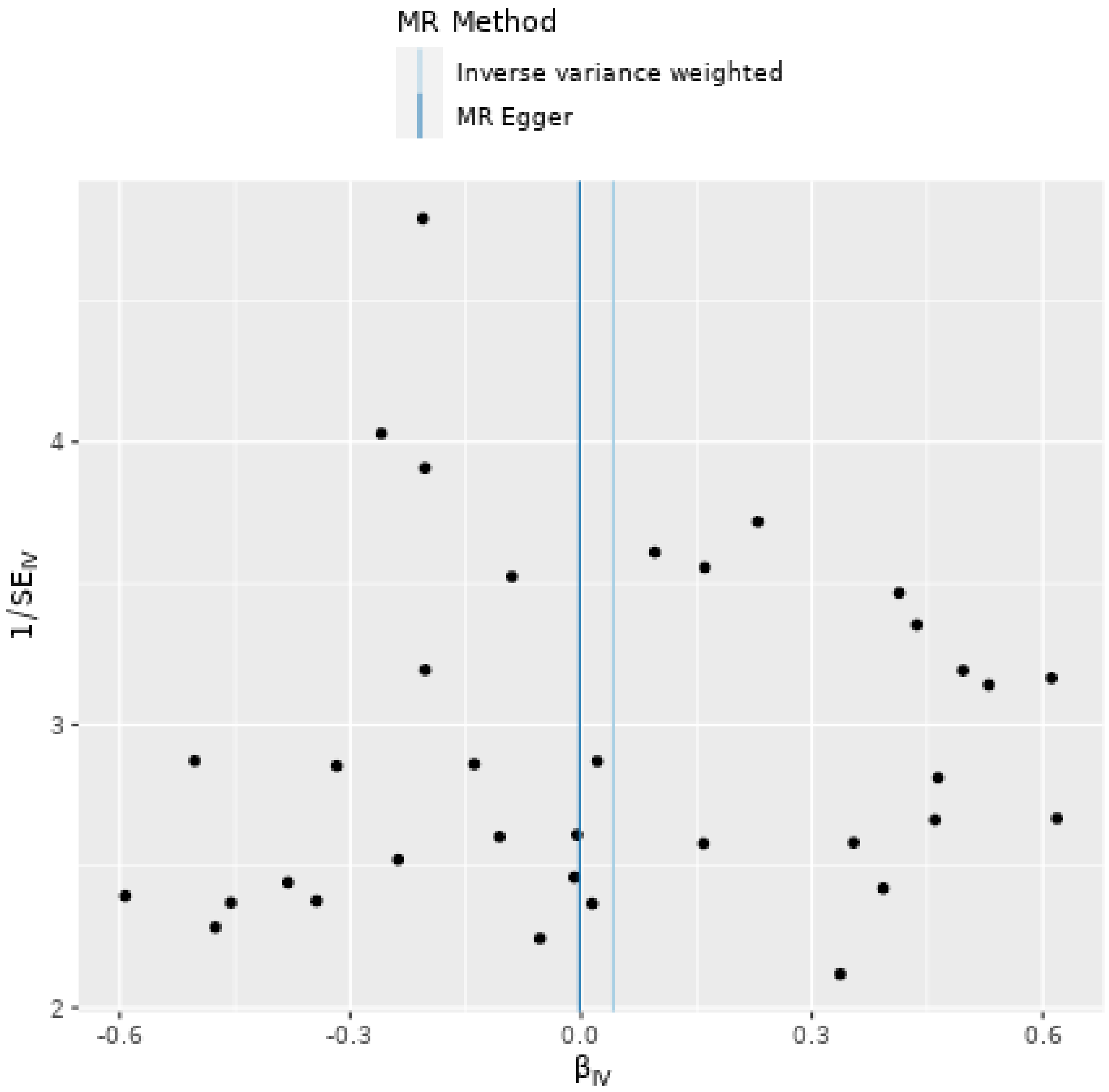

Cutaneous squamous cell carcinoma as exposure

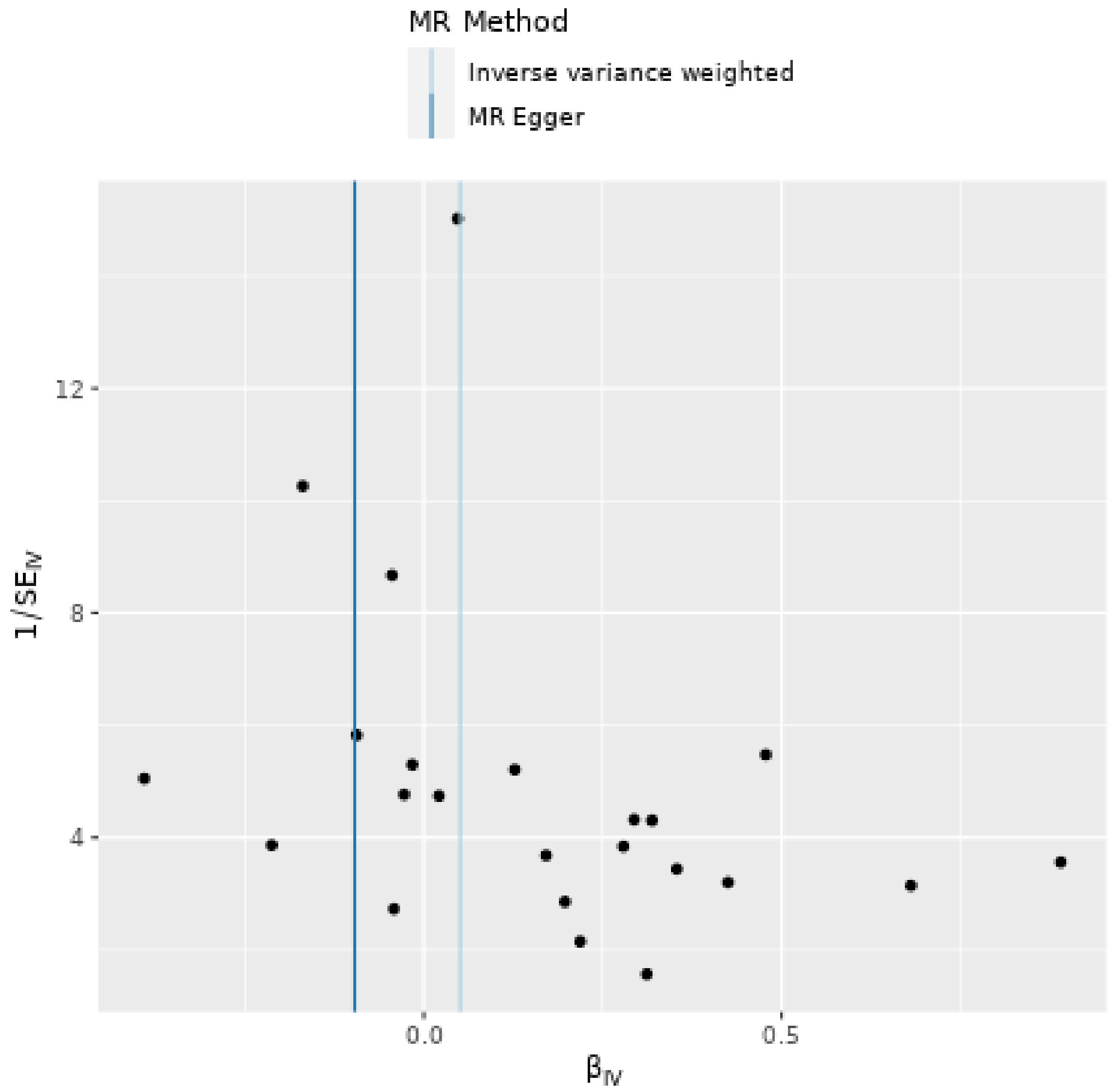

Combined analysis of keratinocyte cancers as exposure

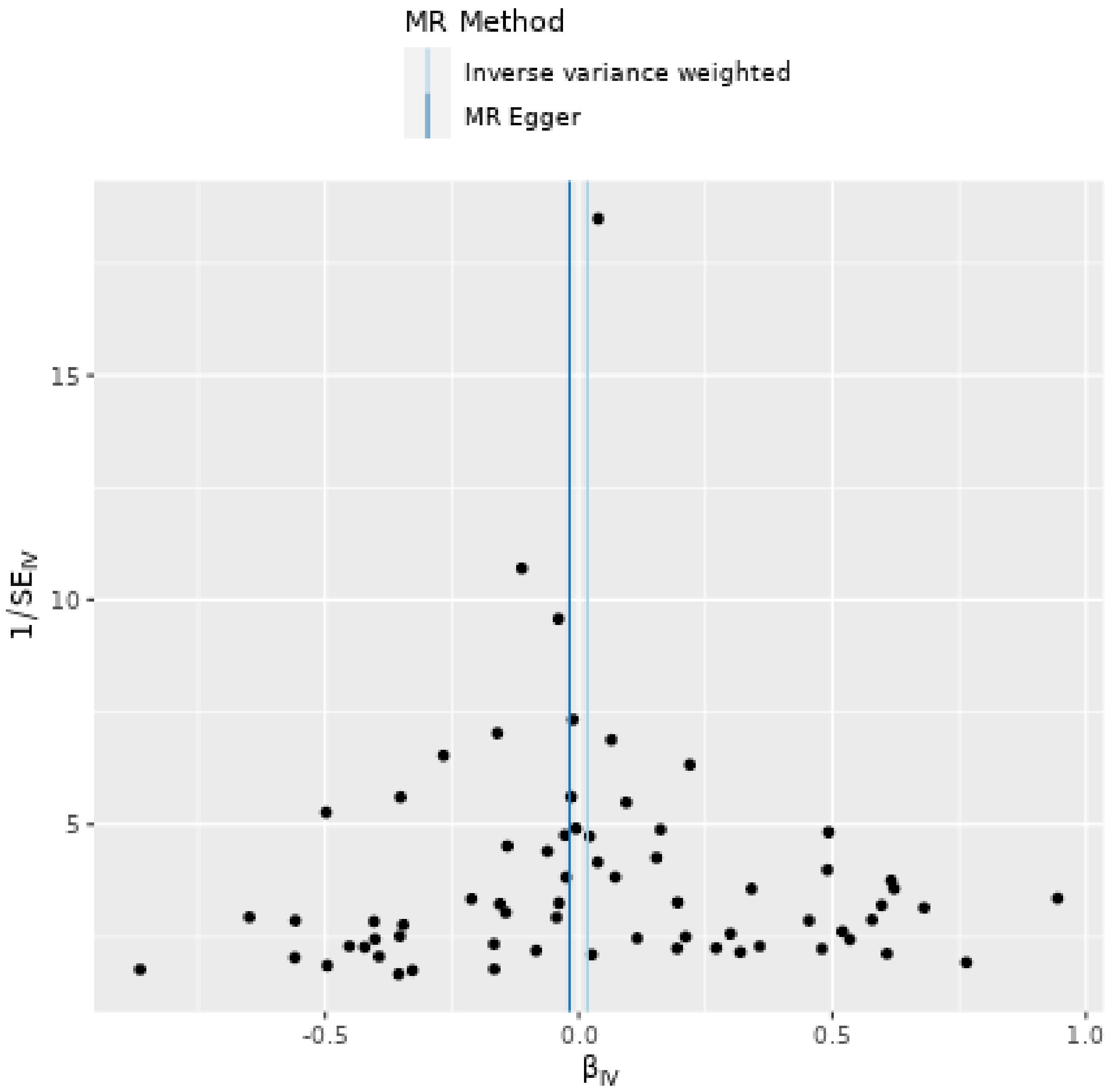

Endometrial cancer as exposure

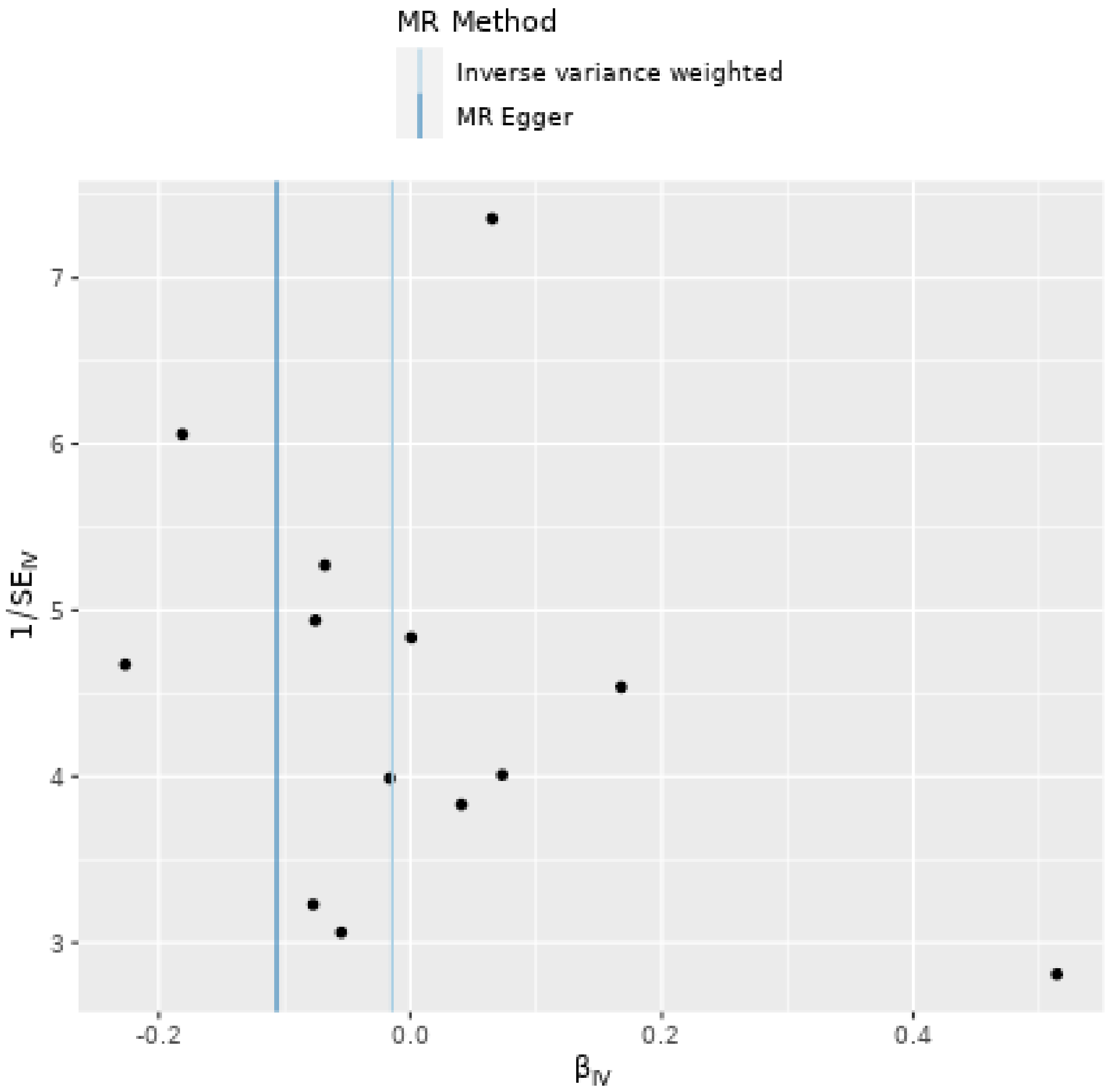

Lung cancer as exposure

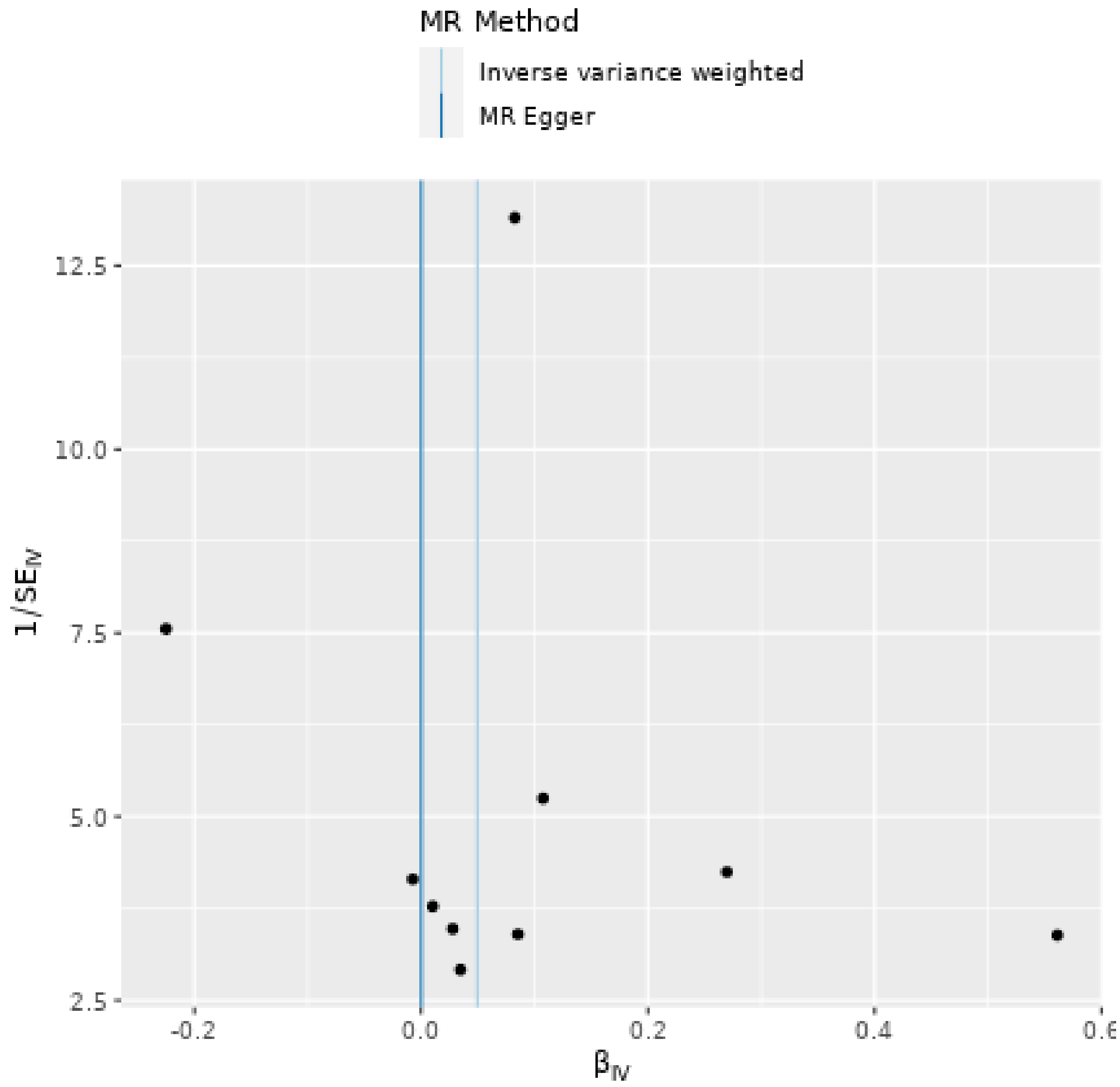

Lymphoma as exposure

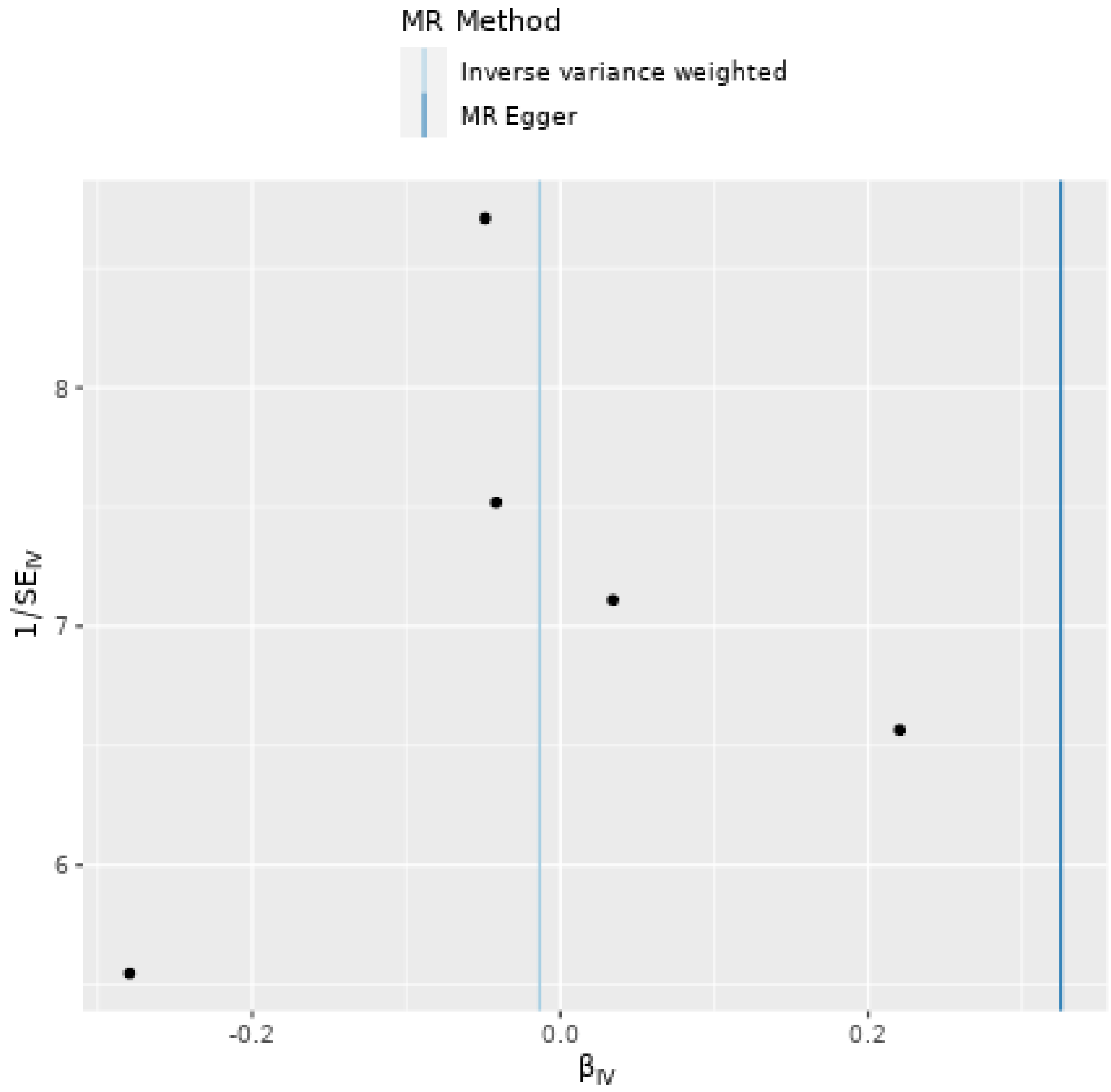

Melanoma as exposure

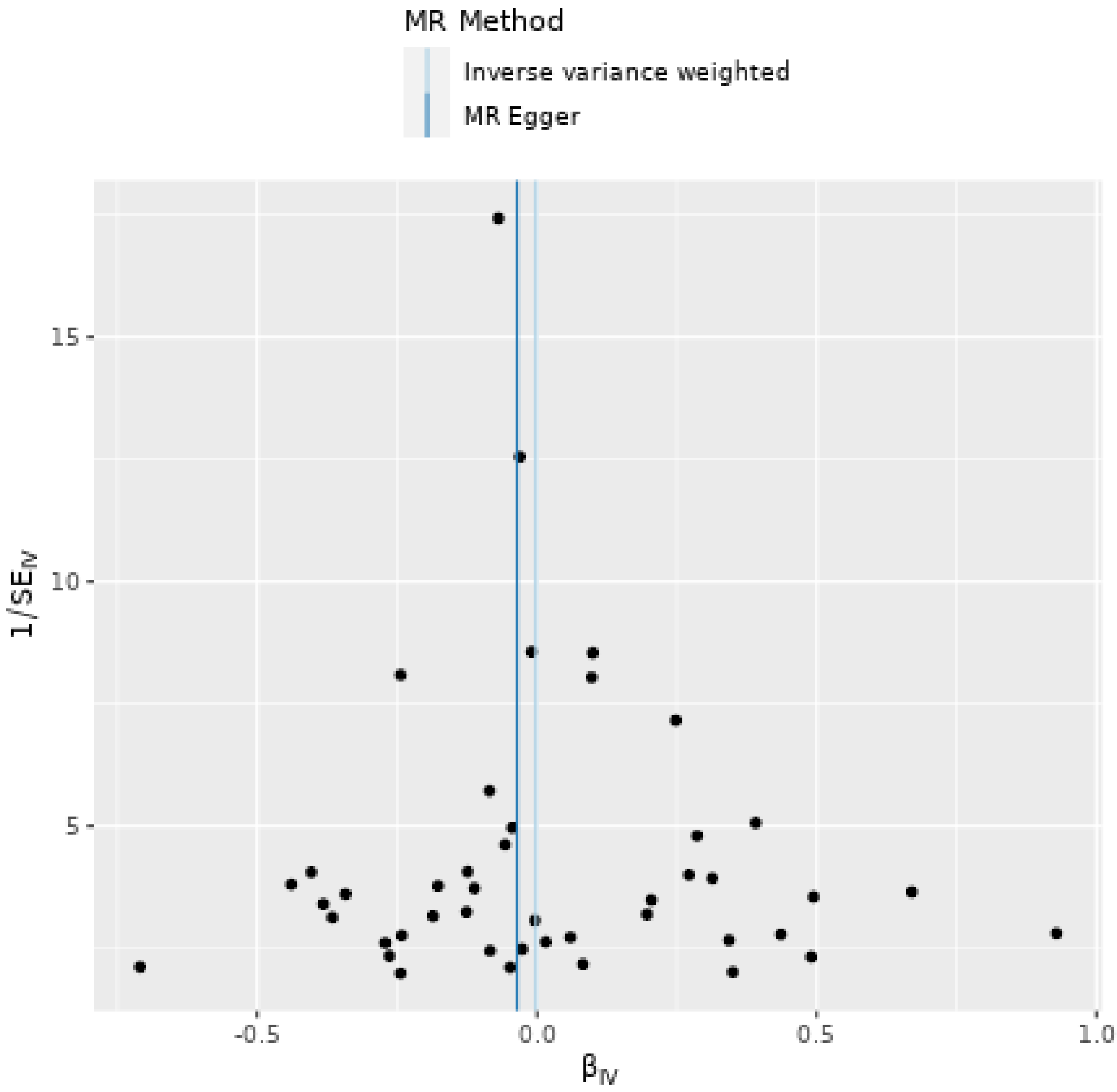

Non-glioblastoma glioma/Glioma as exposure

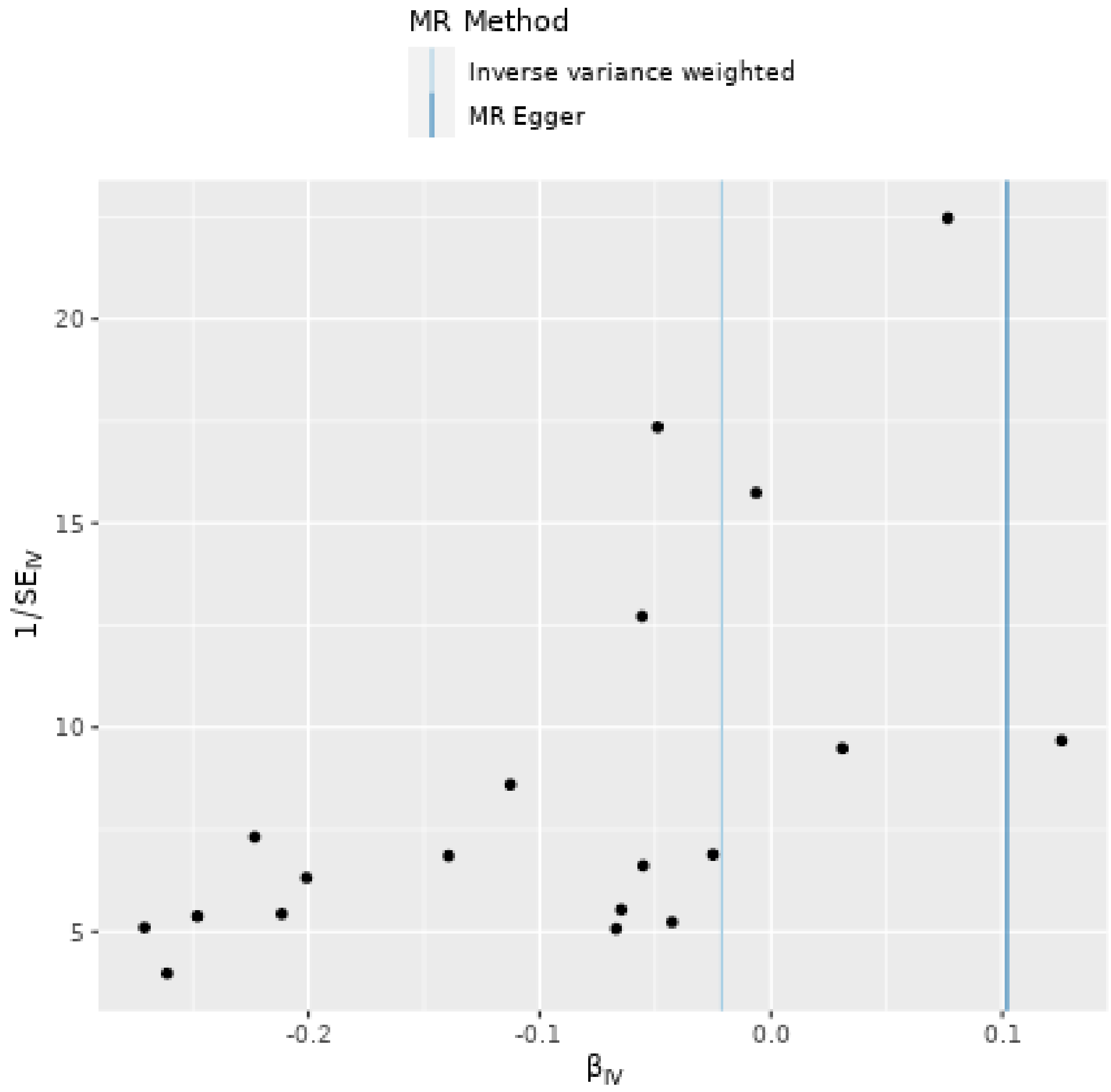

Oral cavity and pharyngeal cancer as exposure

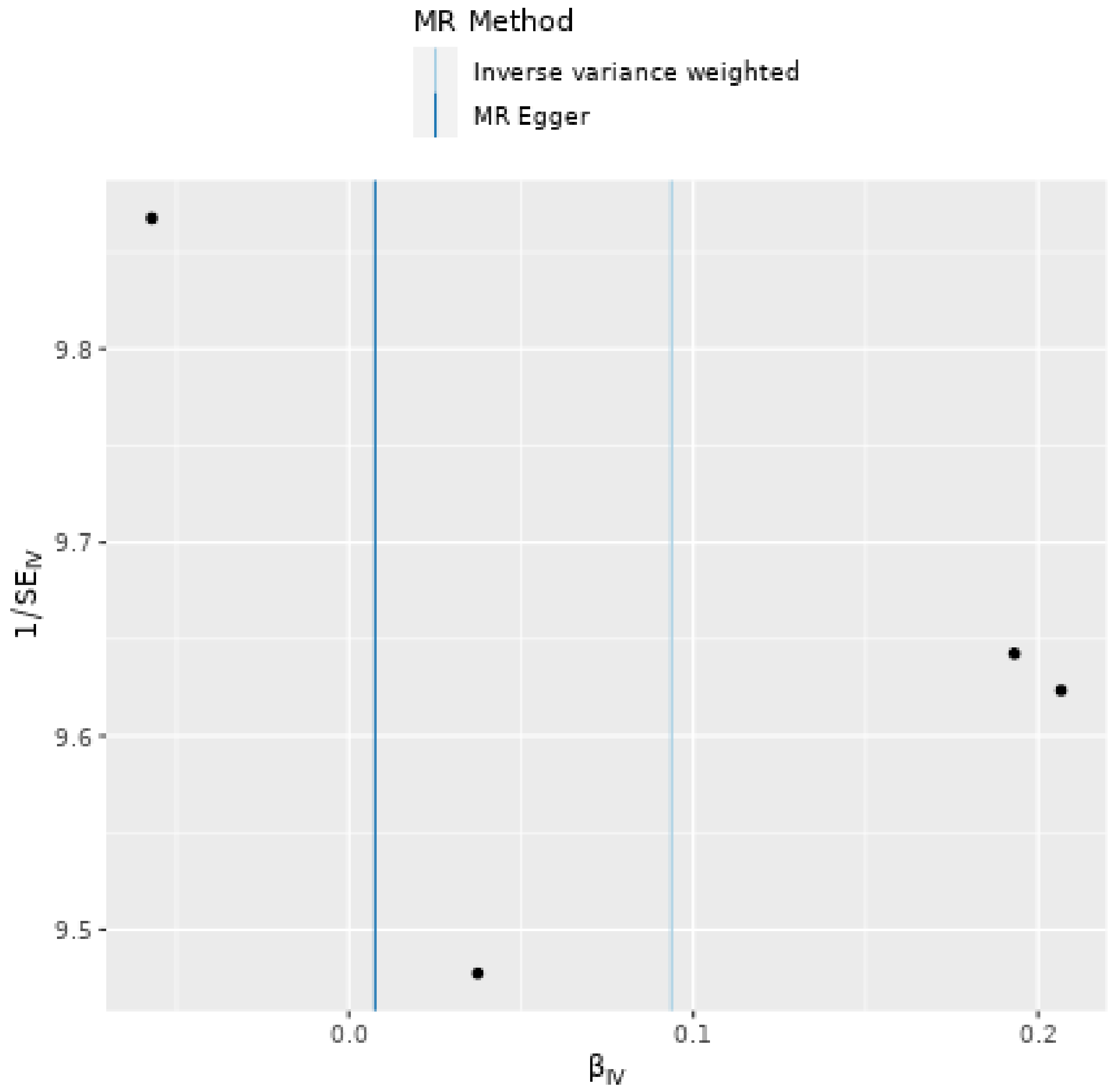

Pancreatic cancer as exposure

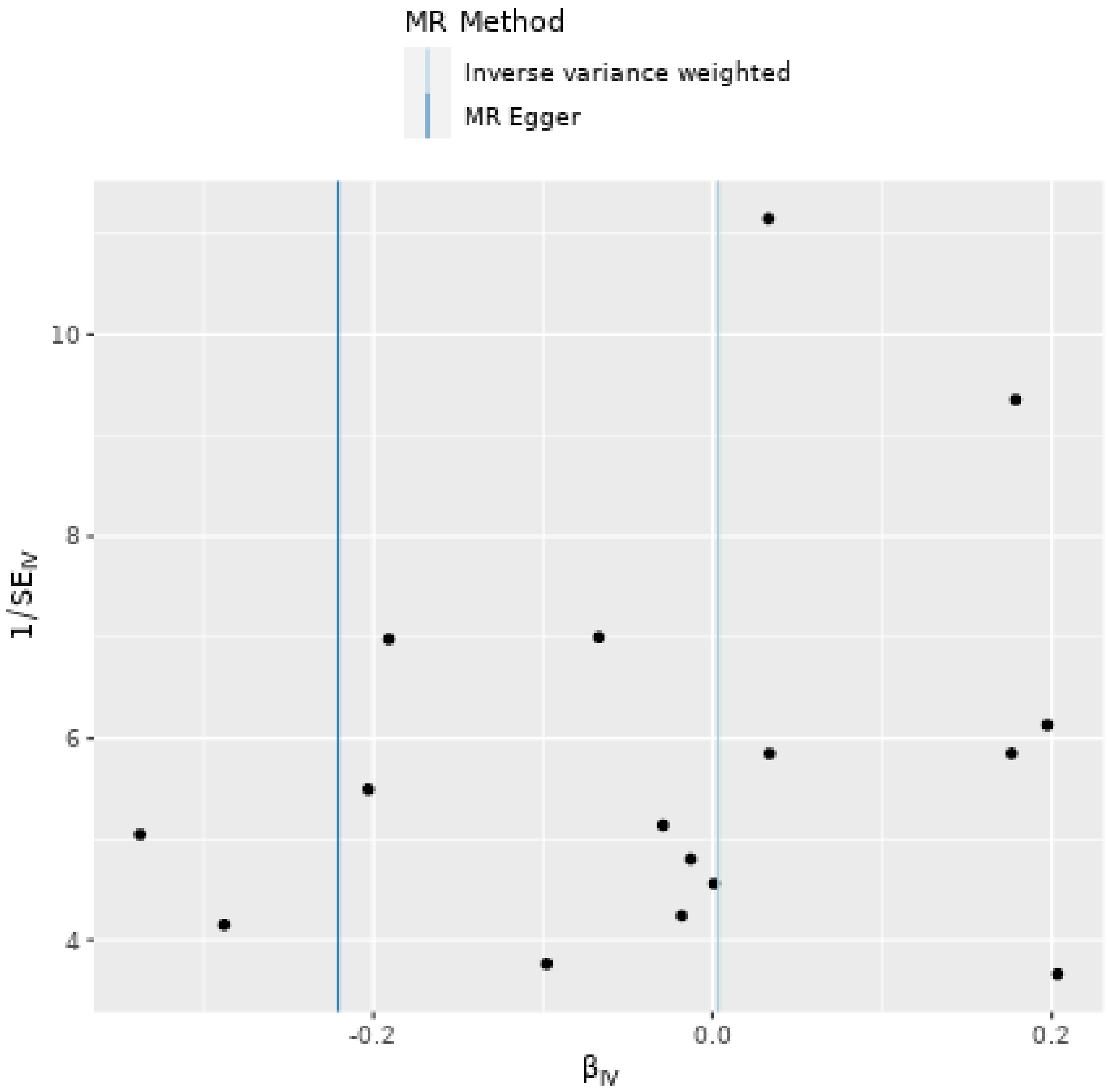

Prostate cancer as exposure

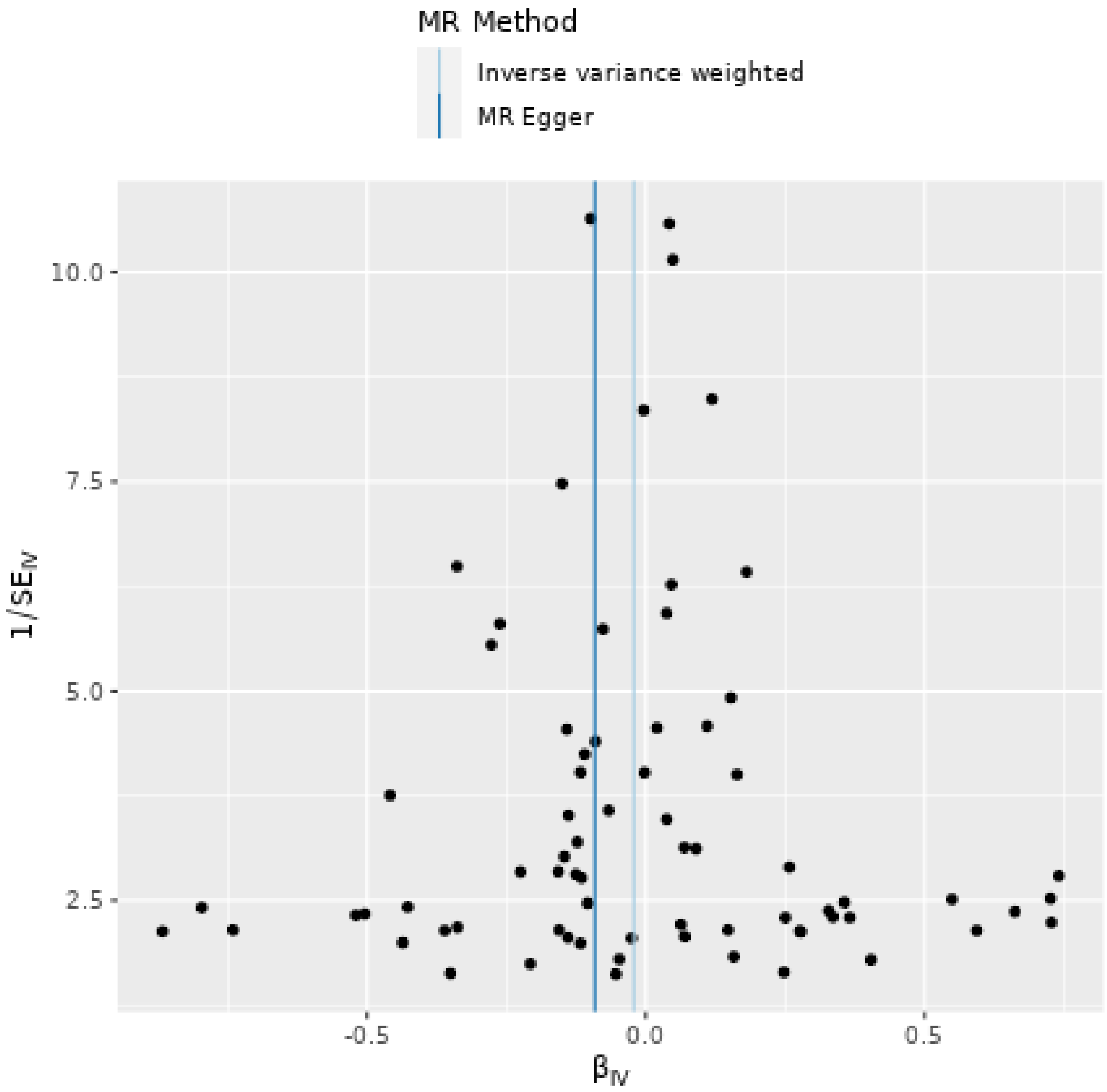

Renal cell carcinoma as exposure

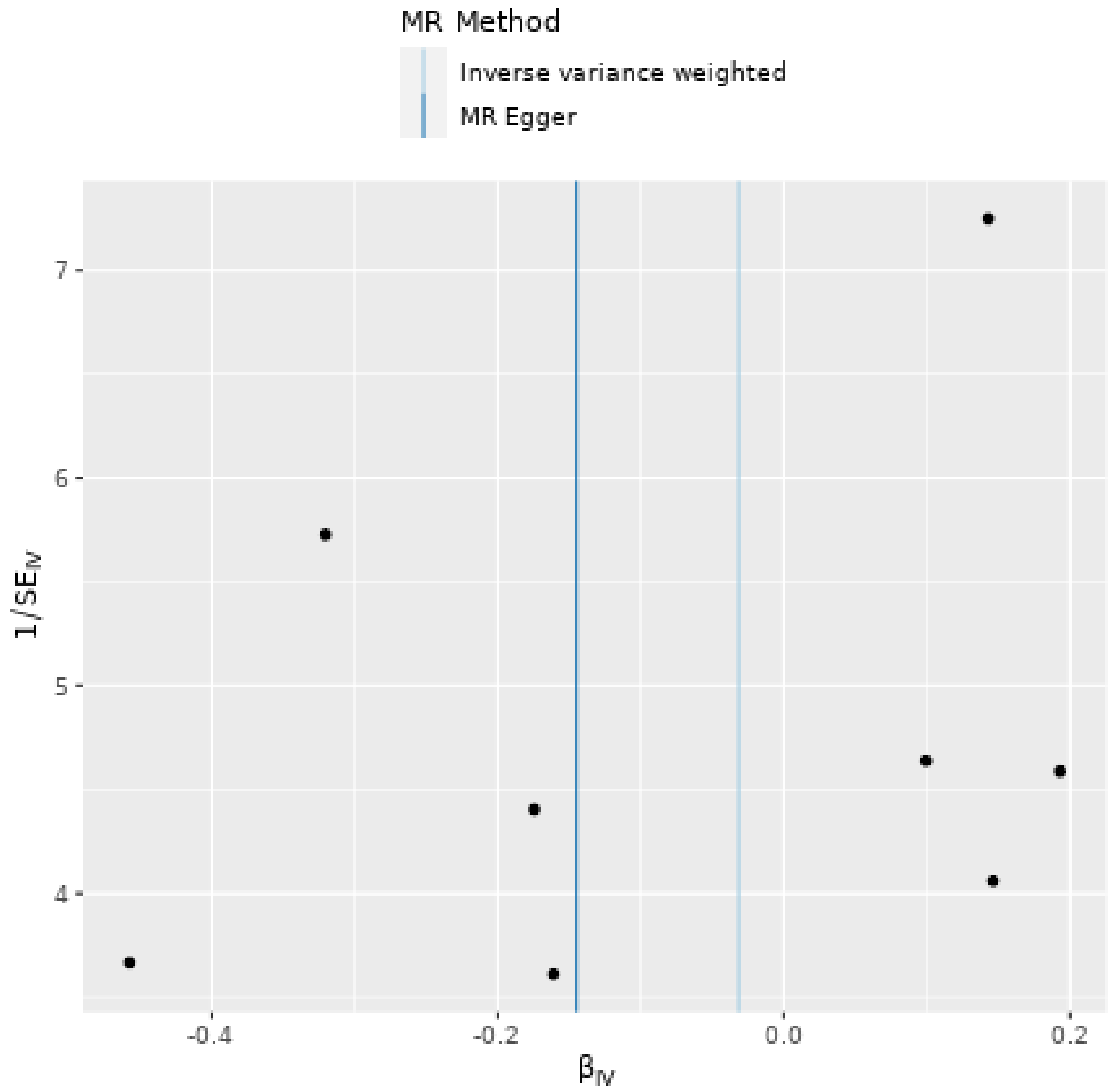

Uterine fibroids as exposure

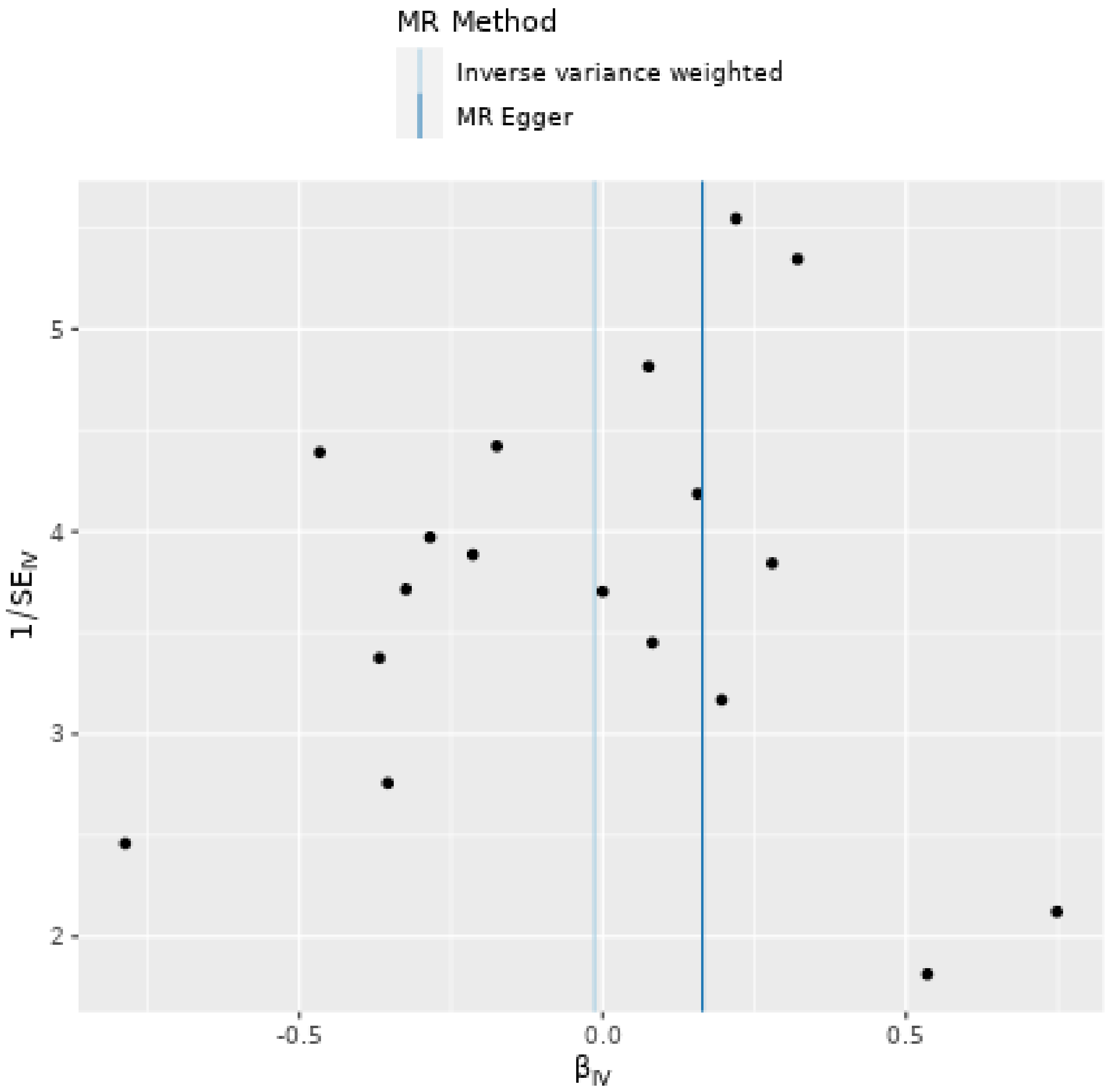

**Supplementary Figure 4. Reverse MR (PD as exposure; Cancers as outcome). Forest plots showing point estimates of the exposures of interest, Exposure of interest at the top of each forest plot**

Breast cancer as outcome

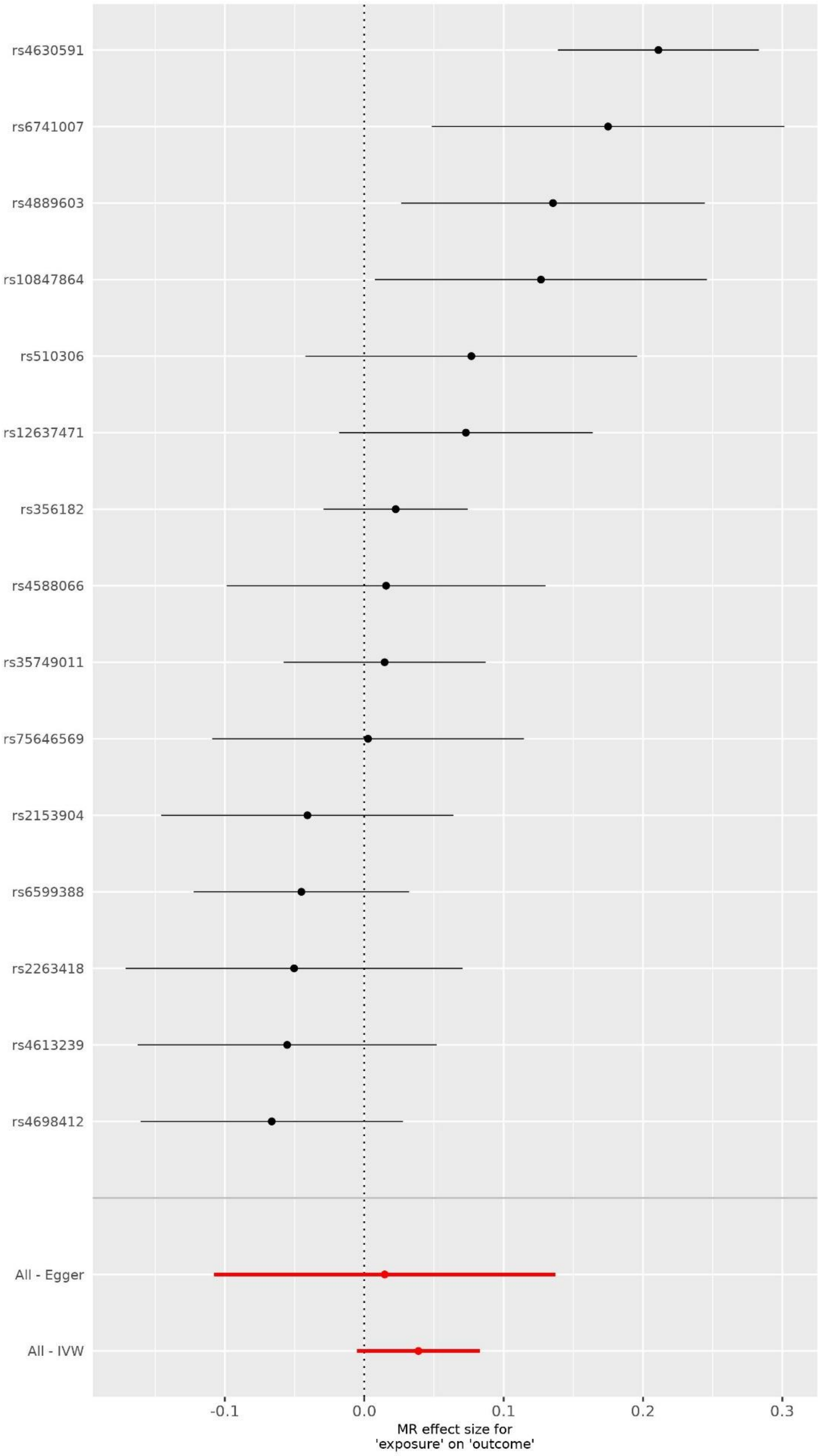

Endometrial cancer as outcome

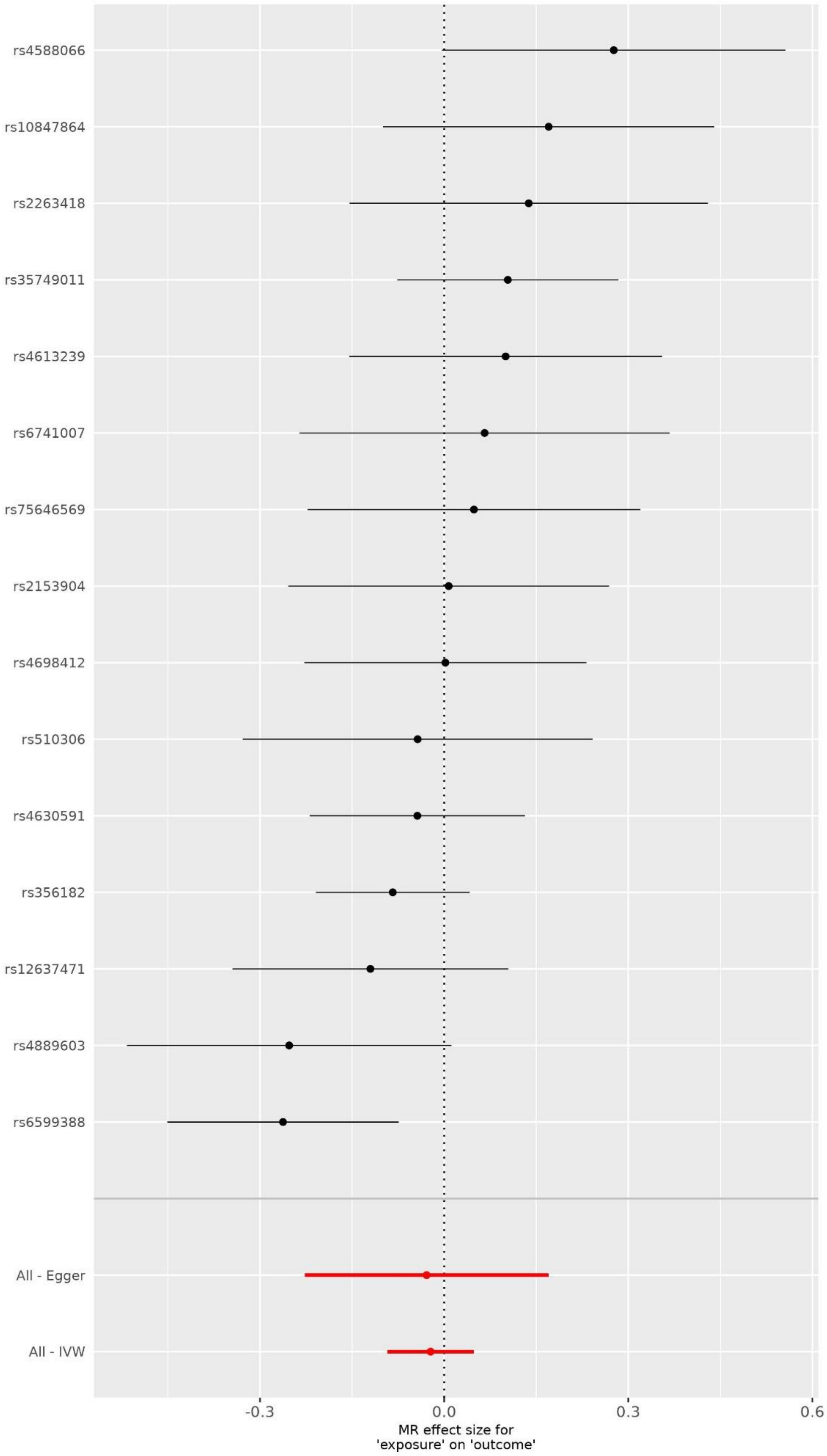

Melanoma as outcome

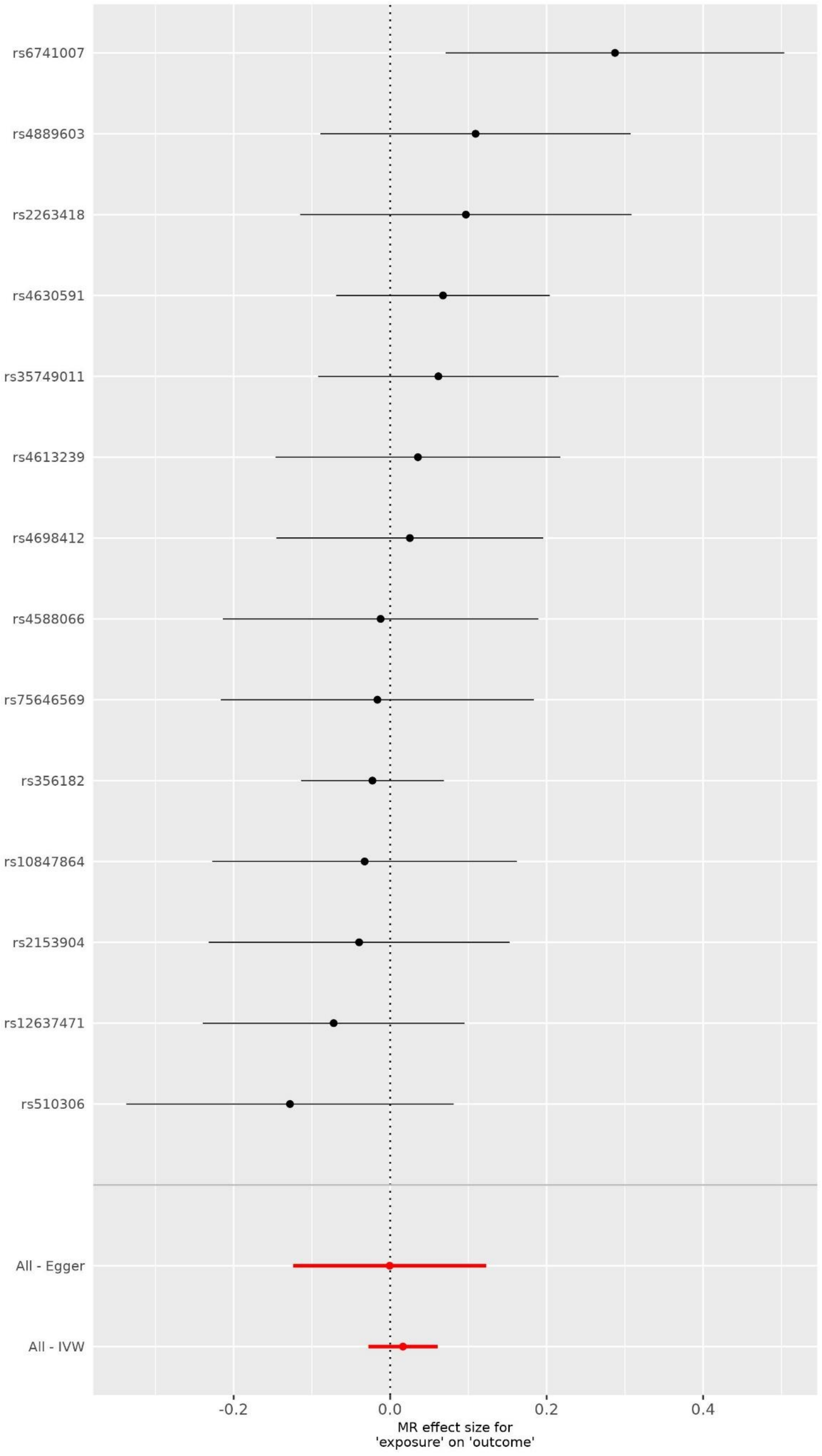

Prostate as outcome

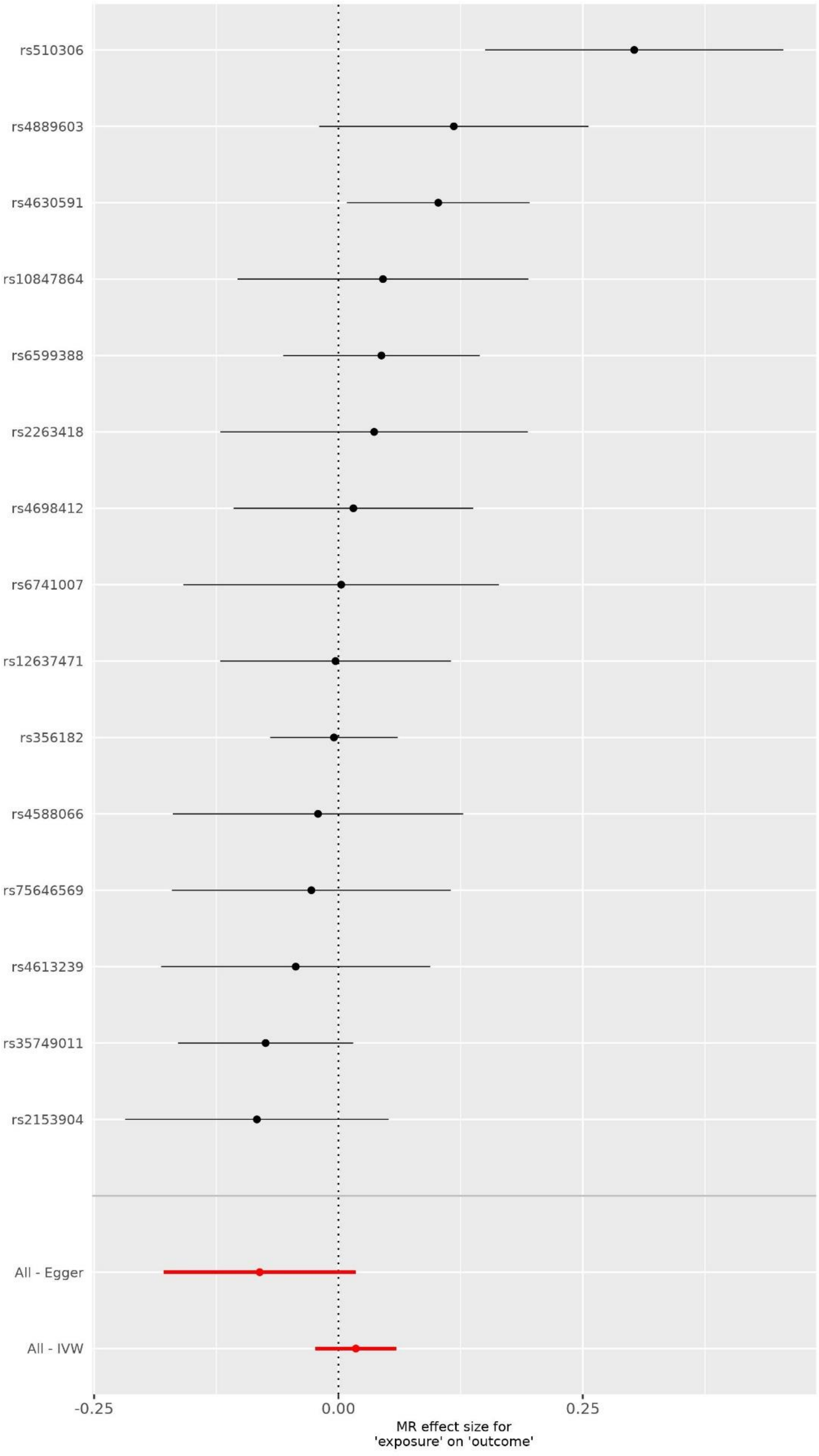

Keratinocytes cancers

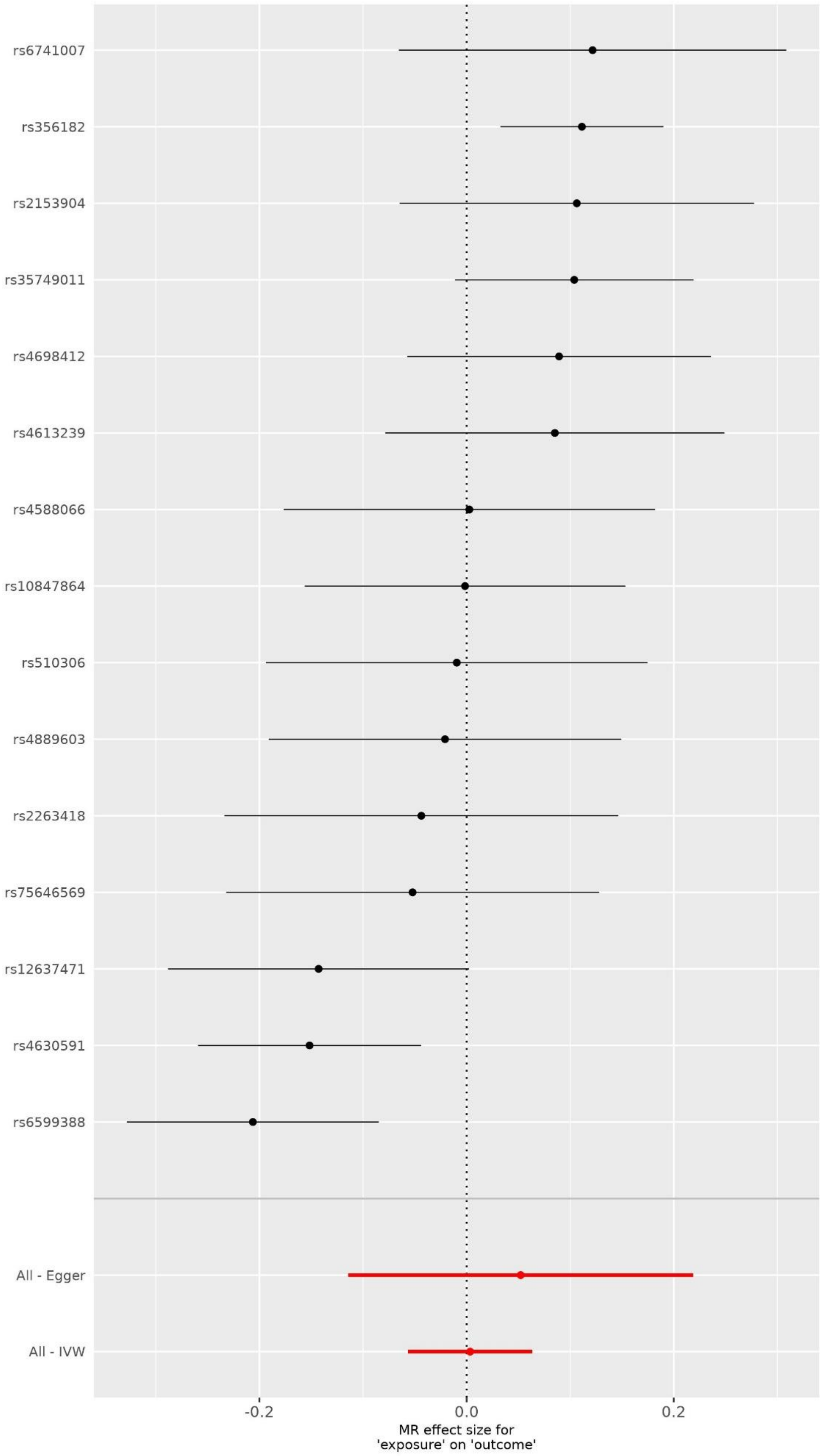

**Supplementary Figure 5. Reverse MR (PD as exposure; Cancers as outcome). Plots showing point estimates of the exposures of interest; Exposure of interest at the top of each plot**

Breast cancer as outcome

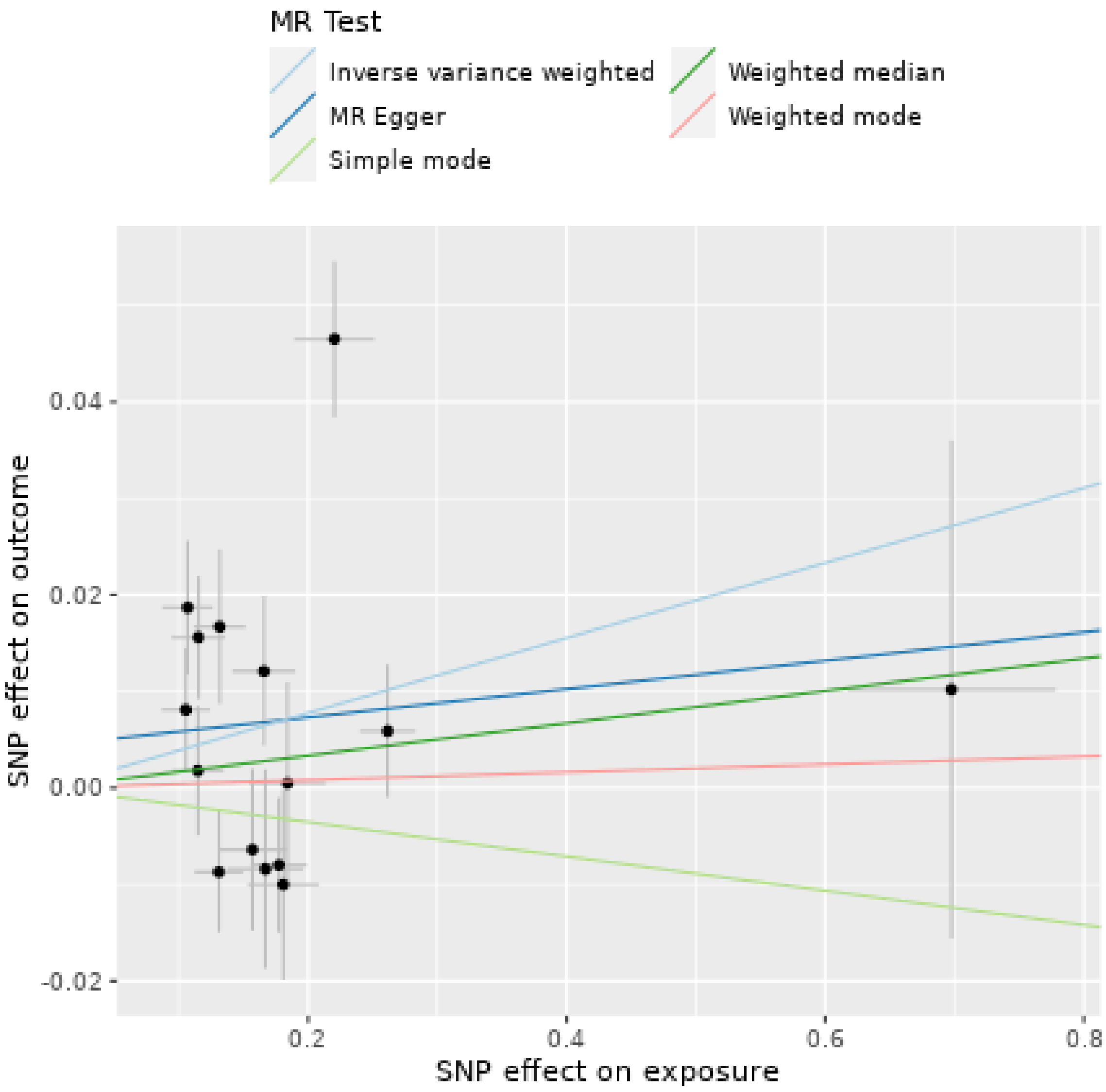

Endometrial cancer as outcome

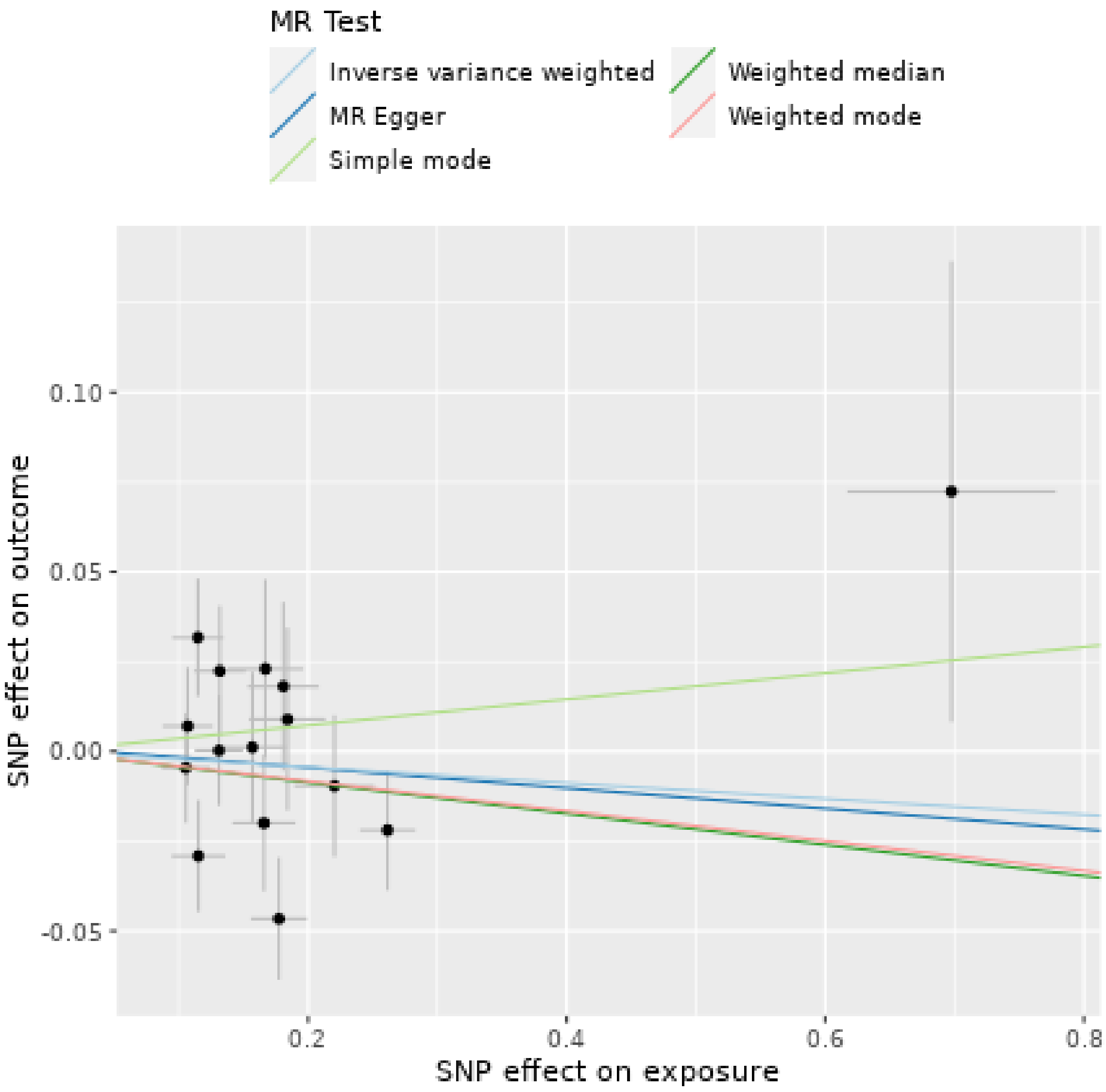

Melanoma as outcome

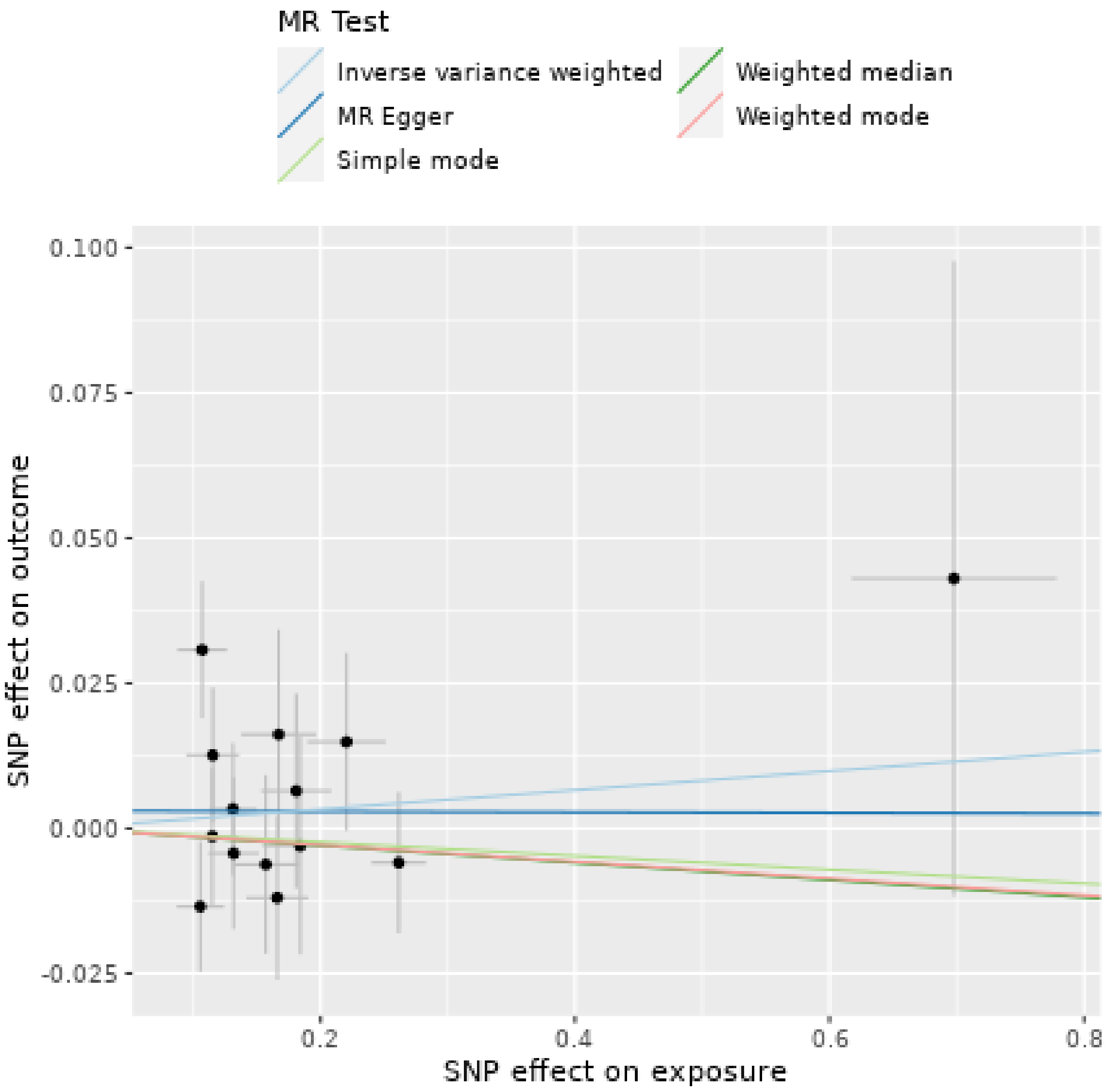

Prostate as outcome

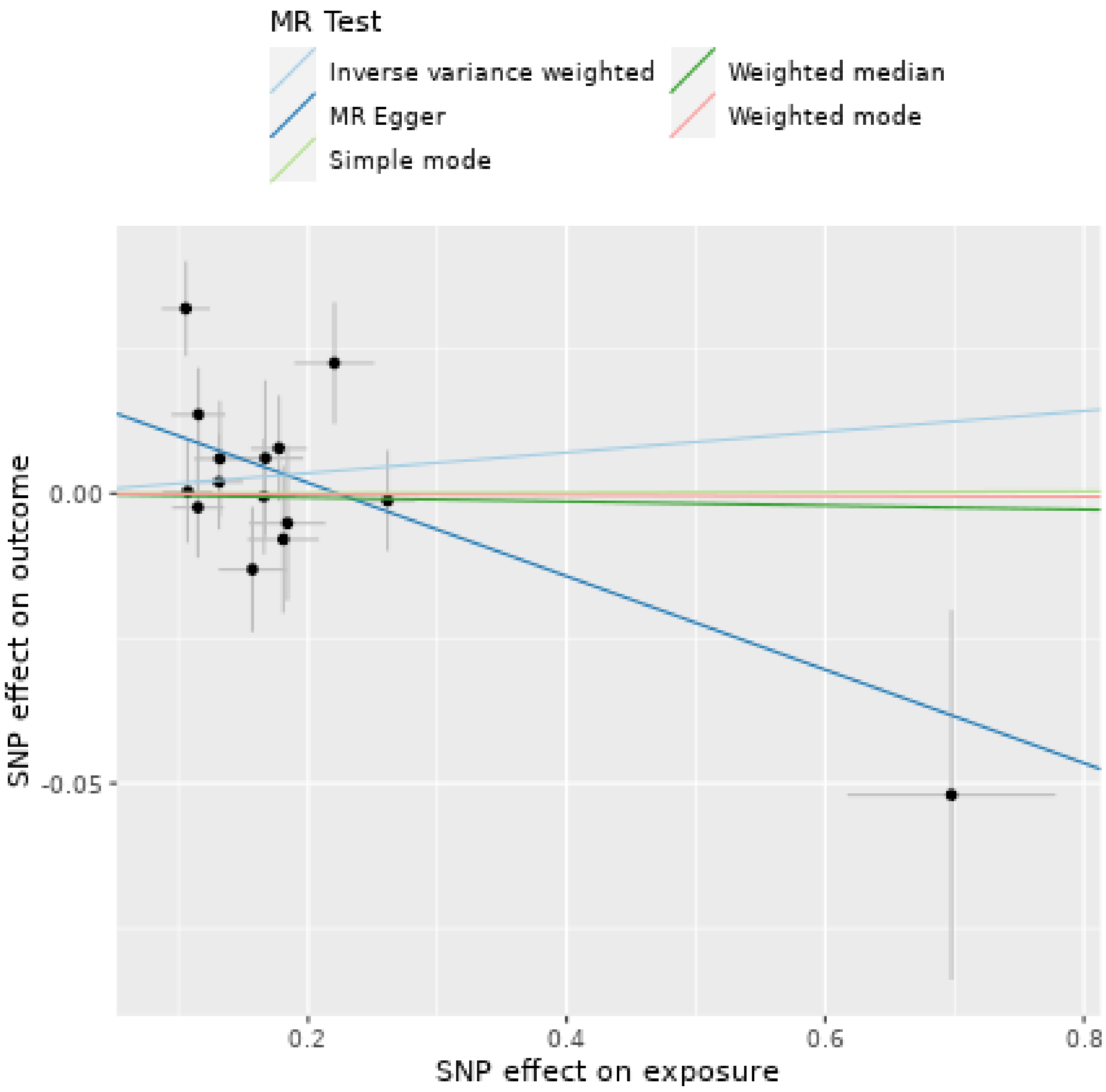

Keratinocytes cancers

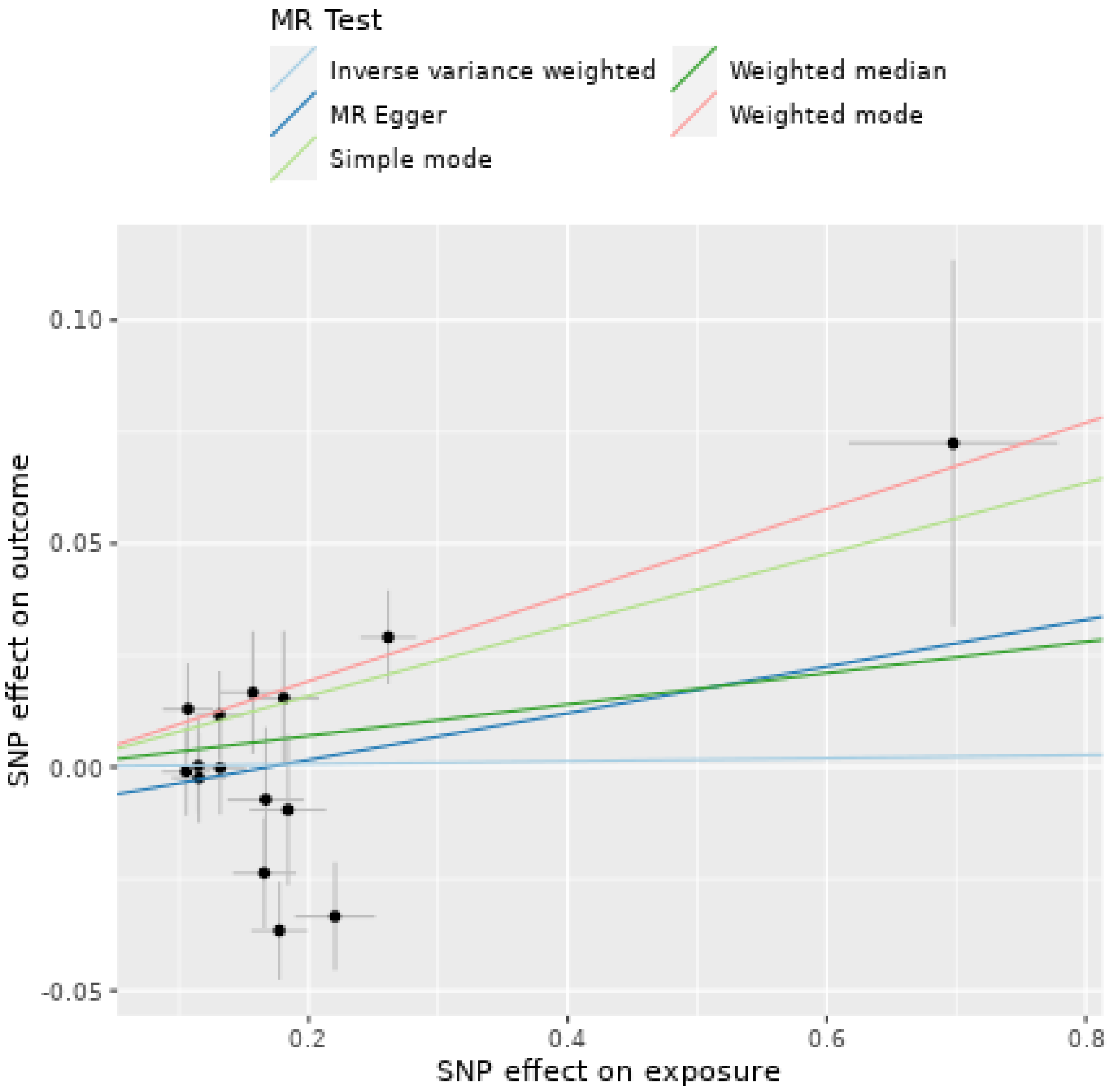

**Supplementary Figure 6. Reverse MR (PD as exposure; Cancers as outcome). Funnel plots evaluated the presence of possible heterogeneity across the estimates. Exposure of interest at the top of each plot**

Breast cancer as outcome

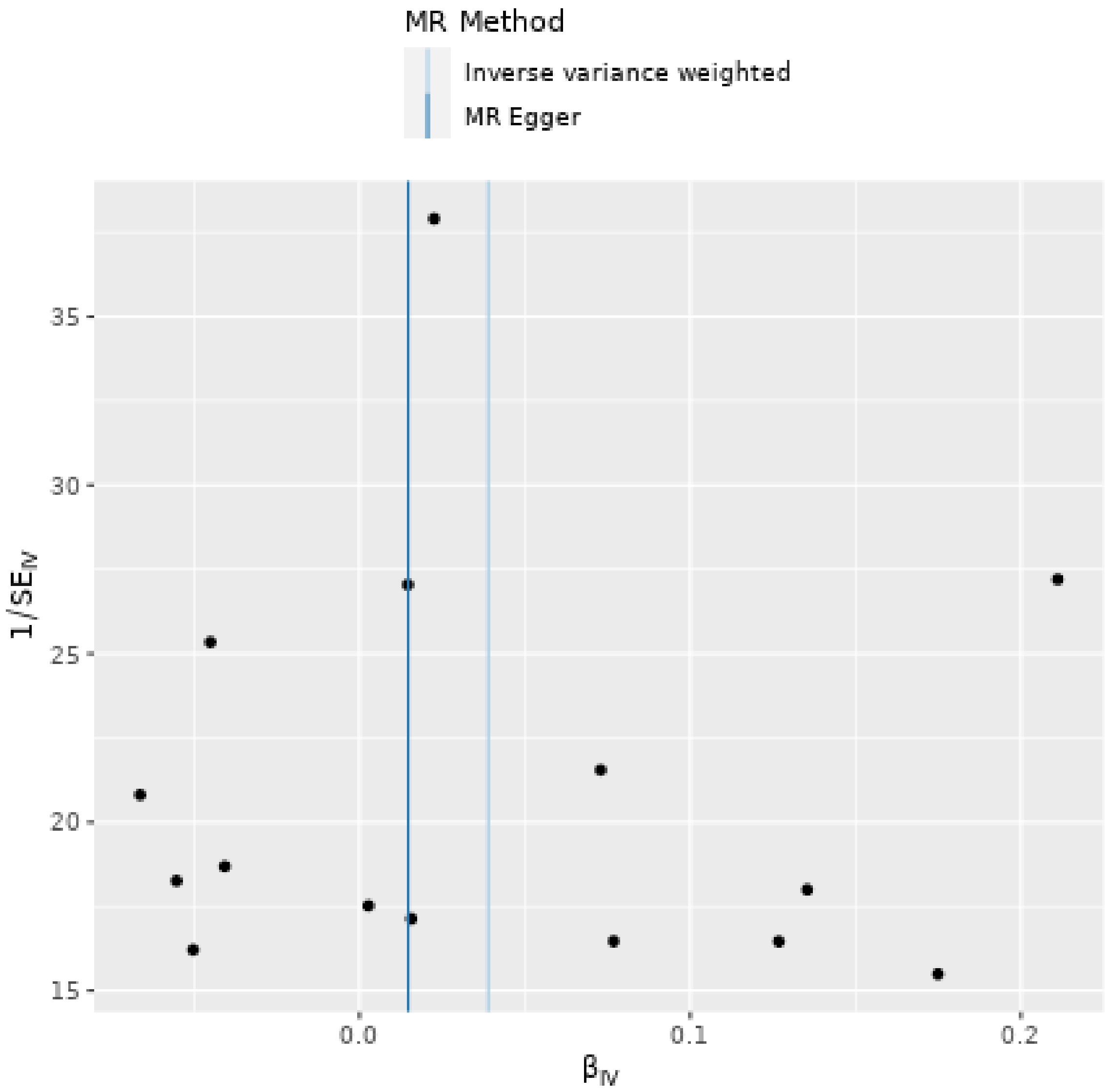

Endometrial cancer as outcome

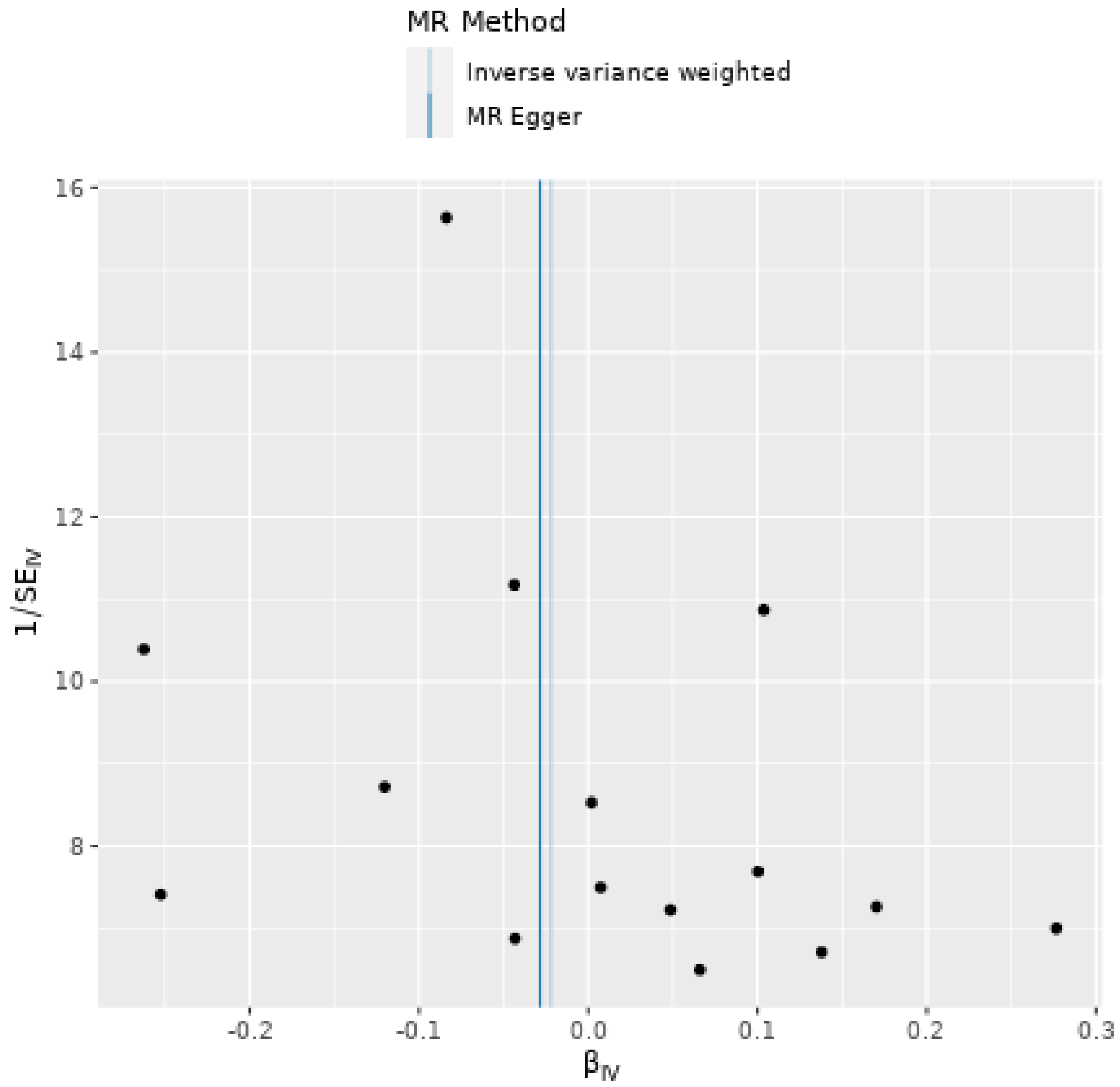

Melanoma as outcome

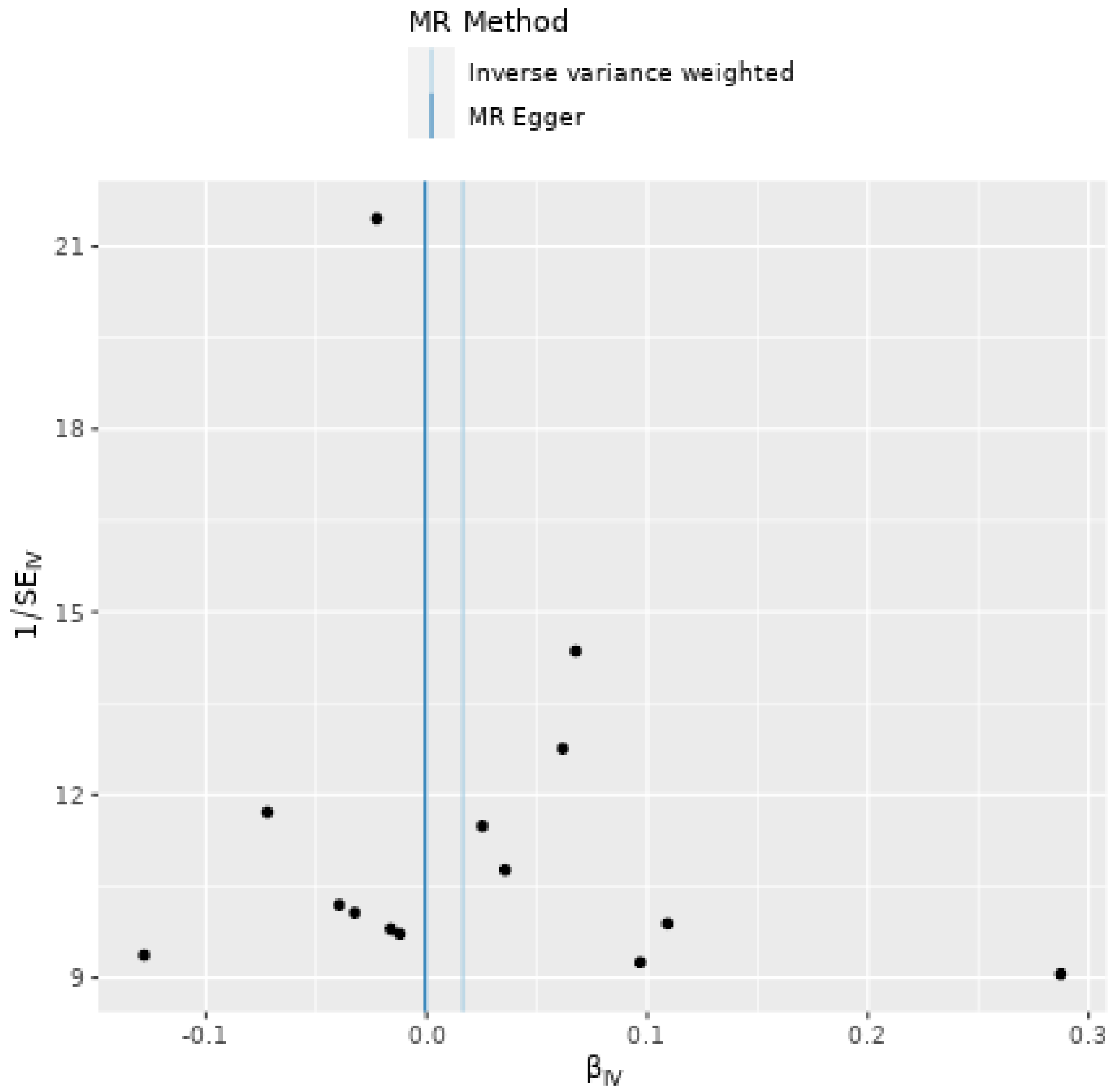

Prostate cancer as outcome

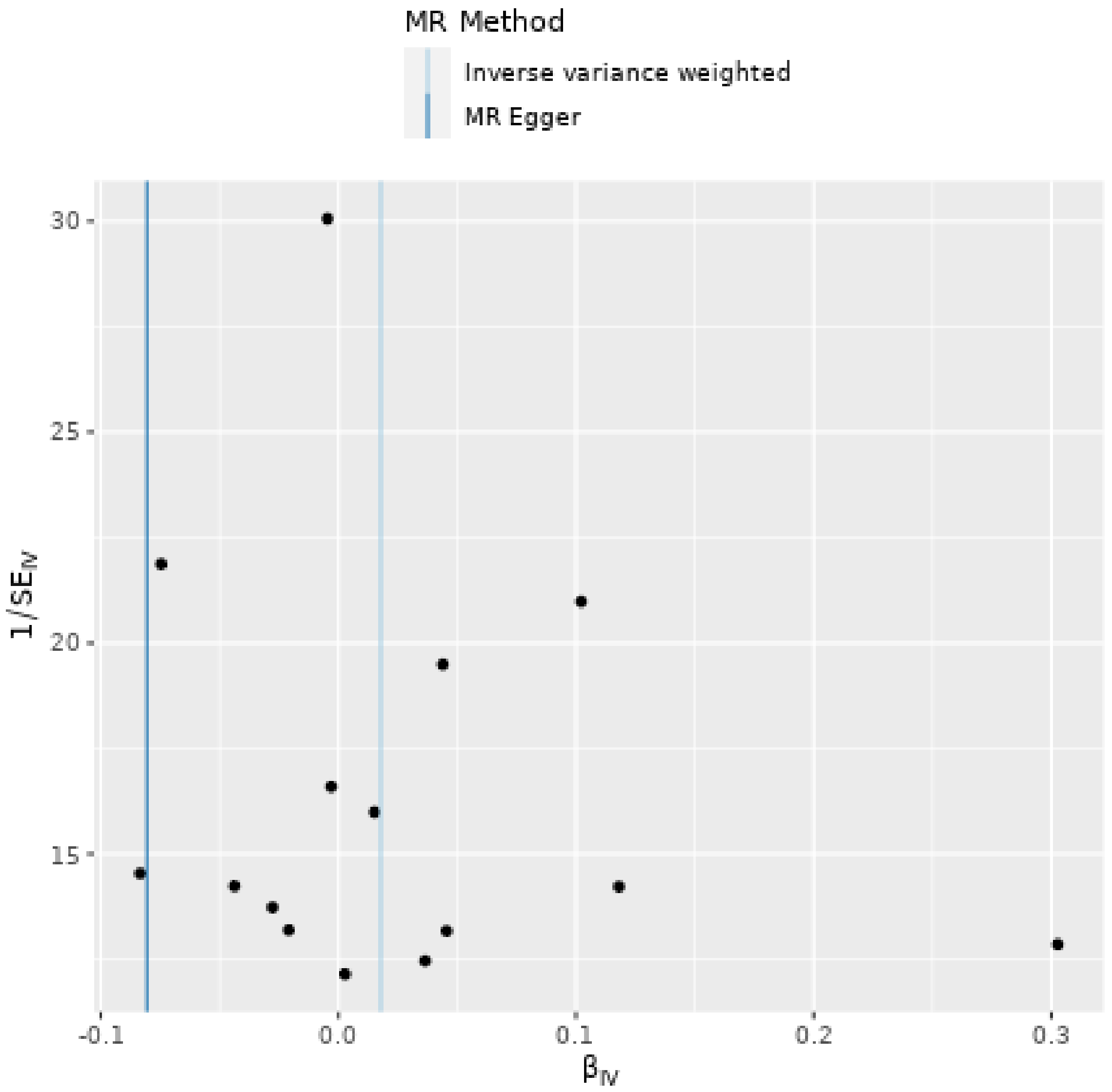

Keratinocytes cancers

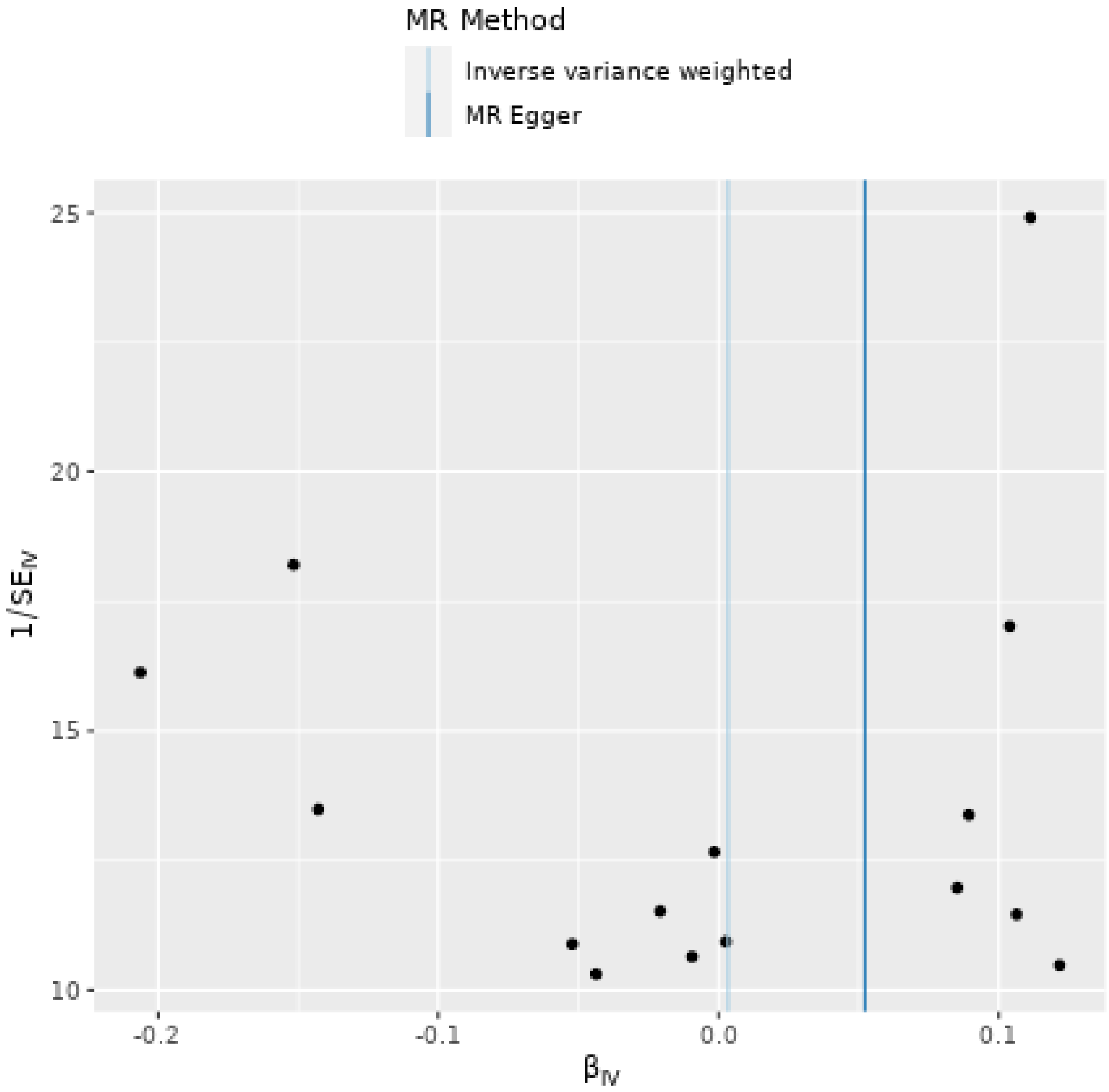
