## Supplementary Table 1 for "Genetic correlation and causality of cancers and Parkinson’s disease"

**Supplementary Table 1. List of all cancer GWAS studies selected for Mendelian randomization analysis**

| **Exposure** | Study | Initial sample size | | Replication sample size | | Power |
| --- | --- | --- | --- | --- | --- | --- |
|  |  | Cases | Controls | Cases | Controls |  |
| **Breast cancer** | Michailidou et al., 2017^1^ | 76,192 | 63,082 | 46,785 | 70,064 | 100.00% |
| **Chronic lymphocytic leukemia** | Law et al., 2017^2^ | 4,478 | 13,213 | 1,722 | 4,385 | 80.70% |
| **Colorectal cancer** | Law et al., 2019^3^ | 31,197 | 61,770 | - | - | 38% |
| **Cutaneous squamous cell carcinoma** | Chahal et al., 2016^4^ | 6579 | 280,558 | 825 | 11,518 | 74.50% |
| **Combined analysis of keratinocyte cancers** | Liyanage et al., 2019^5^ | 31,787 | 619,351 | - | - | 63.00% |
| **Endometrial cancer** | O'Mara et al., 2018^6^ | 12,906 | 108,979 | - | - | 71.50% |
| **Lung cancer** | McKay et al., 2017^7^ | 23,223 | 16,964 | - | - | 71.50% |
| **Lymphoma** | Sud et al., 2017^8^ | 1,278 | 14,325 | 1,586 | 3,069 | 90.60% |
| **Melanoma** | Landi et al., 2020^9^ | 36,760 | 375,188 | - | - | 68.30% |
| **Non-glioblastoma glioma/Glioma** | Melin et al., 2017^10^ | 12,469 | 18,190 | - | - | 93.10% |
| **Oral cavity and pharyngeal cancer** | Lesseur et al., 2016^11^ | 6,009 | 6,585 | - | - | 95.60% |
| **Pancreatic cancer** | Klein et al., 2018^12^ | 9,040 | 12,496 | 2,737 | 4,752 | 82.80% |
| **Prostate cancer** | Schumacher et al., 2018^13^ | 79,148 | 61,106 | - | - | 57.00% |
| **Renal cell carcinoma** | Scelo et al., 2015^14^ | 10,784 | 20,406 | 3,182 | 6,301 | 71.50% |
| **Uterine fibroids** | Rafnar et al., 2018^15^ | 16,595 | 52,3330 | - | - | 64.90% |
